# Post-traumatic stress disorder diagnostic accuracy rates in clinical settings: a systematic review and meta-analysis

**DOI:** 10.64898/2025.12.03.25341390

**Authors:** Angela Harper Gow

**Author notes:** Corresponding author: (AG).

## Abstract

This systematic review assesses the global diagnostic accuracy rate of post-traumatic stress disorder in primary and specialized mental healthcare by analyzing diagnostic accuracy and clinical challenges. Databases were searched for English-language studies published after 2000 that focused on post-traumatic stress disorder diagnostic accuracy, clinician bias, comorbidities, and standardized tools. Risk of bias was evaluated using the Grading of Recommendations, Assessment, Development, and Evaluation guidelines and the Cochrane Risk of Bias Tool for Non-Randomized Studies of Interventions. Studies were grouped by homogeneity into 3 categories for synthesis and meta-analysis: diagnostic accuracy in specialized mental healthcare, accuracy in primary healthcare, and clinician diagnostic capabilities. Forty-seven studies met inclusion criteria: 31 were included in the meta-analysis and 16 in the qualitative synthesis. Of those in the meta-analysis, 18 focused on specialized care, 9 on primary care, 2 reported on both, and 2 examined clinician diagnostic accuracy and bias. Although the 2 studies appearing in both care settings were counted once in the overall tally, they contributed separate estimates to each setting, resulting in 20 analyses for specialized care and 11 for primary care. The weighted mean post-traumatic stress disorder diagnostic accuracy was 55.6% in specialized mental healthcare, 48.2% in primary care, and 41.33% in studies examining clinician diagnostic accuracy and bias. Studies without specific accuracy rates consistently highlighted widespread underdiagnosis in both primary and specialized settings, driven by limited trauma inquiry, inconsistent screening, diagnostic overshadowing due to comorbidities, clinician knowledge gaps, and structural barriers within healthcare systems. Misdiagnosis commonly stems from limited clinician training, time constraints, minimal tool use, comorbidities, and systemic barriers. This review highlights the need for clinician education, routine post-traumatic stress disorder screening in primary care, and integrated mental health services. Diagnostic complexity was frequently linked to symptom overlap, underreporting, and cultural limitations of standard criteria. Longitudinal research is recommended.

## Introduction

In real-world clinical settings, the diagnosis of post-traumatic stress disorder (PTSD) is guided by criteria in the Diagnostic and Statistical Manual of Mental Disorders (DSM) [1] and tools such as the Clinician-Administered PTSD Scale for DSM-5 (CAPS-5) [2]; however, these are often underutilized [3]. Despite the availability of validated instruments like the PTSD Checklist for DSM-5 (PCL-5) [4], the Clinician-Administered PTSD Scale (CAPS) [5], and the Structured Clinical Interview for DSM-5 Disorders (SCID-5) [6], their application in routine clinical practice remains limited, undermining diagnostic accuracy. CAPS-5 is considered the gold-standard for PTSD diagnosis, offering a comprehensive assessment of symptom severity and functional impairment; however, it is time-consuming and requires trained clinicians [7]. The SCID-5 provides a categorical diagnosis based on DSM-5 criteria, while the PCL-5 is a quick, reliable self-report measure aligned with DSM-5 symptoms, making it ideal for screening; however, follow-up interviews are required for a definitive diagnosis [4]. As these tools are not routinely used, many cases of PTSD go undiagnosed or are misdiagnosed, particularly when symptoms overlap with other mental health (MH) conditions, such as anxiety disorders [8]. PTSD is most often identified during psychiatric evaluations prompted by MH symptoms that disrupt daily functioning or well-being, typically following a traumatic event [9]. This highlights the urgent need for routine PTSD screening to enhance diagnostic accuracy and improve patient outcomes [10].

PTSD diagnostic accuracy in clinical settings has been extensively studied; however, challenges with underdiagnosis and misdiagnosis continue to persist. A systematic review conducted in 2023 identified 21 studies that offered valuable insights into PTSD diagnostic accuracy in both primary and specialized care settings. However, given the rapid advancement of PTSD diagnostic tools and the publication of new research since then, an updated review is essential. This review builds upon the original findings by incorporating newly published studies and those previously excluded, reassessing PTSD diagnostic accuracy in real-world healthcare settings.

The clinical classification of trauma types is broad, distinguishing between childhood and adult trauma, with causative factors often playing a critical role in trauma categorization [11]. Consequently, traumatic experiences frequently lead to MH conditions that do not meet PTSD diagnostic criteria, as many individuals report subthreshold or alternative trauma responses that do not meet diagnostic thresholds [12]. While nearly everyone experiences a traumatic event at some point in their lives, PTSD diagnosis remains highly specific, relying on strict criteria for both symptoms and exposure [13]. The minimum diagnostic criteria for PTSD, as outlined in the DSM-5, were established by the Center for Substance Abuse Treatment [14].

Currently, there is a lack of widely used tools [10] for diagnosing PTSD, whether it involves revisions to the DSM-5 criteria [15] or other diagnostic instruments [3]. As a result, ensuring real-world diagnostic accuracy for PTSD primarily depends on identifying the rates of missed or misdiagnosed cases in clinical settings. The accuracy of PTSD diagnosis relies on the clinician’s expertise, and it is essential that patients provide truthful and comprehensive responses to the clinician’s questions. However, data on the real-world accuracy of PTSD diagnoses are limited. Studies that focus on the DSM-5 criteria and related diagnostic tools, such as the SCID-5 and CAPS-5, typically examine aspects like treatment effectiveness or the validity of measures rather than diagnostic accuracy. Additionally, these studies often involve highly skilled clinicians, which may not reflect the performance seen in everyday clinical practice. Nonetheless, evaluating the performance and accuracy of new PTSD diagnostic tools under real-world conditions is essential for making valid comparisons with existing methods in both primary and MH settings.

There is also ongoing disagreement in the field regarding the optimal thresholds for PTSD diagnosis across diverse populations [16], the sensitivity and clinical applicability of DSM-based criteria [17, 18], and the relative value of structured versus unstructured diagnostic tools [19]. These controversies have led to varied practices and inconsistent detection rates in clinical settings, which this review aims to clarify.

The objective of this review was to assess the accuracy of PTSD diagnoses in real-world settings by investigating studies conducted in primary care and MH clinics. This assessment aimed to minimize bias by including studies where initial diagnoses were not part of clinical research but were derived from existing medical records. The review employed accepted reference standards, such as DSM-based structured clinical interviews, to determine the actual PTSD status. In addition to direct measures, the review incorporated studies with diverse characteristics, including those focusing on primary or secondary outcomes related to PTSD, the use of diagnostic tools in healthcare, comorbid illnesses with overlapping symptoms, and clinician bias.

The study objectives were to highlight the current need for improving PTSD diagnostic accuracy and to achieve the highest level of evidence, designated as Level I by the Australian Government’s National Health and Medical Research Council [20], through a systematic review of the existing literature on PTSD diagnostic accuracy in clinical settings.

## Materials and methods

### Search strategy and criteria

This systematic review updates a previously conducted review on PTSD diagnostic accuracy in clinical settings. The original review, completed in June 2023, identified 21 studies that met the inclusion criteria. The search strategy at that time included databases such as PubMed, PTSDpubs, ProQuest, and ScienceDirect, focusing on studies published after 2000.

To update the review, a second systematic search was conducted between December 2024 and July 2025, using the same databases and search terms as the original review. The aim of this updated review was to identify newly published studies since June 2023, as well as studies that may have been missed in the initial review but that met the inclusion criteria. This updated review reassesses PTSD diagnostic accuracy in both primary healthcare and specialized MH settings.

Additional databases and sources were explored to capture new evidence that may not have been available during the initial review. The Preferred Reporting Items for Systematic Reviews and Meta-analyses (PRISMA) [21] flowcharts ((Fig.1A and 1B) detail the study selection process for both the original and updated searches.

**Fig 1A and 1B.**
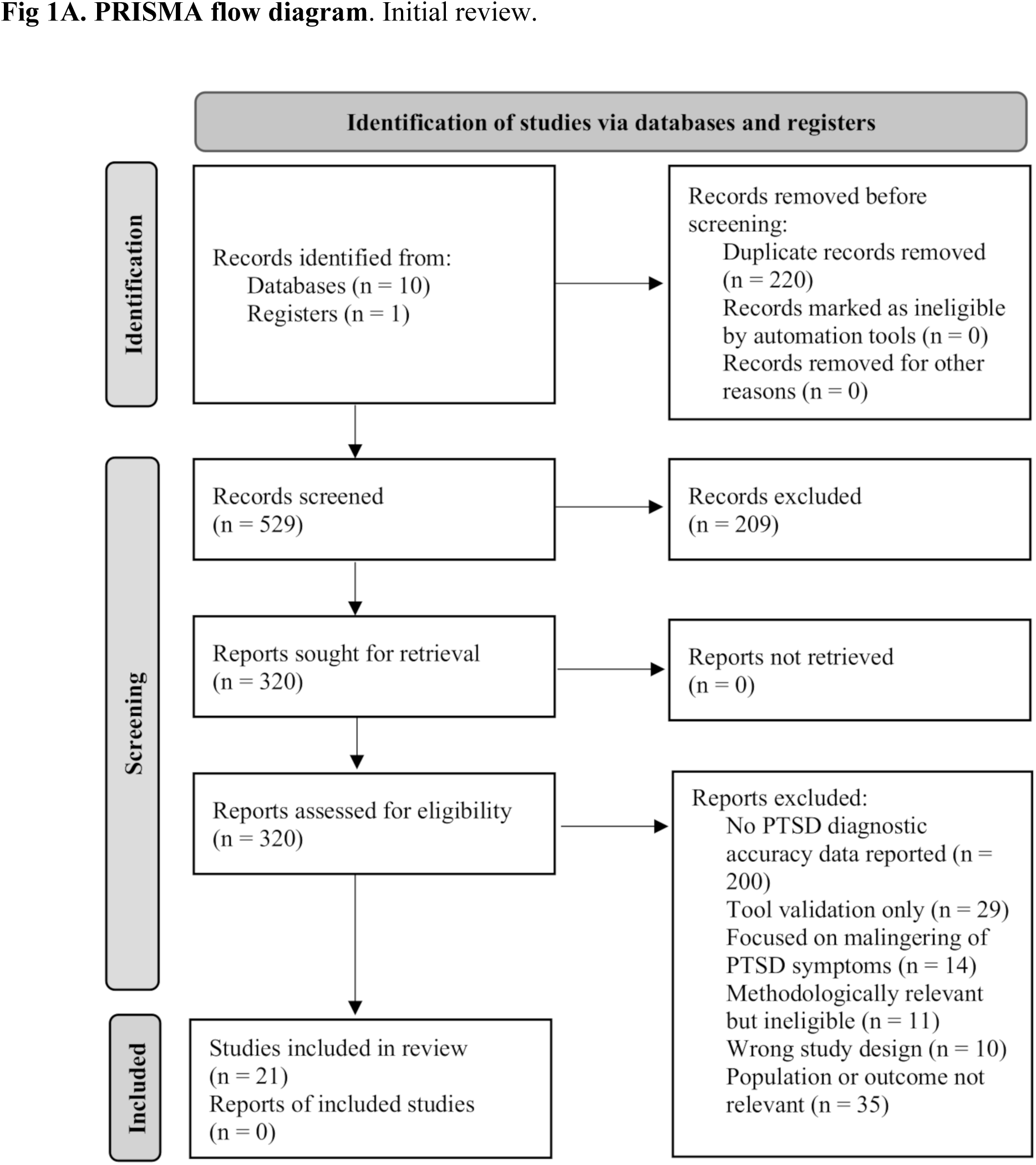
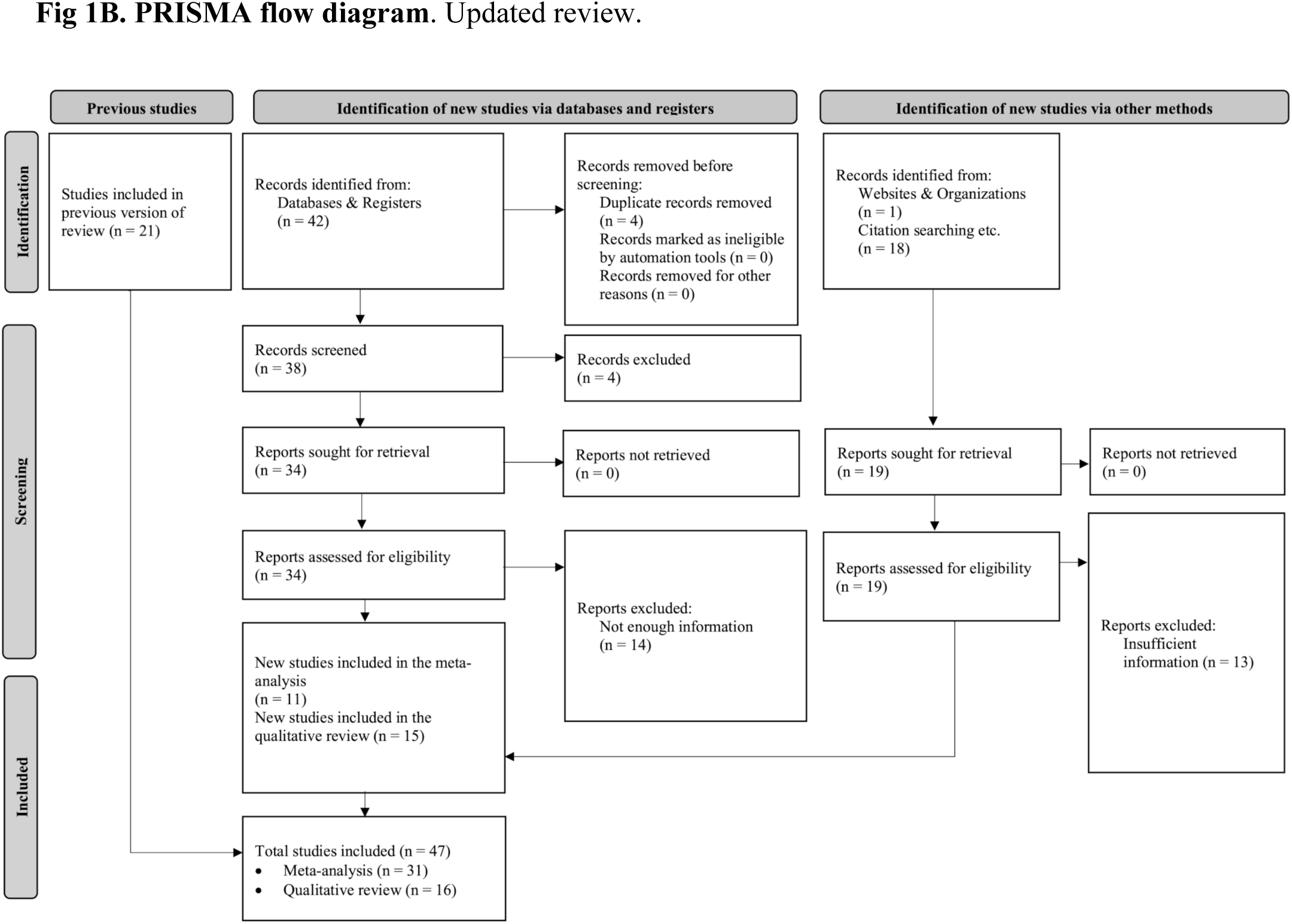
PRISMA flow diagram. PRISMA = Preferred Reporting Items for Systematic Reviews and Meta-Analyses. Source: Page et al. [21].

In addition to major electronic databases, this review update also utilized the consensus database to identify additional studies related to PTSD diagnostic accuracy. A citation-searching approach was also employed, which involved reviewing the following:

- The reference lists of studies previously included in the original review
- The reference lists of newly included studies from the updated search
- The reference lists of studies initially identified via consensus but later excluded after full-text screening

This iterative approach ensured that relevant research that may not have been captured in the initial search was considered for inclusion. Search terms included “PTSD” [AND] “Diagnosis” [OR] “missed diagnosis” [OR] “underdiagnosis” [OR] “overdiagnosis” [OR] “misdiagnosis.” This search was complemented by reviewing bibliographies and websites, such as the National Center for PTSD [22].

This review focuses on assessing real-world PTSD diagnostic accuracy, including studies in which participants received diagnoses in primary care or specialized MH clinics. To minimize bias, only studies with initial diagnoses derived from existing medical records or routine clinical assessments were included, rather than those based on research-driven evaluations. PTSD status was determined using accepted reference standards, such as DSM-based structured clinical interviews (e.g., CAPS, Structured Clinical Interview for DSM-IV [SCID]) or validated PTSD screening tools (e.g., Primary Care PTSD Screen for DSM-5 [PC-PTSD-5], PTSD Checklist [PCL]). Unlike the original review, all newly included studies directly measured PTSD diagnostic accuracy; none assessed indirect variables such as clinician bias or symptom misattribution. One newly included study (Gravely et al. [23]) reported separate diagnostic accuracy results for both primary care and specialized MH settings. Therefore, it was listed under both categories in the summary tables but counted only once in the total number of studies. This study used positive predictive value (PPV) as its diagnostic metric, which was considered appropriate for real-world accuracy due to its reliance on routine clinical diagnoses, but was not included in pooled sensitivity estimates or forest plots because PPV is not directly comparable to sensitivity measures. Similarly, Bohnert et al. [24], initially included in the primary care category, were later divided and presented in both the primary care and specialized care sections during the updated review. This adjustment was made because the study reported distinct diagnostic accuracy rates for primary care alone, integrated primary care–MH services (PC-MHI), and specialty MH services. As PC-MHI involved licensed MH professionals embedded within primary care clinics, it was considered comparable to specialized care for analysis purposes.

#### Literature inclusion criteria

- Research Designs: Literature reviews, observational studies, and experimental studies
- Variables:
  - Group A: PTSD diagnostic accuracy in clinical settings and the impact of clinician bias on PTSD diagnosis.
  - Group B: Prevalence of comorbidities in individuals with PTSD, delayed official PTSD diagnoses in individuals with previously diagnosed comorbidities, diagnostic challenges in PTSD, and missed PTSD diagnoses due to comorbidities.
  - Group C: Standard diagnostic tools routinely used in MH, the efficiency of these tools for PTSD diagnosis, time-related and other constraints in MH affecting diagnostic accuracy, and rates of PTSD-trained clinicians among MH professionals.
- Publication Date: After 2000
- Language: English (including translated studies)

No other variables were sought beyond those directly relevant to PTSD diagnostic accuracy, such as clinical setting, comorbidities, clinician-related factors, and diagnostic tool use.

#### Literature exclusion criteria

- Research Designs: Animal research studies, ideas, editorials, and opinion pieces
- Exclusion of studies that did not include any variables from Groups A, B, or C
- Publication Date: Before 2000
- Language: Any language other than English

Of the 21 studies included in the original review, 10 were conducted in the US, 3 in the UK, and 1 each in Australia, South Africa, the Netherlands, Israel, Greece, and Brazil. Additionally, 2 systematic reviews were included [25, 26], which together analyzed data from 56 studies. The review by Zammit et al. [25] included 29 studies: 15 from the US, 3 from Australia, 3 from the UK, 3 from the Netherlands, 2 from Germany, and 1 each from South Africa, Spain, and Turkey. The review by Greene et al. [26] included 27 studies, with 24 conducted in the US, 2 in Israel, and 1 in South Africa.

As part of the updated review, 11 newly identified studies were included in the meta-analysis, and 1 study [26] from the original meta-analysis was reclassified into the qualitative synthesis due to overlapping data. Additionally, 15 new studies were added to the qualitative section. This brought the total number of included studies in the review to 47, comprising

- 31 studies included in the meta-analysis
- 16 studies included in the qualitative synthesis

Of the 31 studies in the meta-analysis, 19 were conducted in the US, 3 in the UK, 2 in South Africa, and 1 each in Australia, Israel, Greece, Switzerland, the Netherlands, and Brazil. Additionally, there was 1 systematic review [25] comprising 29 studies: 15 from the US, 3 from Australia, 3 from the UK, 3 from the Netherlands, 2 from Germany, and 1 each from South Africa, Spain, and Turkey.

Of the newly identified studies included in the meta-analysis, 9 were conducted in the US, and 1 each was conducted in Switzerland and South Africa.

The 16 studies included in the qualitative synthesis comprised 1 systematic review reclassified from the original meta-analysis [26], 7 new US studies, 3 studies from Canada, 2 from the UK and 1 each from, China, and Israel. Additionally, a new systematic review [27] was included in the qualitative review comprising 41 studies, 29 of which were conducted in the US, 2 each in Israel and South Africa, and 1 each in Canada, Italy, Lithuania, Puerto Rico, Qatar, Spain, Sweden and Switzerland.

In the original review, Covidence—a web-based collaboration software platform designed to streamline the production of systematic and other literature reviews—was used for study screening, selection, and data extraction [28]. Of the 749 studies identified for screening, 220 were excluded due to duplication, irrelevance, or other errors. After the initial screening, 529 studies were retained based on relevant keywords such as “PTSD,” “diagnosis,” and “misdiagnosis.” However, 209 studies were excluded during the abstract screening phase because they did not meet key inclusion criteria, such as PTSD diagnostic accuracy, clinician bias, or challenges related to comorbidities and diagnostic tools in healthcare settings. This led to the review of 320 full-text articles, of which 21 were ultimately included.

As part of the updated review, an additional 61 studies were identified, including newly published studies and others that had been previously missed in the original search. After applying the same eligibility criteria, 26 of these studies were included in the updated review, 11 of which were incorporated into the meta-analysis, and 15 were included in the qualitative synthesis. Additionally, 1 systematic review [26] that was originally included in the meta-analysis was moved to the qualitative synthesis due to study overlap with the newly included studies [29, 30, 31, 32, 33, 34]. This brought the total number of included studies across both the original and updated reviews to 47, comprising 31 studies in the meta-analysis and 16 in the qualitative synthesis. To ensure transparency in the study selection process, both the original PRISMA diagram (Fig. 1A) and the updated PRISMA diagram (Fig. 1B) are presented. The detailed PRISMA [21] flow diagrams illustrating the selection process are shown in Fig. 1A and 1B.

Studies were grouped into 3 categories for the synthesis and meta-analysis to ensure an appropriate degree of homogeneity. The distribution of the groups was as follows:

Group 1: Studies measuring PTSD diagnostic accuracy among patients in specialized MH.

Group 2: Studies measuring PTSD diagnostic accuracy among patients in primary healthcare.

Group 3: Studies assessing clinicians’ ability to accurately diagnose PTSD based on varying background information about patients, and how diagnostic expectation biases influence diagnostic accuracy.

Two studies [23, 24] reported separate PTSD diagnostic accuracy outcomes by clinical setting, including primary care and specialized care. Therefore, they were included in both the primary and specialized care summary tables. This dual inclusion reflects stratified data, not duplication, and is footnoted accordingly in each table.

### Ethical considerations

Ethics approval and informed consent procedures were documented in all included studies, as detailed in Table S3. This systematic review and meta-analysis did not involve direct interaction with human participants or the collection of new data.

### Assessment of study quality

The sample participants in each study included either patients, clinicians, or both. In the original review, the overall sample comprised 37,032 patients and 424 healthcare professionals. The updated review added 8,236 patients and 0 clinicians, bringing the total sample across all studies included in the meta-analysis to 45,268 patients and 424 clinicians. These figures reflect participants who met all study-specific eligibility criteria and had complete data available, including the completion of all required assessments and accessible medical records necessary for effect size extraction. These totals include only participants from studies included in the meta-analysis; those from studies either transferred to the qualitative synthesis or newly added to it were excluded from these counts.

In the qualitative synthesis, newly included studies contributed an additional 8,280,456 patients and 273 clinicians to the overall sample. This total includes large administrative datasets and community-based surveys, as well as 129 clinicians from Ehlers et al. and 144 clinicians from Della Porta et al., both of whom focused on diagnostic practices.

The systematic review by Greene et al. [26], which was originally included in the meta-analysis of the previous review, was moved to the qualitative section due to its overlap with studies that were already independently included in this review. Specifically, 6 studies [29, 30, 31, 32, 33, 34] from Greene et al. [26] were already part of our meta-analysis, and 2 [35, 36] were included in our qualitative synthesis. After subtracting patient counts from overlapping studies, Greene et al. [26] contributed 19 unique studies, 18 of which met our baseline inclusion criteria, adding 21,227 non-overlapping patients to the qualitative synthesis.

Overall, the combined sample across the meta-analysis and qualitative synthesis included 8,346,951 patients and 697 clinicians.

The original meta-analysis included 11 cross-sectional diagnostic accuracy studies, 5 cross-sectional survey studies with diagnostic comparisons, 1 experimental vignette study, 1 cross-sectional study (not involving diagnostic comparison), 2 systematic reviews (one of which was subsequently moved to qualitative synthesis due to overlapping data), and 1 retrospective cohort study.

The updated meta-analysis included 7 cross-sectional diagnostic accuracy studies, 1cross-sectional survey study with diagnostic comparison, 1 prospective diagnostic validation study, 1 retrospective cohort study, and 1 retrospective survey with diagnostic comparison.

The qualitative synthesis includes the systematic review derived from the original meta-analysis, as well as 1 newly included systematic review. It also incorporates 1 experimental vignette study, 1 cross-sectional diagnostic validation study, 4 cross-sectional survey studies, 1 cross-sectional epidemiological study, 1 retrospective cohort study, 1 retrospective diagnostic coding analysis, 1 retrospective observational electronic medical records (EMR)-based study, 1 prospective comparative cohort study, 1 prospective naturalistic longitudinal study (reporting PTSD-related findings based on cross-sectional baseline data), 1 psychometric validation study, and 1 observational validation study.

With these additions, the total number of studies included across the original and updated reviews is 47. This update enhances the comprehensiveness of the review by incorporating additional cross-sectional and cohort studies, thereby providing a broader assessment of PTSD diagnostic accuracy across various healthcare settings. A detailed breakdown of the methods used to collect data from reports is presented in Table S1.

Software tools used for data collection included the EndNote reference management software [37] and Covidence [28]. The original review was conducted with the involvement of a secondary reviewer, who contributed to study selection and quality assessment. However, the updated review was conducted solely by the author, who applied the same rigorous methodology independently.

In the original review, quality assessments of the studies by da Silva et al. [38], Holowka et al. [39], Kostaras et al. [40], Lewis et al. [41], Reynolds et al. [42], van Zyl et al. [43], and Zammit et al. [25] were conducted independently by both the author and the secondary reviewer. Quality assessments for the remaining studies in the original review were conducted by the author under the supervision and revision of the secondary reviewer. A risk of bias assessment of the included observational and experimental studies was conducted using the Cochrane Risk of Bias Assessment Tool for Non-Randomized Studies of Interventions (ROBINS-I) [44]. In addition, a risk of bias assessment for each included systematic review was performed using the Grading of Recommendations, Assessment, Development, and Evaluations (GRADE) guidelines: 4. Rating the quality of evidence—study limitations (risk of bias) [45] and 5. Rating the quality of evidence—publication bias [46]. This was included in Table S2 after the original review was completed. The newly added studies were evaluated after the updated review, and risk of bias assessments were performed and added to Table S2. Documentation of Institutional Review Board approval for each included study was compiled by the author and added as Table S3 after the original review, with updates reflecting the newly included studies following the updated review. Although some included studies contained diagnostic accuracy components, only the secondary data describing real-world detection or recognition rates (i.e., the proportion of confirmed PTSD cases that were documented or identified in routine care) were extracted and analyzed.

As previously noted, several studies were included in the qualitative synthesis due to their relevance to PTSD underdiagnosis despite lacking the diagnostic accuracy metrics required for inclusion in the meta-analysis. These studies, while excluded from the quantitative synthesis and the corresponding participant totals, provide valuable contextual insight into the challenges of PTSD recognition across clinical settings. Their role is to supplement, rather than to statistically inform, the diagnostic accuracy analyses, thereby ensuring conceptual depth while maintaining methodological consistency.

### Data collection and abstraction

For the original review, two reviewers—the author and a secondary reviewer—participated in the systematic review, contributing to conceptualization, funding acquisition, data collection, and project administration during the initial stages of project development. The initial conceptualization of the project was led by the secondary reviewer. Preliminary data curation and formal analysis of the studies by da Silva et al. [38], Holowka et al. [39], Kostaras et al. [40], Lewis et al. [41], Reynolds et al. [42], van Zyl et al. [43], and Zammit et al. [25] were conducted by the secondary reviewer and were later extended to the author to be completed and expanded independently. The author conducted the investigation, methodology, project administration, validation, visualization, and writing. The software was obtained and independently managed by the author, while resource acquisition, including the screening of included and excluded studies, was primarily conducted by the author with support from the secondary reviewer. Partial funding was provided by the secondary reviewer, while the author contributed additional independent financial support. The selection process, title screening, and inclusion of studies in the systematic review were conducted independently by the author, followed by supervision and approval from the secondary reviewer regarding the appropriateness of the included studies, data extraction, and calculation of weighted means.

For the updated review, the systematic search and inclusion process were conducted solely by the author. The updated search was performed primarily using the consensus database, along with an extensive bibliographic search of previously included studies, newly added studies, and reference lists of studies identified through consensus but excluded during full-text screening. No additional reviewers participated in the updated review process. Study selection, full-text screening, and data extraction were conducted independently by the author, ensuring consistency with the original inclusion criteria.

The updated meta-analysis was performed entirely by the author. Weighted means of sensitivity — defined as the proportion of confirmed PTSD cases correctly identified by routine clinical diagnosis — were calculated along with standard errors and descriptive confidence intervals (CIs) were initially calculated in Excel (Microsoft Corp.) to verify the consistency of individual study estimates. For the pooled analyses, a random-effects meta-analysis of proportions was conducted using established meta-analytic procedures. The primary effect measure synthesized was sensitivity, representing the proportion of true PTSD cases correctly identified in routine clinical diagnosis. Further details on the statistical software, transformations, and visualization methods are provided in the Meta-analysis section of the Results. All calculations and visualizations were cross-validated for accuracy and consistency.

As with the original review, the risk of bias assessment for the new studies was conducted using the ROBINS-I [44] for both observational and experimental studies. The GRADE guidelines were used to assess the risk of bias, study limitations, and publication bias in the included systematic reviews. Any additional tools used for specific study types (e.g., for cross-sectional surveys or diagnostic accuracy studies) are described, and their citations provided, in S2. All additional assessments for the newly added studies were performed solely by the author.

Finally, studies mentioning the underdiagnosis of PTSD that lacked specific diagnostic accuracy statistics were included in the results and discussion sections but excluded from the meta-analysis. For studies reporting full diagnostic classification (including true positives, false positives, true negatives, and false negatives), sensitivity was calculated and used as the primary measure in the meta-analysis to maintain consistency across studies, as the majority of included studies reported only sensitivity. Overall diagnostic accuracy rates were calculated and reported in descriptive tables but were not included in pooled estimates. For studies reporting PPV as a proxy for diagnostic accuracy [23], results were presented separately in subgroup analyses and clearly labeled in all figures and tables. These studies were also not included in the final count of each study type or in the participant totals, as their focus was on identifying trends in PTSD recognition rather than reporting statistical accuracy rates.

The systematic review by Greene et al. [26], which has been included in the original review, was reassessed and excluded from the updated meta-analysis. Many of the primary studies cited in Greene et al. [29, 30, 31, 32, 33, 34, 35, 36] were independently located and included as separate entries during the updated review. To ensure scientific rigor and avoid data duplication, original studies were prioritized over aggregated data from Greene et al. [26]. While the Greene review is not part of the quantitative synthesis, it is retained for contextual discussion to support the broader narrative of PTSD underdiagnosis in primary care.

This comprehensive process ensures that the updated systematic review and meta-analysis reflect the most current research while maintaining methodological rigor and consistency.

## Results

In the original review, 21 studies met the inclusion criteria. As part of the updated review, 65 new studies were identified, and 57 additional studies were assessed in full-text after studies were discarded as duplicates, and were excluded following abstract screening. Of the 57 studies sought for full-text retrieval, 27 were excluded for not meeting eligibility requirements due to reasons such as a focus on populations already diagnosed with PTSD, the use of unrelated diagnostic tools, or a lack of required diagnostic accuracy data. Eleven studies were newly included in the meta-analysis, and 15 additional studies were added to the qualitative synthesis. Additionally, 1 study originally included in the meta-analysis was moved to the qualitative section after re-evaluation due to overlapping data or insufficient diagnostic accuracy metrics.

This brought the total number of included studies in the systematic review to 47: 31 in the meta-analysis and 16 in the qualitative synthesis. The updated PRISMA [21] flowchart (Fig. 1B) illustrates the study selection process, highlighting newly included studies and those excluded based on the revised eligibility criteria.

In the original search, 749 studies were screened, with 220 excluded as duplicates or irrelevant. After the abstract screening, 209 studies were excluded, leaving 320 full-text articles for eligibility assessment. Following the application of the inclusion criteria, 21 studies were included in the original review. The PRISMA flow diagram (Fig. 1A) illustrates this process [21].

For the updated review, an additional 53 studies were retrieved for full-text evaluation. Twenty-seven studies were excluded due to irrelevance, overlapping inclusion criteria, inappropriate methodology, or insufficient information for inclusion. Fifteen studies were retained as qualitative evidence of PTSD underdiagnosis, and 11 were included in the meta-analysis.

In total, 47 studies were included in both the original and updated reviews—31 in the meta-analysis and 16 in the qualitative synthesis. To minimize heterogeneity in the synthesis and meta-analysis, studies were grouped into 3 categories:

Group 1: Studies on PTSD diagnostic accuracy in specialized MH

Group 2: Studies on PTSD diagnostic accuracy in primary healthcare

Group 3: Studies on clinicians’ ability to diagnose PTSD accurately under various scenarios and the impact of diagnostic biases

This grouping strategy facilitated a coherent analysis while reducing the influence of methodological disparities. Table S1 summarizes the peer-reviewed published studies that meet our inclusion criteria, which are also described in the following paragraphs.

### Studies on PTSD diagnostic accuracy in specialized MH

Studies in this group reported PTSD diagnostic accuracy rates in environments staffed by trained MH professionals. These studies revealed a weighted mean diagnostic accuracy of 55.6%, reflecting slightly higher diagnostic reliability than in other settings.

Bonn-Miller et al. [47] conducted a cross-sectional diagnostic accuracy study to examine the accuracy of PTSD and other Axis I disorder diagnoses in the EMRs of US military veterans receiving care within the Veterans Health Administration. The study focused on a sample of 84 military veterans (96.4% male; mean age = 51.85 years) who met the criteria for cannabis use disorder and were recruited from MH clinics, including specialty substance use disorder (SUD) clinics, at a Veterans Affairs (VA) Medical Center.

PTSD and other Axis I disorders were assessed using structured clinical interviews: the Structured Clinical Interview for DSM-IV Non-Patient Version for most Axis I disorders [48], and the CAPS specifically for PTSD [49]. Diagnoses obtained from these structured interviews were compared with retrospective chart reviews of participants’ EMRs from VA MH clinics [50].

The results showed that 36.9% (31 out of 84) of participants met the DSM-IV criteria for PTSD based on the structured CAPS interview. However, only 21.4% (18 out of 84) had a PTSD diagnosis documented in their VA medical records. This yields a diagnostic accuracy of 58.1% (18/31), meaning that just over half of the true PTSD cases were recognized in routine clinical documentation.

The study also found similar patterns of underdiagnosis for cannabis and other anxiety disorders, while mood disorders and other substance use disorders were often overdiagnosed. The authors suggested that the underdiagnosis of PTSD may be due to a lack of structured assessments in routine care, delays between symptom onset and clinical visits, and clinicians focusing on the most prominent or disruptive symptoms (e.g., substance use). All structured interviews were administered by trained research staff, and diagnoses were confirmed through audio recording reviews by the lead author, enhancing diagnostic reliability.

The risk of bias was moderate. Strengths included the use of validated structured diagnostic instruments (DSM-based and CAPS), diagnostic confirmation by experienced clinicians, and comparison with real-world EMRs. However, limitations included the small and relatively homogeneous sample (primarily older male veterans with cannabis use disorder), potential selection bias from self-referral, and lack of inter-rater reliability reporting. Additionally, the generalizability of the findings to other veteran populations (e.g., female veterans, non-cannabis users) may be limited.

The findings highlight a significant gap in the recognition of PTSD among veterans with co-occurring substance use disorders and underscore the importance of implementing regular, structured assessments in VA MH services.

Cusack et al. [51] conducted a cross-sectional survey study with diagnostic comparison and examined trauma exposure and PTSD recognition among individuals with serious mental illness in a psychosocial rehabilitation program in the US The study involved 142 adult participants recruited from a community MH center, all of whom had a history of psychiatric hospitalization. PTSD symptoms were assessed using the Trauma Assessment for Adults Self-Report Version (TAA) and the PCL, and the findings were compared with documentation in clinical charts.

The study reported that 87.2% of participants had experienced at least one traumatic event. PTSD prevalence varied by scoring method: 19% of participants met the criteria for PTSD using a PCL cut-off of 50, 29.6% using a cut-off of 45, and 30.3% using a DSM-based method. Notably, only 3% of participants (5 out of 142) had a chart diagnosis of PTSD. This indicates that the diagnostic accuracy was only 11.6% when using the DSM-based method as the benchmark (5 out of 43 participants identified by the DSM method had a documented PTSD diagnosis). The authors attributed this underdiagnosis to limited clinician recognition, despite the MH center’s involvement in a state-wide trauma initiative.

Although the study used validated screening tools and attempted to address literacy issues by reading questions aloud, it relied on self-report data and did not confirm diagnoses through clinician-administered interviews (e.g., CAPS). These limitations may introduce some risk of bias, particularly regarding detection and diagnostic confirmation. Nevertheless, the study emphasized the under-recognition of PTSD in public MH systems and called for improved trauma assessment and training in these settings.

da Silva et al. [38] conducted a cross-sectional diagnostic accuracy study with 200 psychiatric outpatients at the Institute of Psychiatry of the Universidade Federal do Rio de Janeiro in Brazil. The mean age was 48 years (range 20–76), and 59% were women. Participants were assessed using the PTSD module of the SCID [48], and medical records were reviewed for diagnoses made by psychiatrists in training. The study found that 20.5% (41 participants) met the criteria for current PTSD; however, only 1 of them (2.4%) had been previously diagnosed, indicating a diagnostic accuracy of 2.4% and an underdiagnosis rate of 97.6%. The authors attributed this gap to the diagnostic complexity of PTSD, the overlaps of symptoms with other mental disorders, and the tendency of patients to underreport traumatic experiences due to shame or avoidance. The diagnostic procedure was iterative and supervised by clinical experts, although no formal inter-rater reliability was reported. Overall, the study demonstrated a moderate risk of bias, primarily due to potential selection and detection limitations. A detailed risk of bias assessment is provided in S2.

de Bont et al. [52] conducted a cross-sectional diagnostic accuracy study that examined adult patients (aged 18–65 years) with psychotic or mood disorders featuring psychotic symptoms in Dutch MH facilities. The study was conducted across 13 long-term MH organizations in the Netherlands. The researchers used CAPS to diagnose PTSD and the Mini-International Neuropsychiatric Interview-Plus to evaluate lifetime psychotic disorders.

Participants were initially screened using the Trauma Screening Questionnaire (TSQ), but PTSD diagnoses were based on CAPS [5]. Of the 2,608 participants screened, a subsample of 455 was interviewed using the CAPS and the Mini-International Neuropsychiatric Interview-Plus. Among these, 146 participants (32.1%) met the criteria for PTSD according to CAPS. Only 13 individuals (0.5%) had a PTSD diagnosis recorded in their charts. Of these, 12 cases (92.3%) were confirmed by CAPS, reflecting a high PPV among recognized cases. However, only 12 of the 146 CAPS-confirmed cases were documented in medical records, resulting in a diagnostic recognition rate of approximately 8.2%. This substantial under-recognition highlights a critical gap in PTSD detection within this clinical population. The TSQ demonstrated strong psychometric properties as a PTSD screening tool. At the optimal cut-off score of 6, it yielded a sensitivity of 78.8%, a specificity of 75.6%, and an area under the receiver operating characteristic curve of 0.85 (95% CIs 0.83–0.87), indicating good classification accuracy.

Holowka et al. [39] conducted a cross-sectional diagnostic accuracy study that investigated the diagnostic validity of PTSD in VA EMRs by comparing them with structured clinical interviews among Iraq and Afghanistan veterans participating in Project VALOR (Veterans After-Discharge Longitudinal Registry) [53] in the US A total of 1,649 participants underwent the PTSD module of the SCID, conducted by trained doctoral-level clinicians who were blinded to EMR status. Concurrently, researchers extracted PTSD diagnoses from the VA’s Problem List and Encounter data in the National Patient Care Database.

The results showed that 1,039 participants met the criteria for current PTSD as determined by the SCID. Using the Problem List as the EMR-based indicator, 1,175 participants were classified as having PTSD, while 474 were classified as non-cases. The diagnostic concordance between the EMR Problem List and SCID diagnoses for current PTSD was 73.2%, and 72.3% for encounter data. Based on the SCID as the reference standard, the Problem List yielded 886 true positives, 289 false positives, 153 false negatives, and 321 true negatives, resulting in a diagnostic accuracy of 73.2% for current PTSD using VA EMRs. The recognition rate—defined as the proportion of true PTSD cases correctly identified in EMRs—was 85.3% (886 out of 1,039). The authors highlighted concerns regarding the overreliance on informal clinical assessments in real-world settings, noting that 26.8% of EMR diagnoses either underrepresented or overrepresented true PTSD cases. They also identified factors associated with misclassification, such as lower combat exposure, fewer PTSD avoidance symptoms, reduced functional impairment, and a lower likelihood of panic disorder among false negatives. Conversely, false positives were more likely to have sought treatment for emotional problems and demonstrated lower overall symptom severity and impairment.

While structured interviews ensured rigorous case identification, the study acknowledged limitations, including the use of retrospective data, potential interviewer variance, and limited generalizability to veterans receiving VA care. Overall, the study emphasized the importance of structured assessments in improving PTSD diagnostic accuracy in routine clinical practice.

The study had a moderate risk of bias due to potential misclassification from retrospective EMR coding, reliance on a single structured interview, and limited generalizability to broader non-veteran populations.

Lommen and Restifo [54] conducted a cross-sectional survey study with diagnostic comparison that investigated the under-recognition of trauma and PTSD in 33 adult outpatients with schizophrenia (n = 23) or schizoaffective disorder (n = 10) at a psychiatric clinic in the Netherlands. Participants completed the Trauma History Questionnaire-Revised, the PTSD Symptom Scale–Self-Report, and the Post-Traumatic Cognitions Inventory (PTCI). According to DSM-IV criteria, 39.4% (13 participants) met the criteria for current PTSD using a liberal scoring method that excluded the A1 criterion and counted symptoms as present if rated at least 1 (i.e., “once a week or less”). When the A1 criterion (objective threat) was included, the prevalence decreased to 18.2%. Using a more conservative scoring rule (with symptoms rated at least 2) while excluding A1, 21.2% of participants met PTSD criteria, dropping to 9.1% when A1 was included. This most conservative scoring method (including A1 and symptoms rated ≥ 2) was used as the diagnostic standard in the current review. However, none of the participants with PTSD had a diagnosis documented in their medical charts, resulting in a diagnostic accuracy (recognition rate) of 0%. Although 69.7% of the worst trauma events reported by participants were recorded in their charts, the PTSD diagnosis itself was consistently missed. The authors suggested several possible reasons for this underdiagnosis, including clinicians’ focus on psychotic symptoms, patients’ reluctance to disclose trauma, and symptom overlap between schizophrenia and PTSD. The study also examined the relationship between trauma-related cognitions and PTSD symptom severity. Negative post-traumatic cognitions, as measured by the PTCI total score, were strongly associated with PTSD symptom severity (r = .74, p < .001). All 3 PTCI subscales (negative self-cognitions, negative worldviews, and self-blame) were positively correlated with PTSD symptoms; however, backward regression analysis indicated that only self-blame and self-related cognitions remained significant when controlling for overlap. The study concluded that PTSD is likely under-recognized in this population and emphasized the importance of incorporating trauma screening and cognitive PTSD models into assessments of individuals with psychotic disorders. The study had a moderate risk of bias, primarily due to its small sample size, reliance on clinician referrals, and the use of self-report instruments rather than structured interviews.

Marx et al. [55] conducted a cross-sectional diagnostic accuracy study that enrolled 764 veterans as part of Project VALOR in the US and assessed both current and lifetime PTSD using the SCID. These diagnoses were compared with evaluations conducted by compensation and pension (C&P) examiners and with VA PTSD service-connection status. The study reported a 70.4% diagnostic concordance rate between SCID and C&P diagnoses for current PTSD and 77.7% for lifetime PTSD. True positives (62.9%) were the most common, followed by false positives (16.4%), false negatives (13.1%), and true negatives (7.4%). The overall diagnostic accuracy was 70.4%. Veterans diagnosed by C&P examiners were over 3 times more likely to meet PTSD criteria according to the SCID (odds ratio [OR] = 3.39, 95% CI 2.25–5.15). The study also explored racial disparities, finding that Black veterans were significantly less likely than White veterans to receive PTSD diagnoses from C&P examiners when SCID-positive and were more likely to be false negatives, especially in the absence of psychometric testing. White veterans were more likely to be false positives (26.5% vs. 54.5%; OR = 4.07, p < .001). When psychometric testing was used, these racial disparities were no longer significant. Psychometric testing was included in only 24.2% of exams. The study had a low risk of bias, employing blinded interviewers, standardized diagnostic instruments, and noting potential confounders such as the use of psychometric tests and site-level variation.

Schwartz et al. [56] conducted a cross-sectional diagnostic accuracy study that examined 184 African American outpatients from an urban community MH clinic in the US over an 18-month period to assess the prevalence of trauma exposure and PTSD. PTSD symptoms were initially evaluated using the Perceived Stress Scale, which indicated that 43% (80/184) of participants met the symptom criteria for PTSD. A random subsample of 72 participants completed the Structured Clinical Interview for DSM-IV Axis I Disorders (SCID-I), administered by trained interviewers who were blinded to the survey responses. Among this subsample, 29 participants (40%) met the PTSD diagnostic criteria based on the SCID-I. To evaluate clinical recognition of PTSD, medical records were reviewed for 66 of the 72 participants who completed the SCID. Among the 26 individuals in this group with a SCID-based PTSD diagnosis, only 3 (11.5%) had a corresponding diagnosis recorded in their clinical records, indicating substantial under-recognition. The study emphasized the high prevalence of trauma and PTSD in this underserved population, with 83% of the total sample reporting at least one trauma that meets DSM-IV Criterion A for PTSD. Comorbid conditions were also common; PTSD-positive participants had significantly more SCID-diagnosed comorbidities, including major depression and nonschizophrenic psychotic disorders. While the use of validated assessment tools and structured interviews improved internal validity, limitations included single-site recruitment, retrospective chart review, and potential selection bias due to attrition, as well as reliance on a subsample for diagnostic confirmation.

Wang and Vivek [57] conducted a cross-sectional survey with a diagnostic comparison study among 62 psychiatric outpatients at 2 inner-city MH clinics in Queens, New York, US Participants completed the Traumatic Life Events Questionnaire and the PTSD Checklist–Civilian Version (PCL-C). Results from these instruments were then compared with participants’ clinical records to assess the under-recognition of PTSD.

The study found that 61% (n = 38) of participants screened positive for PTSD using the PCL-C (cut-off = 44), while only 13% (n = 8) had a PTSD diagnosis documented in their clinical charts (p < .01). This yields a diagnostic accuracy of approximately 21%, inferred as the proportion of diagnosed cases in medical records (n = 8) among those identified with PTSD by the PCL-C (n = 38). The study also found that 88% of participants reported experiencing traumatic life events in the survey, while only 22% had trauma noted in their medical records (p < .05), indicating that trauma exposure was frequently under-recorded. Additionally, participants who screened positive for PTSD were more likely to report major depressive disorder (MDD) and to be prescribed selective serotonin reuptake inhibitors (SSRIs), suggesting potential comorbidity or diagnostic overlap. However, these differences did not reach statistical significance due to the small sample size (MDD prevalence in the PTSD group = 37% vs. 13% in the non-PTSD group; p = .07).

The study did not use a gold-standard structured interview (e.g., CAPS), which limits the validity of the PTSD classifications. Additionally, the authors acknowledged this limitation, along with the possibility of selection bias from voluntary participation and diagnostic inaccuracies during routine clinical encounters. They concluded that PTSD and trauma exposure were substantially under-recognized in these outpatient psychiatric settings, reinforcing the need for more consistent screening and diagnostic evaluation.

Van Zyl et al. [43] conducted a cross-sectional diagnostic accuracy study at Neuroclinic C, a therapeutic inpatient unit at Stikland Hospital in Bellville, Cape Town, South Africa, specializing in mood and anxiety disorders. The researchers enrolled 40 (82.5% women; mean age = 35.65 years) consenting participants, all of whom had previously undergone structured intake interviews based on DSM-IV-TR criteria. For the purposes of the study, participants were further assessed using the CAPS to diagnose PTSD and other psychiatric conditions [43].

The study found that 16 participants (40%) met DSM-IV-TR criteria for current PTSD using CAPS; however, none had been diagnosed with PTSD at admission, resulting in a diagnostic accuracy of 0% within the study cohort. During the study period, 293 patients were admitted to the ward. Of these, 30 were diagnosed with PTSD at discharge, including the 16 identified through the research protocol. This implies that only 14 patients (4.8%) were diagnosed with PTSD through routine clinical care, reinforcing the study’s conclusion regarding the under-recognition of PTSD.

MDD was the most common admission diagnosis (44%) among participants who were later found to have PTSD. Cannabis use was also significantly associated with PTSD in the sample (p = .01), whereas alcohol and other substance use were not. Although the prevalence of PTSD was higher among women than men (45.5% vs. 14.3%), and slightly higher among mixed-ethnicity participants than others, these differences were not significant—likely due to the small sample size. The authors suggested that many PTSD cases are missed because clinicians do not routinely inquire about trauma exposure, and patients may be reluctant to disclose traumatic experiences without being prompted.

Overall, the study demonstrated a mix of low, moderate, and high risks of bias. Key concerns included the lack of blinding of participants and evaluators, as well as unclear reporting of allocation concealment and inclusion/exclusion procedures. Although CAPS ensured diagnostic rigor, the study’s small sample size and lack of blinding contributed to a moderate overall risk of bias.

Zammit et al. [25] conducted a systematic review and meta-analysis to estimate the prevalence of undetected PTSD in secondary-care MH services. The review included 29 peer-reviewed studies comprising 6,412 individuals with psychiatric diagnoses based on DSM or International Classification of Diseases (ICD) criteria. PTSD was assessed using validated self-report questionnaires or structured interviews, and diagnoses recorded in clinical notes were reviewed to identify recognition rates.

The study reported a median PTSD prevalence of 33.3% (interquartile range [IQR]: 23.4–40.0%) based on screening tools compared to 2.3% (IQR: 1.1–4.5%) based on clinical diagnoses. The median detection rate was 11.5% (IQR: 2.8–19.4%), indicating that only a small proportion of true PTSD cases was documented clinically. The median proportion of undetected PTSD was 28.6%, with substantial heterogeneity across studies (I² = 93.3%). This corresponds to a median diagnostic recognition rate of 11.5%, based on the proportion of true PTSD cases identified in clinical records. Notably, detection rates were lowest in US studies, inpatient samples, and among those with high proportions of individuals with psychotic disorders. There was no evidence that studies using structured interviews yielded significantly different detection rates compared to those using self-report questionnaires.

The authors acknowledged that self-report screening tools may slightly overestimate PTSD; however, all tools used were validated and demonstrated good specificity. Only a minority of studies masked assessors from clinical information or screening results, potentially introducing bias. Despite this, subgroup analyses revealed robust findings. Prevalence estimates were slightly lower in studies with a lower risk of bias, but it was emphasized that all tools used were validated and demonstrated good specificity. Few studies reported the masking of reviewers, which may have introduced bias. Nevertheless, the findings were robust and consistent across the subgroup analyses. The study concluded that PTSD is substantially underdiagnosed in secondary MH globally and called for clinical trials to evaluate the utility of routine PTSD screening protocols in specialized MH settings.

Reynolds et al. [42] conducted a cross-sectional diagnostic accuracy study at a specialist inpatient addiction services center in South West Thames to assess the prevalence of comorbid PTSD in a UK SUD clinical population. The study involved interviews with 52 patients and reviews of their medical case notes. Participants were assessed using validated instruments, including the Trauma History Questionnaire, the Posttraumatic Stress Disorder Symptom Scale – Interview version, and the Addiction Severity Index.

The results showed that 38.5% (20 patients) met DSM-IV criteria for current PTSD, and 51.9% met the criteria for lifetime PTSD. Despite these high rates, trauma history was referenced in only 42.5% of patient charts, and only 1 patient had a documented PTSD diagnosis and was referred for treatment, yielding a PTSD diagnostic accuracy of 5%. The authors noted that PTSD symptoms were often embedded within broader psychiatric presentations and were routinely overlooked in clinical documentation.

Patients with PTSD reported significantly more distress related to trauma memories, greater impairment in work, social life, and family responsibilities, and higher levels of psychiatric and medical issues on the Addiction Severity Index for lifetime PTSD, while only the medical composite score was significantly higher for current PTSD. They also showed significantly greater use of multiple substances before treatment and stronger associations between substance use and trauma-related coping.

The authors highlighted the diagnostic gap, suggesting that PTSD symptoms in this population may be masked by comorbid psychiatric conditions or go unrecognized due to inadequate trauma inquiry. The study had a moderate risk of bias due to its small sample size and reliance on retrospective chart reviews; however, its use of structured diagnostic tools and in-depth assessments provided valuable insights into the under-recognition of PTSD in inpatient addiction treatment settings.

Lewis et al. [41] conducted a cross-sectional survey study with diagnostic comparison in the UK involving 1,946 adults with diagnosed mental disorders recruited through the National Centre for Mental Health. Participants were screened for PTSD using the TSQ, and diagnostic status was based on self-reported clinician diagnoses. Of the sample, 438 participants (23%) screened positive for current PTSD, while only 247 (13%) reported receiving a clinician-diagnosed PTSD diagnosis. Among them, only 169 (9%) had both a positive TSQ screen and a self-reported diagnosis, indicating that 271 individuals (13.9%) had probable undetected PTSD.

The study reported that 13.9% of the sample screened positive for PTSD but did not have a clinical PTSD diagnosis, reflecting an inferred diagnostic accuracy of approximately 38.6% (169 diagnosed out of 438 screening positive). Rates of undetected PTSD were higher in individuals with trauma histories involving childhood abuse (23.3%), sexual assault (8.1%), and domestic violence (7.8%), and were particularly elevated in those with personality, psychotic, bipolar, and anxiety disorders. Undetected PTSD was significantly more common among women, individuals with lower socioeconomic status, and those who were younger at their first psychiatric contact.

Although the study did not use structured clinician-administered diagnostic interviews such as the CAPS, it utilized the validated TSQ anchored to specific DSM-5 qualifying traumatic events. Key limitations included reliance on self-reported clinician diagnoses, the absence of lifetime PTSD screening, and potential underreporting of trauma due to participant perceptions. The authors emphasized the clinical importance of routine PTSD screening in psychiatric settings to reduce under-recognition and improve treatment outcomes for this commonly missed but highly treatable condition.

Meltzer et al. [58] conducted a cross-sectional diagnostic accuracy study involving 592 adult primary care patients at an urban safety-net hospital in the US to examine the discrepancy between PTSD diagnosis and treatment in routine care. PTSD was assessed using the Composite International Diagnostic Interview (CIDI) Version 2.1, depressive symptoms were measured using the 9-item Patient Health Questionnaire, and symptom severity was evaluated with the PTSD Checklist – Civilian Version (PCL-C). Researchers also reviewed participants’ EMRs from the previous 12 months to assess MH diagnoses and treatments, including SSRI prescriptions and MH service utilization.

Among the 133 participants who met PTSD diagnostic criteria through the CIDI, 94 (71%) also met criteria for comorbid depression. Despite this, only 14 participants (11%) had PTSD documented in their EMRs. PTSD documentation rates were similarly low in both subgroups: 10% among those with PTSD alone and 11% among those with comorbid PTSD and depression, highlighting a widespread under-recognition irrespective of comorbidity status. The overall recognition rate of 11% reflects a weighted average of both subgroups (4 of 39 for PTSD-only and 10 of 94 for PTSD + depression). Overall, 49% of PTSD-positive patients received some form of MH treatment: 18% received SSRIs, 14% saw a MH professional, and 17% received both. Notably, 88% of those who received treatment had some form of MH diagnosis documented in their EMRs, most commonly depression (71%) and PTSD (18%).

The authors concluded that PTSD is often under-recognized in primary care, particularly among patients without comorbid depression, and that many received treatment “fortuitously” due to therapeutic overlap, especially when depression was documented instead of PTSD. This yielded a diagnostic accuracy of approximately 11.5%.

The study had a low to moderate risk of bias. Strengths included the use of validated diagnostic tools and systematic EMR reviews, while limitations involved reliance on self-report measures, the absence of information on treatment type or duration, and a cross-sectional design that limited causal inference.

Note: Although conducted in a primary care setting, this study is classified under mental health services due to its focus on patients actively engaged in mental health treatment, making it relevant to PTSD diagnostic accuracy rates within MH.

Kostaras et al. [40] conducted a cross-sectional diagnostic accuracy study that examined the comorbidity of MDD and PTSD in 101 outpatients at 2 psychiatric outpatient units of the University Hospital of Athens, Greece. PTSD was diagnosed using the Mini-International Neuropsychiatric Interview (MINI), and a traumatic history was assessed with the Life Events Checklist. The study found that 38.6% of participants met the criteria for lifetime MDD–PTSD comorbidity, and 26.7% had current comorbidity. Notably, all patients diagnosed with PTSD also met the criteria for MDD. However, only 28.2% (11 of 39) of patients with PTSD had a PTSD diagnosis documented in their medical records, implying a diagnostic accuracy of 28.2%. The average duration of PTSD was 16 years, and PTSD typically preceded the onset of MDD by an average of 6.2 years, with 79.5% of cases showing PTSD onset before or simultaneously with MDD. This suggests that under-recognition may result in delayed or ineffective treatment. The authors identified several contributing factors to underdiagnosis, including clinicians’ reluctance to inquire about trauma, the absence of routine PTSD symptom assessments, and patients’ avoidance in disclosing traumatic experiences. Risk factors associated with MDD–PTSD comorbidity included chronic depression (OR = 8.81), prolonged or repeated trauma, earlier onset of psychiatric symptoms, lower educational attainment, and reduced global functioning. Contrary to most epidemiological findings, male sex was also associated with higher odds of comorbidity, potentially due to selection bias in a treatment-seeking population. Childhood trauma was initially associated with comorbidity but lost significance when controlling for repeated trauma. The study concluded that undiagnosed PTSD may contribute to treatment-resistant or chronic depression and emphasized the importance of trauma screening in patients with depression. It reported a moderate risk of bias, primarily due to its retrospective design and relatively small sample size.

Gravely et al. [23] conducted a cross-sectional diagnostic validation study that evaluated the validity of PTSD diagnoses in the US VA administrative data by comparing them to scores from the PCL collected through a national postal survey of 4,777 veterans. The study used PPV as a measure of diagnostic accuracy, defined as the proportion of veterans with a PTSD diagnosis in administrative data who also scored ≥ 50 on the PCL, a validated self-report screening tool for PTSD. PPV was calculated as the number of true positives (diagnosed cases with PCL ≥ 50) divided by the total number of administrative PTSD diagnoses (true positives + false positives). The overall PPV for a single PTSD diagnosis was 74.8%, indicating that about three-quarters of those diagnosed in the system were likely true cases of PTSD. When at least 2 PTSD diagnoses were recorded within a 4-month period, the PPV increased to 81.8%, indicating improved diagnostic validity. However, this stricter algorithm reduced the sample size by approximately 40%, excluding a substantial portion of real-world cases with only 1 documented diagnosis.

Importantly, the study stratified the results by clinical setting to assess contextual variation in diagnostic accuracy. Of the total sample, 2,071 veterans (43%) were first diagnosed in primary care, where the PPV was 69.3%, suggesting that nearly 31% of diagnoses in this setting may not represent true PTSD. In contrast, among the 2,662 veterans diagnosed in specialized MH clinics, including 2,122 (44.4%) in non-PTSD MH clinics and 540 (11.3%) in PTSD specialty clinics, the PPVs were 79.9% and 77.6%, respectively. These findings indicate that PTSD diagnoses in specialized settings are more likely to reflect true PTSD cases than those made in primary care. Diagnostic accuracy was also lower among older veterans (aged ≥ 65 years), with a PPV of 61.4% based on a single diagnosis, improving to 76.1% when 2 diagnoses were required.

While the study’s design provided rich administrative and survey-based data, it only included veterans with at least 1 PTSD diagnosis in the VA system, which means it could not assess how many veterans with PTSD were never diagnosed (i.e., false negatives). Consequently, it was not possible to calculate sensitivity, specificity, or negative predictive value; therefore, PPV was the only diagnostic accuracy metric reported. This limitation means that the study does not capture underdiagnosis but rather focuses on the validity of existing diagnoses. The authors noted that the observed PPVs were consistent with adjusted estimates accounting for non-response, and the 95% CIs were considered accurate due to bootstrapping methods.

For the purposes of this review, PPV is treated as a valid proxy for real-world diagnostic accuracy, as it reflects how often PTSD diagnoses made during routine clinical encounters align with validated screening results. Diagnoses in this study were recorded during standard VA care, based on routine clinical assessments rather than research protocols, which increases the generalizability of the findings to real-world clinical settings. Although the use of ≥ 2 diagnoses improved diagnostic precision, the review relies on the PPV derived from a single PTSD diagnosis to best reflect how PTSD is typically identified and recorded in everyday practice. This approach aligns with the objective of the review, which is to assess the accuracy of diagnoses as they are commonly made, rather than under ideal or filtered conditions. The authors concluded that while the PPV of administrative PTSD diagnoses is moderate, implementing a requirement for at least 2 PTSD-coded encounters within a defined timeframe may improve diagnostic validity in both clinical and research contexts.

For consistency and accuracy within the current meta-analysis, the PPV values derived from a single PTSD diagnosis were used to estimate real-world diagnostic accuracy in both primary care and specialized care settings. Specifically, for the purpose of this review, the number of participants diagnosed with PTSD in each setting was multiplied by the corresponding PPV to estimate the number of true PTSD cases. This yielded approximately 1,435 true PTSD cases in primary care (2,071 × 0.693) and 2,086 in specialized care (2,662 × 0.784). These estimates were used to enable comparability with other studies reporting diagnostic accuracy based on confirmed PTSD cases.

Although the study is described in specialized MH settings, its stratified data are also used to estimate diagnostic accuracy in primary care. This is the only study in the review that contributes data to both settings and is therefore included in the analyses for each subgroup; however, it is described only once to avoid duplication. Although the study is robust in size and statistical handling, its inability to capture undiagnosed PTSD cases and reliance on self-reports introduces potential selection and measurement bias.

Magruder et al. [59] conducted a cross-sectional diagnostic accuracy study to examine patient-level factors influencing the detection of PTSD in VA primary care settings. The study sampled veterans from 4 southeastern US VA medical centers, ultimately including 819 participants who completed structured diagnostic assessments. PTSD diagnoses were determined using the CAPS, which was administered by telephone within 2 months of the participants’ primary care visits. Provider recognition of PTSD was defined by the presence of an ICD-9 PTSD diagnosis code (309.81) in the VA EMR within a 2-year window centered on the clinic visit.

Among the 819 participants, 98 individuals (12%) met DSM-IV criteria for current PTSD based on the CAPS. However, only 42 of these 98 individuals (43%) had a PTSD diagnosis documented in their medical records, resulting in a real-world diagnostic accuracy (recognition rate) of 43%. This suggests that 57% of true PTSD cases were not identified by providers in primary care. The study also identified several factors that are significantly associated with higher recognition rates. Veterans with musculoskeletal pain diagnoses (e.g., arthritis, back pain) were more likely to be identified as having PTSD (OR = 3.8, 95% CI 1.17–12.39). Higher odds of recognition were also observed among those aged 50–64, those with prior substance use disorders (OR = 9.91, 95% CI 1.12–87.55), and those who had served in a war zone (OR = 3.0, 95% CI 1.1–7.9).

Recognition was more likely among individuals whose PTSD symptom presentation was characterized by reexperiencing (Cluster B) and hyperarousal (Cluster D) symptoms, rather than by avoidance/numbing (Cluster C). Emotional impairment, as measured by the Short Form Health Survey role-emotional subscale, was also associated with higher rates of recognition. No significant associations were observed between recognition and race, sex, education, or employment status.

The study highlights the substantial under-recognition of PTSD in VA primary care during the pre-screening era and underscores the influence of symptom presentation and comorbid conditions on providers’ diagnostic behavior. The authors advocate for improved provider education and the use of structured PTSD screening to enhance recognition accuracy. The study’s moderate risk of bias primarily stems from its reliance on retrospective medical record reviews and the absence of data on provider-level or systemic factors. Nonetheless, it provides a robust, real-world estimate of the extent to which PTSD goes undetected in primary care, with a diagnostic accuracy rate of 43%.

Note: Although conducted in Veterans Affairs (VA) primary care clinics, this study is categorized under specialized mental health settings due to the integration of mental health services within these clinics. The VA’s Primary Care–Mental Health Integration (PC-MHI) program embeds mental health professionals into primary care teams to provide timely and coordinated care for conditions such as PTSD, depression, and anxiety [22]. This model facilitates immediate access to mental health support during primary care visits, reflecting a specialized approach to trauma-related mental health care typical of VA treatment settings.

Ivanov et al. [60] conducted a cross-sectional diagnostic accuracy study at the University Hospital Lausanne in Switzerland to evaluate the prevalence and diagnostic accuracy of PTSD in a psychiatric emergency setting. The researchers compared structured PTSD assessments using the PTSD module of the MINI with diagnoses recorded during routine clinical care.

A prospective sample of 403 patients aged 18 to 65 years was enrolled between December 2009 and February 2010. After excluding individuals for refusal, language barriers, or incomplete data, 316 individuals were screened using the French version of the MINI. Of these, 64 patients (20.3%) met the diagnostic criteria for current PTSD. In comparison, a chart review of a historical sample of 2,983 patients seen over a 1-year period in the same setting found that only 21 patients (0.75%) had a PTSD diagnosis documented in their clinical records.

Using the 20.3% MINI-based prevalence as the reference, approximately 605 PTSD cases were expected in the historical sample. With only 21 PTSD diagnoses documented, this corresponds to a diagnostic accuracy of 3.47%, indicating that over 96% of likely PTSD cases were missed in routine care. Even within the screened sample, only 37.5% (24 out of 64) of MINI-positive cases were documented in the medical charts.

PTSD prevalence was highest among refugees and undocumented individuals (50.0%), individuals from countries with recent war histories (47.1%), migrants (29.8%), and patients without professional income (25.0%). Prevalence also increased with psychiatric comorbidity, rising from 14.6% in patients with 1 co-occurring disorder to 44.4% in those with 4. No significant differences were observed based on age or sex.

A focus group of 8 psychiatric residents found that, while the MINI tool was perceived as clinically useful and easy to use, it was considered unsuitable for systematic application in psychiatric emergency settings due to time constraints, limited follow-up, and the potential disruption of the initial therapeutic relationship. Many residents continued to use elements of the tool in follow-up consultations.

The study’s moderate risk of bias stemmed from factors such as a lack of blinding in medical chart documentation, no inter-rater reliability assessment, and the exclusion of 87 patients from the prospective sample (21.6%). Additionally, PTSD diagnoses in the chart review were based on unstructured clinical evaluations, which increases the risk of detection bias.

The study concluded that PTSD is severely under-recognized in psychiatric emergency services, with standard diagnostic procedures capturing only 3.47% of probable PTSD cases. Targeted screening for high-risk groups and improved clinician training in trauma assessment are recommended.

Tiet et al. [61] conducted a prospective diagnostic validation study to examine the prevalence and diagnostic accuracy of PTSD in US VA outpatient treatment settings, specifically within SUD specialty clinics and general MH clinics. The study compared PTSD diagnoses obtained through a structured diagnostic interview with those recorded in patients’ EMRs, and evaluated the diagnostic performance of several brief PTSD screening tools.

A total of 400 veterans participated: 158 from SUD clinics and 242 from a general MH clinic. PTSD was diagnosed using the Computerized Diagnostic Interview Schedule for DSM-IV, which is a structured diagnostic tool. According to this reference standard, 36.7% (58 out of 158) of SUD patients and 52.1% (128 out of 242) of MH patients met the criteria for current PTSD.

Medical records were reviewed to determine whether PTSD had been documented by clinicians in the 9 months before or after study participation. In the SUD sample, only 28 of the 58 PTSD-positive individuals had a diagnosis recorded in the EMR, resulting in a diagnostic accuracy of 48.3%. In the MH sample, 77 of the 128 PTSD-positive individuals were diagnosed during routine care, yielding a diagnostic accuracy of 60.2%. When combining both samples, 105 out of 186 PTSD cases were recognized clinically, producing a weighted diagnostic accuracy of 56.5%. This indicates that nearly 44% of PTSD cases identified by structured assessment were missed during usual care.

The study also evaluated several screening tools. In SUD clinics, a 4-item PCL variant demonstrated the highest efficiency (81%) at a cut-point of 8, with a sensitivity of 71% and a specificity of 88%. The full PCL achieved 79% efficiency at cut points 33–34. The Primary Care PTSD Screen (PC-PTSD) had a sensitivity of 79% and a specificity of 67% at a cut-point of 3, with an overall efficiency of 72%. In MH clinics, the screening tools also performed well. The PC-PTSD achieved 76% efficiency (sensitivity = 70%, specificity = 82%), while the while the 4-item and 2-item PCL variants showed equal efficiencies of 74%. Notably, the 2-item PCL variant, despite having only 2 items, showed a sensitivity of 82% and a specificity of 65% in the MH sample.

The risk of bias was considered to be moderate. Strengths included the use of a validated structured interview, real-world samples from multiple VA sites, and a comprehensive chart review. Limitations included the use of non-clinician interviewers, the possibility of over-reporting of symptoms due to a lack of clinical consequences, and differing time frames between screening instruments (30 days) and the diagnostic interview (12 months). Additionally, screening instruments were administered after the diagnostic interview, which diverges from standard clinical practice.

The authors concluded that PTSD is common but significantly underdiagnosed in VA outpatient settings. Brief validated screening tools, particularly the -item and 2-item variants of the PCL, can improve detection in both SUD and MH clinics. Routine PTSD screening is recommended to enhance diagnostic accuracy and ensure appropriate access to care.

### Studies on PTSD diagnostic accuracy in primary healthcare

Studies in this group reported PTSD diagnostic accuracy rates within primary care settings, where clinicians often lack specialized training in MH. These studies revealed a weighted mean diagnostic accuracy of 48.2%, reflecting a lower diagnostic reliability than that observed in specialized MH environments.

Bohnert et al. [24] conducted a retrospective cohort study using administrative data to examine whether same-day integrated MH services improved PTSD diagnosis and treatment initiation among veterans who screened positive for PTSD in primary care. Using administrative data from a nationally representative US Veterans Health Administration sample, the study included 21,427 primary care participants who screened positive on the Primary Care PTSD Screen (PC-PTSD) and had no prior PTSD diagnosis or MH treatment in the preceding 12 months. Participants were categorized by the type of care received on the day of screening: (1) primary care only, (2) same-day primary care–MH integration (PC-MHI), or (3) same-day specialty MH.

PTSD diagnostic accuracy is defined as the proportion of screen-positive individuals who received a formal PTSD diagnosis (ICD-9) in their medical records within 1 year of screening, varying by setting. Among the 18,157 individuals who received only primary care, 8,705 (47.9%) were diagnosed with PTSD. Among the 1,507 individuals seen by integrated PC-MHI services, 908 (60.3%) received a diagnosis, while 1,199 of the 1,763 individuals (68.0%) who received same-day specialty MH were diagnosed. Multivariable analyses showed that, compared to primary care only, the odds of receiving a PTSD diagnosis within 1 year were significantly higher for those seen by PC-MHI services (adjusted odds ratio [aOR] = 1.67) and specialty MH services (aOR = 2.03). These findings indicate that same-day access to MH services significantly improved diagnostic accuracy compared to primary care alone.

The study also evaluated treatment initiation among the 5,966 individuals who were diagnosed on the same day as their screening. Both PC-MHI and specialty care were associated with increased odds of initiating PTSD treatment within 12 weeks, including psychotherapy, antidepressants, or referral to a PTSD clinic. “Any treatment” was defined as the receipt of at least 1 of the following: psychotherapy, a PTSD clinic visit, or an antidepressant prescription. Compared to primary care alone, the odds of initiating any PTSD treatment within 12 weeks were significantly higher for those receiving PC-MHI services (aOR = 3.39) and specialty MH (aOR = 4.45). Although treatment outcomes were largely similar across both MH service types, antidepressant use was slightly but significantly more common among those referred to specialty MH (53.0%) compared to those referred to PC-MHI (52.4%, p < .05). Although it did not employ structured diagnostic interviews such as the CAPS, the study provides robust real-world evidence of PTSD recognition using routinely collected administrative data from a large, nationally representative veteran population. Diagnostic rates across all care settings were based on diagnoses made at any point within 1 year following the initial positive PTSD screen, rather than solely on those recorded at the point of care. Limitations include the use of administrative data, which may not reflect clinical severity, and treatment measures that capture only initiation, rather than completion or effectiveness. Additionally, the diagnosis and treatment codes were not always specific to PTSD. The use of administrative data and the lack of structured diagnostic assessments introduce a moderate risk of bias, particularly due to potential misclassification and the absence of clinical severity data.

Although discussed in the primary care section, this study is included in both the primary care and specialized MH meta-analyses, as it provides stratified diagnostic accuracy rates for each setting. Consistent with the approach used by Gravely et al. [23], the stratified data are incorporated into both pooled estimates without duplicating participants.

PTSD diagnosis counts for this study were calculated by multiplying the number of screen-positive participants in each setting by the cumulative incidence rates of the study. For primary care only, 18,157 × 47.9% = 8,703 diagnosed; for PC-MHI, 1,507 × 60.3% = 908 diagnosed; for specialty MH, 1,763 × 68.0% = 1,198 diagnosed; for combined MH settings, 908 + 1,198 = 2,106 diagnosed, yielding a combined diagnostic accuracy of 64.4% (2,106 / 3,270).

Meltzer-Brody et al. [62] conducted a cross-sectional diagnostic accuracy study at a university-affiliated gynecology clinic in the US to assess the prevalence of PTSD and diagnostic recognition among trauma-exposed women in a primary care setting. Of the 292 patients screened, 88 reported a history of trauma and were eligible for further evaluation. Among these, 32 completed structured diagnostic interviews using the MINI, and 25 individuals (78%) met DSM-IV criteria for current PTSD. However, only 3 of the 25 PTSD-positive women (12%) were receiving psychiatric care at the time of assessment, indicating a significant treatment gap. As the authors noted, “nearly all of these patients (n = 22 [88%]) with a diagnosis of PTSD were not receiving any kind of psychiatric treatment for PTSD or any other psychiatric disorder” [62].

The study also evaluated the Startle, Physiological Arousal, Anger, and Numbness, a brief 4-item PTSD screening tool, using the MINI as a reference standard. At a cut-off score of 5, the Startle, Physiological Arousal, Anger, and Numbness demonstrated a sensitivity of 72%, a specificity of 71%, and a PPV of 90%. The majority of PTSD-positive individuals exhibited psychiatric comorbidities, including major depression (62%), panic disorder (28%), social phobia (28%), generalized anxiety disorder (19%), and substance abuse (15%). These findings suggest that even in high-contact care environments, PTSD and related conditions may remain undetected unless actively screened for.

This study is included in the primary care meta-analysis due to its use of a validated screening tool and a structured clinical interview in a non-specialist medical setting. However, it presents a moderate risk of bias, primarily due to its small, trauma-selected sample, absence of blinding, and use of a single-site convenience sample. These factors limit generalizability to broader populations but still offer valuable insights into PTSD under-recognition in women’s primary care.

Note: The PTSD accuracy rate obtained from this study is estimated based on the proportion of PTSD-positive women receiving psychiatric care at the time of assessment. While this does not confirm a formal PTSD diagnosis or treatment specifically for PTSD, the study provides quantifiable diagnostic recognition data in a real-world primary care setting. Although this measure is less precise than diagnostic accuracy figures derived from direct comparisons with structured clinical interviews, the study was included in the meta-analysis because, despite its limitations, it contributes valuable quantitative evidence reflecting real-world diagnostic challenges. Additionally, its relatively small sample size limits its weight in the pooled estimates, and the statistical methods used in the meta-analysis account for between-study variability, minimizing the risk of undue influence while enhancing the overall generalizability of the findings.

Prins et al. [29] conducted a cross-sectional diagnostic accuracy study that examined the recognition of PTSD in real-world VA primary care by comparing PTSD diagnoses documented in veterans’ medical records with those established using CAPS, the gold-standard for PTSD assessment. The study included 188 veterans recruited from general and women’s primary care clinics at the VA Palo Alto and Menlo Park facilities in California. Among all participants, 46 (24.5%) met the diagnostic criteria for PTSD based on the CAPS. However, in a subset of 134 participants with available VA medical records, only 39% of CAPS-confirmed PTSD cases had a PTSD diagnosis documented in their charts, indicating that 61% were missed. This corresponds to a real-world diagnostic accuracy of 39% for routine clinical recognition of PTSD.

The study also evaluated the PC-PTSD. At the standard cut-off score of 3, it demonstrated a sensitivity of 78%, specificity of 87%, PPV of 65%, negative predictive value of 92%, and overall diagnostic accuracy (efficiency) of 85%. Its correlation with CAPS diagnoses was strong (r = 0.83).

When comparing tools, the PC-PTSD outperformed the PCL in all diagnostic metrics—sensitivity (78% vs. 46%), specificity (87% vs. 79%), and efficiency (85% vs. 71%)—though a corrigendum later corrected these numbers for the PCL, showing it actually had higher accuracy at an optimally efficient cut-off of 48 (sensitivity 84%, specificity 90%, efficiency 89%). Nonetheless, the PC-PTSD was noted to be preferable in primary care due to its brevity and ease of use.

Performance varied by sex: among men, the PC-PTSD exhibited 94% sensitivity and 92% specificity, while among women, it demonstrated 70% sensitivity and 84% specificity.

The authors concluded that PTSD is often under-recognized in routine VA primary care and that structured screening tools, such as the PC-PTSD, substantially improve diagnostic accuracy. They recommend its use as a first-stage screening tool to better identify veterans needing further evaluation and care.

The study was assessed as having a moderate risk of bias, primarily due to sample attrition (only 56% completed both study phases), limited availability of medical records (only 134 participants had chart data available for analysis), potential volunteer bias, and the single-site design, which may limit generalizability.

Note: This study is classified under primary care settings due to its focus on general and women’s primary care clinics within the Veterans Affairs system. It evaluates PTSD recognition using a brief screening tool (PC-PTSD) specifically developed for use in non-specialized, fast-paced primary care environments, reflecting the real-world challenges of PTSD detection in general medical settings.

Taubman-Ben-Ari et al. [30] conducted a national cross-sectional survey study with diagnostic comparison in Israel to examine the prevalence of PTSD and the diagnostic accuracy of PTSD identification by primary care physicians. The sample included 2,975 adult patients recruited from 26 randomly selected primary care clinics operated by Israel’s largest national health provider. Participants completed a series of validated self-report instruments, including a Trauma History Questionnaire, the PTSD Inventory (based on DSM-III criteria), and the 28-item General Health Questionnaire to assess psychological distress. Physicians who were blinded to patient responses independently completed a clinical assessment form indicating whether they believed the patient exhibited psychological distress or PTSD.

Among the total sample, 684 patients (23%) reported exposure to a traumatic event, and 247 of these individuals (36%) met the diagnostic threshold for PTSD based on the PTSD Inventory. This corresponds to a PTSD prevalence of 8.3% in the overall population. Prevalence differed significantly by sex, with 10.1% of women and 7.3% of men meeting PTSD criteria (p < .01). However, only 6 of the 247 PTSD-positive individuals (2.4%) were identified as having PTSD by their primary care physician, indicating a markedly low recognition rate. In addition, 8 individuals who did not meet the criteria for PTSD were incorrectly diagnosed with the disorder, resulting in 8 false positives.

Using these data, 2 key diagnostic accuracy metrics were calculated. First, the recognition rate (sensitivity), defined as the proportion of true PTSD cases correctly identified, was 6 / 247 = 2.4%. Second, the PPV, defined as the proportion of individuals diagnosed with PTSD who actually met the diagnostic criteria, was 6 / 14 = 42.9%. Although an overall diagnostic accuracy of 91.6% can be calculated using the formula (TP + TN) / Total = (6 + 2,720) / 2,975, this figure is heavily influenced by the large number of individuals without PTSD who were not diagnosed correctly. Thus, it provides limited insight into the challenges of accurately identifying PTSD in clinical practice.

In this review, the primary diagnostic accuracy metric used is the recognition rate (2.4%), as it best reflects the frequency with which PTSD cases are identified in real-world primary care, consistent with the review’s overarching aim. The PPV (42.9%) is also reported to contextualize the probability that PTSD diagnoses made by physicians are accurate, facilitating comparison with studies such as Gravely et al. [23], which used administrative data to assess diagnostic validity. However, the overall diagnostic accuracy metric is not included in the meta-analysis due to its limited interpretive value in imbalanced clinical populations.

The study had a moderate to high risk of bias, primarily due to its reliance on self-report diagnostic instruments rather than structured clinical interviews, the use of DSM-III criteria, a lack of comorbidity data, and the absence of a standardized reference interview. Additional limitations included the possible underreporting of trauma exposure, as the PTSD Inventory relied on unaided recall without probes for specific events and did not account for whether participants experienced multiple traumas or focused on the most impactful one. Nevertheless, its strengths included a large, nationally representative sample, blinded physician assessments, and strong real-world clinical relevance within a primary care context. The findings highlight both the under-recognition of PTSD and the limited diagnostic precision when PTSD is identified, reinforcing the need for improved screening and diagnostic strategies in general medical practice.

Graves et al. [31] conducted a cross-sectional diagnostic accuracy study that examined the prevalence, diagnosis, and treatment of PTSD among African American adults in urban primary care settings in the US The study recruited 738 participants from 4 academically affiliated primary care clinics, of whom 375 completed structured diagnostic interviews using the CAPS and the SCID. Among these 375 individuals, 24.3% (n = 91) met the diagnostic criteria for current PTSD.

Only 30.8% (n = 28) of these PTSD-positive individuals were recognized or treated within the healthcare system, as indicted by provider documentation or evidence of MH treatment (e.g., psychotropic medication or psychotherapy). This corresponds to an estimated PTSD diagnostic accuracy of 30.8%, calculated as the proportion of clinically confirmed PTSD cases that were identified or treated in real-world care. Importantly, the total sample used for this diagnostic accuracy estimate comprised the 375 individuals who completed structured clinical interviews (SCID and CAPS) rather than the full recruited sample of 738. This approach aligns with the diagnostic accuracy methodology used in studies employing structured diagnostic tools.

The study further found that 69.2% (n = 63) of PTSD-positive individuals had never received any form of MH treatment, and 47.8% (22 out of a subsample of 46) had never disclosed trauma-related symptoms to their healthcare providers. Among the 91 individuals diagnosed with PTSD, 29 (31.8%) were prescribed psychotropic medications, 17 (18.6%) were receiving psychotherapy, and 81.4% were not engaged in any form of psychotherapy at the time of the study.

Comorbidity was prevalent among those with PTSD: 23% had MDD, 22% had substance use disorders, and 5.5% had bipolar disorder. In total, 46.2% of individuals with PTSD had at least 1 comorbid psychiatric diagnosis. Many individuals also experienced significant social stressors, including housing instability and homelessness.

The authors attributed the underdiagnosis to multiple factors, including patients presenting with physical rather than psychological symptoms, stigma inhibiting trauma disclosure, limited clinician training in trauma-informed care, and systemic gaps in MH integration. They emphasized that more than half of PTSD-positive individuals had disclosed trauma or psychiatric symptoms to their physicians yet remained untreated, suggesting that provider awareness alone was insufficient to ensure adequate care. They advocated for the routine implementation of PTSD screening and collaborative MH models in primary care to improve diagnosis and access to treatment among underserved populations. The small sample size, urban setting, and use of a convenience sample introduce potential bias and limit the generalizability of the findings to broader populations.

Liebschutz et al. [32] conducted a cross-sectional diagnostic accuracy study at a hospital-based primary care internal medicine clinic at Boston Medical Center in Massachusetts, US, to evaluate the prevalence of PTSD and its recognition in routine clinical care. The study recruited 636 English-speaking adult patients, of whom 609 completed a screening interview, and 597 were ultimately eligible for diagnostic analysis based on having complete CIDI data.

PTSD was assessed using the CIDI, a structured diagnostic instrument aligned with DSM-IV criteria. Of the 597 individuals, 117 (19.6%) met the criteria for current PTSD. Researchers then reviewed EMRs for each of these 117 PTSD-positive individuals to determine whether PTSD had been documented by their healthcare providers.

Among those with a research-based PTSD diagnosis, only 13 individuals (11.1%) had any mention of PTSD in their EMRs, whether as a listed diagnosis, noted in the medical narrative, or flagged using PTSD-specific ICD-9 codes. This corresponds to a PTSD diagnostic accuracy of 11.1%, defined as the proportion of clinically confirmed PTSD cases that were recognized and documented by healthcare providers.

The study also found that 51% of PTSD-positive individuals had depression noted in their EMRs, suggesting that clinicians may have recognized psychiatric symptoms but misattributed them to other MH conditions, particularly depression. Other common comorbidities, such as chronic pain and anxiety, were also prevalent and may have contributed to diagnostic overshadowing. In adjusted analyses, current PTSD was significantly more common among participants with chronic pain (23% vs. 12%), anxiety disorders (42% vs. 14%), irritable bowel syndrome (34% vs. 18%), and major depression (35% vs. 11%). PTSD prevalence was lower among immigrants compared to U.S.-born participants (13% vs. 21%, p = .046), a finding the authors attributed to the study’s English-language requirement and possible “healthy immigrant effect.”

Importantly, this study emphasized the under-recognition of PTSD even in a safety-net primary care setting, where individuals frequently present with complex psychosocial and medical histories. The authors advocate for routine trauma screening and enhanced clinician training to address the high rate of missed PTSD diagnoses.

For the purpose of this meta-analysis, only the 597 participants who completed the full diagnostic assessment (CIDI) were included, as they provided the complete data required to calculate PTSD prevalence and recognition rates. This approach aligns with the methodological framework of the original study. The study’s reliance on a single-site, English-speaking sample and self-reported data may limit generalizability and introduce selection bias.

Lu et al. [63] conducted a retrospective survey study with diagnostic comparison in an urban primary care setting (the Infectious Diseases Unit at a Rutgers-affiliated clinic in New Jersey, US) to assess the prevalence of PTSD symptoms and associated health factors among individuals living with HIV. The study included 135 primarily African American individuals who completed the PC-PTSD for DSM-5 (PC-PTSD-5) between February 2019 and September 2020. Retrospective chart reviews were conducted between May 2020 and April 2022 to assess PTSD documentation in EMRs. Inclusion criteria required a documented HIV diagnosis and completion of the PC-PTSD-5 screening. Of the 135 individuals, 49.6% (n = 67) screened positive for probable PTSD using a cut-off score of 3; however, only 11.9% (n = 16) had PTSD documented in their EMRs. This corresponds to a diagnostic accuracy of 23.9% (16/67). The authors interpreted this as evidence of significant underdiagnosis, consistent with past literature on low PTSD detection rates in HIV care settings.

PTSD symptoms were moderately correlated with depression (r = 0.40, p < .001), alcohol use (r = 0.28, p = .003), insomnia (r = 0.21, p = .022), and anxiety (r = 0.36, p < .001). They were not significantly correlated with drug use, pain, or any HIV-related laboratory results. Individuals with both PTSD and MDD experienced significantly worse outcomes, including increased homelessness (22.2% vs. 5.1%, p < .05) and a higher number of medical comorbidities (M = 11.67 vs. M = 9.17, p < .05). Those with PTSD and MDD also had significantly worse scores in depression, anxiety, and insomnia. The study underscores a substantial gap between PTSD symptom prevalence and clinical recognition in primary care, and it supports the integration of trauma-informed care using validated screening tools, such as the PC-PTSD-5. It is one of the first studies to evaluate the implementation of the PC-PTSD-5 in an HIV-positive, urban, predominantly African American population, and it supports the feasibility and utility of screening in this high-risk group.

The primary limitation of this study is the reliance on a screening tool (PC-PTSD-5) rather than a structured clinical interview, such as the CAPS or SCID, to establish probable PTSD, which limits diagnostic precision. Additionally, the small sample size (n = 135) and the retrospective chart review may introduce selection and documentation bias. The study’s generalizability is further constrained by its single-site design and demographic homogeneity, as it primarily involves African American patients in an HIV clinic.

Carey et al. [33] conducted a cross-sectional diagnostic accuracy study at a primary healthcare clinic in a South African township to assess trauma exposure, PTSD prevalence, psychiatric comorbidities, functional impairment, and diagnostic recognition. Among 201 adult participants, 94% reported exposure to traumatic events (mean = 3.8 lifetime trauma exposures). The prevalence of current PTSD was 19.9%, with 40 individuals meeting the diagnostic criteria. PTSD was significantly associated with poverty and single status, and it showed strong comorbidity with major depression (75%), somatization (35%), and panic disorder (25%; all *p* < .01). Both sexes were equally likely to develop PTSD. Functional impairment was significantly greater among individuals with PTSD and comorbid depression (*p* = .04). Despite this high burden, none of the 40 PTSD-positive individuals had previously received a PTSD diagnosis (0% diagnostic accuracy), and only 1% of the total sample had been prescribed psychotropic medication, indicating a near-total lack of PTSD recognition and treatment in this primary care setting. Neither PTSD nor trauma exposure was associated with increased service use or dissatisfaction with care, and clinicians failed to identify any cases of trauma or psychopathology in the medical records. The study had a moderate risk of bias due to its single-site design, reliance on retrospective chart review for diagnostic recognition, and lack of blinding between the diagnostic assessment and medical record review.

Seal et al. [64] conducted a retrospective cohort study to examine the identification and follow-up of PTSD in routine clinical settings among post-9/11 US veterans returning from deployment. The study analyzed the medical records of 750 veterans from Operation Iraqi Freedom and Operation Enduring Freedom who received care at VA primary care or community-based outpatient clinics. As part of a post-deployment MH screening program, 338 veterans (45%) completed a structured assessment that included the PC-PTSD and the 2-item Patient Health Questionnaire for depression. High-risk alcohol use was also assessed using the Alcohol Use Disorders Identification Test – Consumption (AUDIT-C), which the authors described as a 4-item version, although the standard AUDIT-C includes 3 items.

Among those screened, 171 veterans (50.6%) tested positive for PTSD, indicating a high prevalence of trauma-related symptoms. However, only 24% of screen-positive veterans had a completed VA MH appointment within 90 days of screening, and 46% had appointments scheduled. Over the entire study period, 73% of screen-positive veterans were eventually seen in VA MH services. In contrast, only 8% of screen-negative veterans received follow-up care, suggesting that screening status strongly influenced referral patterns.

While the study did not use structured clinical interviews (e.g., CAPS or SCID) to determine diagnostic accuracy in the strict sense, it provides important real-world data on PTSD recognition and care engagement in primary care. The findings highlight a gap between symptom detection and timely MH referral, particularly since nearly one-third of screen-positive veterans did not receive any follow-up.

The study demonstrated a moderate risk of bias. Strengths included the use of validated screening tools, the analysis of system-level referral outcomes, and real-world VA clinical settings. Limitations include the single-site design, the absence of a diagnostic gold-standard, reliance on chart data, and a lack of information on whether PTSD was formally diagnosed following referral. Additionally, the study lacked data on non-VA MH treatment, which could have influenced follow-up rates. Nonetheless, the study provides valuable insights into the recognition and early management of PTSD among recently deployed veterans in routine care settings.

Note: The diagnostic accuracy rate obtained from this study is based on follow-up MH clinic attendance after positive PTSD screening rather than a gold-standard structured diagnostic interview. While this measure does not definitively confirm PTSD diagnosis, it offers a useful estimate of clinical recognition and linkage to care. The reliance on administrative data and absence of formal diagnostic confirmation limits the reliability of this measure, potentially resulting in under or overestimation of true PTSD recognition. Despite these limitations, the study highlights significant under-recognition and gaps in timely referral for PTSD within routine primary care settings. It was included in the meta-analysis because it provides quantitative data on real-world clinical recognition and treatment engagement, broadening the evidence base and capturing pragmatic aspects of PTSD diagnosis beyond structured interviews. Although methodologically distinct, its inclusion adds valuable insight into PTSD recognition in clinical practice and is unlikely to unduly bias pooled estimates due to its indirect measure and the meta-analytic methods used.

Kimerling et al. [34] conducted a cross-sectional diagnostic accuracy study at VA primary care clinics in the US to assess the diagnostic accuracy of PTSD and the performance of a brief screening instrument. A total of 134 veterans (66% women) were recruited from general internal medicine and women’s health outpatient clinics. All participants completed the Breslau 7-item PTSD screen and a structured diagnostic interview using the CAPS, which served as the reference standard. Based on the CAPS, 34 participants (25.4%) met the criteria for current PTSD.

Medical records were reviewed to assess clinical recognition of PTSD. Among the 34 individuals with CAPS-confirmed PTSD, 13 (38.2%) had a documented PTSD diagnosis in their VA EMRs, indicating a diagnostic accuracy of 38.2%. The authors interpreted this as evidence of substantial under-recognition of PTSD in primary care, even within a large integrated healthcare system. The Breslau screen demonstrated strong performance, with a sensitivity of 85% and a specificity of 84% at the recommended cut-off of 4, and a higher specificity of 95% (but lower sensitivity of 67%) at a stricter cut-off of 6.

The study highlights the potential utility of brief self-report tools for identifying PTSD in time-limited primary care environments and supports the routine use of validated screeners to improve recognition. It also emphasizes the role of provider discomfort, competing medical demands, and lack of structured screening as contributors to underdiagnosis in routine care. The study is included in the primary care meta-analysis due to its structured diagnostic reference and the use of real-world EMR data to assess PTSD documentation. Risk of bias was moderate due to the relatively small sample size and single-site VA setting; however, the methodology was rigorous and directly aligned with diagnostic accuracy evaluation.

### Studies on clinicians’ ability to diagnose PTSD accurately under various scenarios and the impact of diagnostic biases

Studies in this group evaluated clinicians’ PTSD-related knowledge and their susceptibility to diagnostic biases under controlled conditions. McKenzie and Smith [65] assessed clinicians’ factual understanding of PTSD symptomatology, reporting a correct knowledge rate of 74%. McGuire et al. [66] using an experimental vignette design, examined the influence of contextual bias (e.g., foster care vs. biological mother) on PTSD diagnosis and found that only 22.7% of clinicians selected PTSD as the primary diagnosis despite identical symptom presentations. Based on these studies, the weighted mean performance estimate reflecting either PTSD knowledge or diagnostic consistency in the presence of bias was 41.33%, underscoring notable limitations in both foundational knowledge and bias mitigation in clinical reasoning.

McGuire et al. [66] conducted an experimental vignette study involving 270 UK MH professionals who completed an online survey. Participants were randomized to one of two vignettes describing a teenage boy presenting with identical PTSD symptoms, one living in foster care and the other with his biological mother. They were asked to select a primary diagnosis and recommend appropriate treatment approaches. Across the full sample, 22.7% of participants selected PTSD as the primary diagnosis. The difference in diagnosis rates between the two vignettes was significant (χ² (1) = 10.09, p = .001, OR = 2.65), with clinicians being more likely to assign PTSD as the primary diagnosis in the biological mother vignette (31.0%) compared to the foster care vignette (14.5%). For the biological mother vignette, the most frequently selected diagnoses were developmental trauma (51.2%), PTSD (31.0%), and attachment problems (7.0%). In contrast, for the foster care vignette, clinicians most commonly selected developmental trauma (57.3%), followed by attachment problems (22.1%) and PTSD (14.5%). Overall, only 33.9% of participants selected a National Institute for Health and Care Excellence (NICE)-recommended PTSD treatment (trauma-focused cognitive behavioral therapy or eye movement desensitization and reprocessing), with those viewing the mother vignette significantly more likely to do so (40.4%) than those viewing the foster care vignette (27.2%; χ² (1) = 4.94, p = .03, OR = 1.82). Although the overall diagnostic accuracy could not be determined, the study estimated that 16.5% of the variance in PTSD diagnosis was attributable to diagnostic expectation biases. These findings suggest that contextual factors, such as a child’s living arrangement, may unconsciously influence clinicians’ diagnostic judgments, potentially contributing to the under-recognition of PTSD. Further, even among those who selected PTSD as the primary diagnosis, only 52.7% went on to recommend a NICE-recommended treatment, underscoring additional barriers to evidence-based care. Overall, this study demonstrated a low to moderate risk of bias. However, the absence of participant and personnel blinding, as well as the potential for scenario-related biases, may introduce some uncertainty into the results.

McKenzie and Smith [65] conducted a cross-sectional study that investigated the knowledge of general practitioners (GPs), psychologists, and psychiatrists regarding PTSD to identify their understanding, beliefs, and knowledge gaps. The study recruited 154 MH professionals in Victoria, Australia, including 59 GPs, 56 psychologists, and 39 psychiatrists, all registered with the Department of Veterans’ Affairs. Participants completed 2 questionnaires: the General Information Questionnaire and the PTSD Knowledge Questionnaire, which assessed knowledge across 8 domains, including symptomatology, comorbidity, and treatment. Across all groups, the mean percentage of correct responses was 74% (standard deviation = 12%), incorrect responses was 11% (standard deviation = 6%), and uncertain responses was 15% (standard deviation = 11%). Knowledge scores varied: GPs averaged 68% correct, psychologists averaged 77%, and psychiatrists averaged 79%. Notably, GPs demonstrated significantly less familiarity with the DSM-IV criteria and expressed greater uncertainty in their responses. Psychologists and psychiatrists scored significantly higher than GPs on the symptom and course domains (p < .001, d = 1.18 and 0.82 respectively), and psychiatrists scored significantly higher in understanding that PTSD is not a normal response to trauma. Although the study did not report direct diagnostic accuracy, it found substantial variation in clinician knowledge, with critical gaps that may adversely affect PTSD identification. Self-rated familiarity with PTSD was poorly correlated with actual performance across all groups, indicating that many clinicians were unaware of their knowledge gaps. The study concluded that inadequate clinician training likely impairs diagnostic accuracy and recommended targeted educational interventions to address these deficiencies. This study showed a moderate risk of bias due to potential selection and response bias, which may limit generalizability.

### Evidence of PTSD underdiagnosis in clinical settings (studies not included in the meta-analysis)

Extensive research demonstrates that PTSD remains significantly underdiagnosed in both primary and specialized MH settings. Studies employing structured interviews, EMR analysis, clinician surveys, and patient data consistently reveal that diagnostic accuracy in routine clinical practice is low due to systemic, procedural, and clinician-related factors.

Bovin et al. [67] conducted a cross-sectional diagnostic validation study of the PC-PTSD-5 among 396 veterans in VA primary care settings in the US, using the CAPS-5 as a structured diagnostic reference. The study found that 17.2% of participants met PTSD diagnostic criteria according to the CAPS-5 (15% of men, 29% of women). The PC-PTSD-5 demonstrated high diagnostic accuracy, with an area under the curve of 0.927 overall and similarly strong performance across sexes. The optimal cut-point varied by sex: a cut-point of 4 maximized efficiency for men but missed 33.3% of PTSD-positive women, prompting the authors to recommend a lower threshold of 3 for female patients in some settings.

Although the study did not assess real-world diagnostic coding or provider recognition, the authors noted that most participants who met the criteria for PTSD were not receiving specialty MH at the time of the study. This gap highlights a potential failure of detection in routine care and supports the need for systematic screening. Exploratory analyses also showed that most false negatives (i.e., missed cases using PC-PTSD-5) would likely still be flagged by other routine VA screens (e.g., 9-item Patient Health Questionnaire, AUDIT-C), suggesting partial mitigation of underdetection in practice. Nonetheless, the authors emphasized that false negatives represent a missed opportunity for early PTSD intervention, especially among women.

The study concluded that, while the PC-PTSD-5 is highly accurate and acceptable to veterans, screening cut points should be tailored to the population and clinical goals, balancing sensitivity with efficiency. These findings underscore the importance of ongoing evaluation and sex-specific considerations in PTSD screening tools used in primary care.

Potential study limitations include limited generalizability beyond VA primary care populations and potential non-response bias, as only 31% of eligible veterans participated.

Kosowan et al. [68] conducted a retrospective diagnostic coding analysis study that analyzed the Manitoba Primary Care Research Network dataset within the national Canadian Primary Care Sentinel Surveillance Network EMR system, which comprises 289,523 patients. They found that the PTSD diagnosis rate was only 1.3%, demonstrating that diagnostic coding alone substantially underestimates the prevalence of PTSD. Using both structured diagnostic codes from the International Classification of Diseases, Ninth Revision, Clinical Modification, and unstructured short diagnostic text fields, they tested 4 case definitions and found that the most comprehensive method (case definition 4) identified PTSD with 91.1% sensitivity and 99.1% specificity. However, prevalence estimates still remained low (0.8%–1.3%), reinforcing concerns about under detection. The study further showed that free-text encounter notes, when available, did not dramatically improve detection compared to short diagnostic text fields, though a subset of patients exhibited PTSD symptoms without receiving a formal diagnosis. Their study advocated for advanced methods, such as natural language processing, to extract trauma-related information from free-text clinical notes and improve detection. They concluded that integrating natural language processing into case definitions can enhance diagnostic capture, particularly in systems lacking standardized coding practices or access to detailed narrative documentation.

This study may be limited by the potential underestimation of PTSD due to provider variability in documentation and diagnostic coding, as well as the reliance on algorithmically defined cases without validation from structured clinical interviews.

Singer et al. [69] conducted a retrospective observational EMR-based study that reinforced these findings using the same Canadian Primary Care Sentinel Surveillance Network EMR dataset, analyzing 689,301 primary care patients across 7 provinces. They found that only 1.3% (n = 8,817) had a recorded PTSD diagnosis, consistent with prior EMR-based prevalence estimates. Using logistic regression analysis, the study found that PTSD was strongly associated with comorbid depression (aOR = 4.4), anxiety (aOR = 2.5), alcohol use disorder (aOR = 1.7), and drug use disorder (aOR = 2.6). Patients with PTSD were also significantly more likely to reside in areas of high material (aOR = 2.1) and social deprivation (aOR = 3.8).

Notably, PTSD was often documented years after other MH conditions: on average, depression (3.5 years), anxiety (3.3 years), and substance use disorders (3.7 years) were recorded before PTSD. This lag suggests that PTSD frequently exists as a hidden or secondary comorbidity, identified only after prolonged treatment for overlapping conditions. Despite this, individuals with PTSD had more frequent primary care visits and higher medication use, indicating elevated healthcare needs.

The authors concluded that PTSD is under-recognized in routine care, especially when presented alongside other psychiatric or social risk factors. They recommended enhanced screening and resource allocation in primary care to improve PTSD detection and management, especially among medically and socially complex patients.

The study relied on EMR data, which may underreport PTSD due to inconsistent or missing documentation. Additionally, social and material deprivation data were available for only a small subset (8%) of the cohort, limiting the generalizability of those findings.

Greene et al. [26] conducted a systematic review of 27 primary care studies comparing structured PTSD assessments with provider documentation. The review included 23,941 patients, with individual study sample sizes ranging from 134 to 4,416. They reported that the prevalence of PTSD in primary care samples ranged from 2% to 39%, depending on trauma exposure and population characteristics. Detection rates by primary care physicians ranged from 0% to 52%, with a weighted average of 15.46% across studies. The review highlighted that structured assessments, such as the CAPS, SCID, and CIDI, consistently identified more PTSD cases than physician judgment or medical record review alone. Studies that relied on patient self-reports or structured interviews uncovered significantly higher PTSD rates, indicating that routine clinical detection is often insufficient. The review also noted high psychiatric comorbidity and functional impairment among individuals with PTSD, reinforcing the clinical importance of improving recognition. These findings suggest that structured tools identify substantially more PTSD cases than clinical judgment alone and underscore the need for routine screening and provider education in primary care.

As a systematic review, the study was limited by heterogeneity in methodologies across the included studies, particularly the variation in diagnostic tools and trauma exposure, which complicates comparisons and pooled estimates.

Ehlers et al. [70] surveyed GPs in the UK in their cross-sectional survey study and found a severe underestimation of PTSD prevalence. A total of 129 GPs responded to the survey (an 18% response rate), and they reported a median estimated PTSD prevalence of just 0.2% among their patient panels, despite epidemiological data suggesting that the true prevalence was between 1.5% and 3.6%. Twenty-seven percent of respondents had not diagnosed a single adult case in the past 3 months, and 71% had not seen any children with PTSD, indicating a substantial diagnostic blind spot. Although 55% were aware of the NICE PTSD guidelines, only 15% had read them, and only 27% were aware that they included recommendations for children. Treatment practices were also inconsistent with the guidelines. Only 11% of GPs reported offering the recommended 8 to 12 sessions of trauma-focused cognitive behavioral therapy or eye movement desensitization and reprocessing, and most referrals were limited by long waiting times or a lack of awareness of trauma services. Instead, medications, particularly SSRIs, were prescribed in 52% of cases, often as first-line treatments, despite NICE’s advice to the contrary. These results reflect diagnostic blind spots driven by limited training, overreliance on comorbid symptoms, and a low confidence or awareness regarding trauma-focused care.

The low GP response rate (18%) raises the possibility of selection bias, as participants may have been more interested in or aware of PTSD, potentially overestimating their knowledge and underestimating diagnostic blind spots among GPs in general.

Della Porta [71] conducted an experimental vignette study using a randomized design to evaluate diagnostic reasoning for PTSD among primary care physicians in the US Participants (N = 144) were randomly assigned to one of two clinical vignettes describing a patient meeting DSM-5 PTSD criteria: one presented a straightforward PTSD case, and the other depicted PTSD comorbid with multiple physical complaints. Participants were asked to record up to 3 diagnoses after reviewing the vignette, and their performance was analyzed based on the inclusion of PTSD-related terms.

Overall, 90.3% of physicians (130 out of 144) accurately identified PTSD, with similar accuracy across vignettes (91.0% for straightforward PTSD vs. 89.1% for PTSD comorbid with multiple physical complaints), indicating that physical symptom comorbidity did not significantly impair recognition. Contrary to expectations, neither clinical experience nor PTSD knowledge scores predicted diagnostic accuracy. Years of experience (1–10 vs. 11+ years) were not significantly associated with detection rates (χ² = 0.075, p = .784), and logistic regression analysis indicated that PTSD knowledge did not significantly predict diagnostic success (p = .331).

As the authors note, their findings stand in “stark contrast” to prior studies reporting underdiagnosis of PTSD in primary care, and they caution against interpreting this solely as evidence of improved clinician detection abilities. They acknowledged several limitations, including potential priming effects, a lack of ecological validity, and the use of simplified case vignettes. The trauma event was always disclosed as the reason for the visit, which may have increased diagnostic salience compared to real-world encounters. Importantly, the study supports the notion that when PTSD symptoms are clearly presented, primary care physicians are capable of accurate diagnosis; however, it does not reflect the typical diagnostic challenges posed by more ambiguous or somatically masked presentations.

This study demonstrates a low risk of bias in its design but acknowledges limited generalizability due to its vignette-based format and participant self-selection. Nevertheless, its results suggest that PTSD may go unrecognized in clinical practice not due to a lack of diagnostic skill per se, but because of how symptoms are presented, the absence of structured assessment tools, or competing clinical priorities.

Note: This study [71] was excluded from the meta-analysis because its vignette-based design involved clearly defined PTSD presentations, which differs from studies assessing clinician diagnostic accuracy and biases in naturalistic settings [65,66]. While it demonstrates clinicians’ capability to identify PTSD when symptoms are explicit, it does not capture the complexity and ambiguity encountered in routine clinical practice, limiting its comparability with empirical diagnostic accuracy data.

Kaltman et al. [72] conducted a prospective comparative cohort study of low-income immigrant women in primary care and found that trauma-related symptoms were frequently overlooked due to language barriers, cultural stigma, and insufficient provider training. The study was conducted in 2 clinics in the US serving uninsured, immigrants from Central and South America. Of the 138 patients who completed baseline and follow-up assessments, 83.3% reported trauma exposure, and approximately two-thirds screened positive for PTSD at baseline (67.7% in the collaborative care clinic and 64.4% in the on-site therapist clinic). Structured interviews revealed high rates of PTSD that were not reflected in patients’ medical records. More than half of all participants received no care or care that fell below a minimal threshold of adequacy, and many comorbid cases of PTSD and depression went untreated. The collaborative care model, which included proactive screening and integrated care management, was associated with greater symptom reduction in PTSD compared to standard co-located services. These findings highlight persistent disparities in trauma recognition and access to care in under-resourced primary care environments.

This study was limited by a small sample size from 2 clinics serving uninsured immigrant populations. The use of self-report tools and the absence of clinician-confirmed diagnoses could lead to misclassification. Findings may not generalize to broader primary care settings.

Cook et al. [73] conducted a retrospective cohort study that analyzed data from 5,256 individuals across 5 large US civilian healthcare systems. They found that only 16.6% of PTSD diagnoses were made in primary care or other non-mental health clinics, while 83.4% were made by specialty mental health care (MHC) providers. Individuals diagnosed in MHC settings received significantly more treatment: they were 4 times more likely to receive psychotherapy (incidence rate ratio [IRR] = 4.25, p < .001) and had nearly 1.5 times higher rates of psychotropic medication use (IRR = 1.48, p < .001) compared to those diagnosed in primary care.

Treatment adequacy also varied significantly. While 79.3% of patients diagnosed in MHC received psychotherapy, only 24.4% of those diagnosed in primary care did. Similarly, 67.2% of MHC patients had PTSD-diagnosed psychotherapy sessions, compared to just 8.1% in the primary care group. Even when treatment was initiated, most patients did not receive the recommended course of 9 or more therapy sessions, highlighting a broader gap in the sufficiency of PTSD treatment.

The authors concluded that substantial disparities exist in both PTSD detection and the quality of treatment between care settings. They attributed this gap partly to the absence of routine PTSD screening in civilian primary care and emphasized the need for stronger referral systems, training in trauma-informed care, and the implementation of evidence-based guidelines.

The study used administrative data from 5healthcare systems but lacked standardized diagnostic tools across the sites. This introduces heterogeneity in diagnostic practices, and the reliance on coded diagnoses limits the assessment of underdiagnosis or false negatives.

Bruce et al. [74] conducted a prospective, naturalistic longitudinal study with PTSD-related findings based on cross-sectional baseline data from 504 participants in 14 US primary care clinics, all of whom met the diagnostic criteria for at least 1 anxiety disorder. Using the SCID-I and the Trauma Assessment for Adults, the study found high rates of trauma exposure and PTSD symptoms: 83% of participants reported experiencing at least 1 traumatic event, and 44% of those met the DSM-IV criteria for PTSD. In total, 185 individuals were identified with PTSD, many of whom had experienced multiple traumas and comorbid conditions such as major depression (43%), alcohol/substance abuse (62%), and suicide attempts (33%). Despite the clear symptom burden, the study highlighted that PTSD symptoms often go unrecognized in primary care due to overlapping presentations with more familiar disorders (e.g., depression and Generalized Anxiety Disorder), time constraints, and clinicians not routinely inquiring about trauma histories. The authors noted that trauma-related symptoms may be underreported by patients due to embarrassment or avoidance, which further contributes to underdiagnosis. These results underscore the importance of routine trauma screening and the use of validated diagnostic tools, as well as training providers to recognize PTSD, even when symptoms present in somatic or nonspecific ways.

The study’s reliance on self-reported trauma exposure and structured interviews conducted in a single urban clinic limits its generalizability. There is also the potential for selection bias, as participants willing to discuss trauma may systematically differ from those who declined participation.

Neria et al. [75] conducted a cross-sectional survey study to assess the prevalence, correlates, and treatment patterns of probable PTSD in a low-income, predominantly Hispanic sample of 930 adult primary care patients in the US 7 to 16 months after the 9/11 attacks. Participants were systematically recruited from an urban general medicine clinic and screened using the PCL-C, with probable PTSD defined using both a symptom cluster-based DSM-IV algorithm and a strict cut-off score of 50. The prevalence of current 9/11-related probable PTSD ranged from 4.7% (strict cut-off) to 10.2% (symptom cluster definition). PTSD was significantly associated with female sex, Hispanic ethnicity, immigrant status, unmarried status, family psychiatric history, and prior trauma exposure.

Most individuals with PTSD screened positive for at least 1 other mental disorder (68.4%), with major depression (57.9%) and generalized anxiety disorder (33.7%) being the most common. Compared to PTSD-negative individuals, those with PTSD showed significantly greater impairment in family and social functioning, poorer physical and MH-related quality of life, and more frequent work loss (73% vs. 26.5%). However, despite 50% reporting recent psychotropic medication use and 40% reporting MH treatment, a PTSD diagnosis was often absent from EMRs. Further, PTSD was not associated with increased hospital, emergency room, or outpatient service use during the year following 9/11 based on administrative records.

These findings highlight missed opportunities for clinical intervention and suggest a potential disconnect between symptom burden and formal diagnosis or service delivery. The study’s strengths include the use of validated instruments, a large and diverse clinical sample, and linkage to service use records. Limitations include reliance on self-reports for PTSD symptoms and treatment, the absence of structured diagnostic interviews, and restriction to one healthcare system, which may have led to an underestimation of external service use. Nevertheless, the results underscore the unmet MH needs of underserved primary care patients following mass trauma and emphasize the need for proactive screening and trauma-informed care models in general medical settings.

Spottswood et al. [27] conducted a systematic review of 41 studies, assessing a combined total of 7,256,826 primary care patients to estimate the prevalence of PTSD in primary care populations. The included studies represented diverse settings and patient groups (civilians, veterans, and other high-risk populations) and utilized a range of diagnostic methods, including structured clinical interviews, validated self-report measures, and administrative data. Across all studies, the median point prevalence of PTSD in primary care was 12.5%, with notable variation by population group: civilians (11.1%), veterans (24.5%), and special risk groups (12.5%). Prevalence also differed by assessment method: structured interviews yielded rates ranging from 2.0% to 32.5%, questionnaires from 2.9% to 39.1%, and administrative data from 3.5% to 48.8%.

The authors found that PTSD prevalence estimates were generally higher in studies using structured diagnostic tools compared to chart reviews or administrative coding, suggesting under-recognition and limited clinical detection in routine care. They rated most studies as having a low to moderate risk of bias and concluded that PTSD is both prevalent and frequently underdiagnosed in primary care settings. The review emphasized the need for standardized PTSD screening, stepped care pathways, and collaborative care models to improve detection and treatment outcomes in frontline clinical environments.

Sareen et al. [76] conducted a cross-sectional epidemiological study that analyzed data from the Canadian Community Health Survey Cycle 1.2 (n = 36,984) to examine the association of PTSD with comorbid physical and mental conditions, disability, and suicidality. The prevalence of PTSD was 1.0%, based on self-reported professional diagnoses. Even after adjusting for sociodemographic variables and other mental disorders, PTSD was independently associated with a wide range of chronic physical conditions, including cardiovascular diseases, respiratory illnesses, chronic pain, gastrointestinal disorders, and cancer. PTSD was also strongly associated with poor psychological well-being, high levels of distress, short- and long-term disability, and suicide attempts. The study underscores the importance of early clinical detection and treatment of PTSD, especially since many affected individuals remain undiagnosed and untreated. The authors emphasized that improved screening in general medical settings is crucial for addressing the broader public health burden associated with PTSD.

The study relied on self-reported professional PTSD diagnoses rather than structured clinical interviews, which likely underestimated the true prevalence of PTSD and may have biased the sample toward individuals with more severe or treated cases.

Cowlishaw et al. [77] conducted a cross-sectional survey study across 11 general practices in southwest England to assess the prevalence and correlates of probable PTSD in routine UK primary care. A total of 1,058 adult patients were recruited from clinic waiting rooms and completed anonymous questionnaires, including PC-PTSD, 2-item Generalized Anxiety Disorder scale, Whooley scale for depression, and AUDIT-C for alcohol use. A cut-off score of ≥ 3 on the PC-PTSD identified probable PTSD.

Overall, 15.1% of respondents met the criteria for probable PTSD, with the prevalence highest in practices located in highly deprived areas (19.0%) compared to those in moderate (10.9%) and low-deprivation practices (12.5%). Among those who screened positive, 54% expressed a desire for help, indicating high treatment receptivity. Multivariable analyses showed that patients who were younger, unemployed, or not in cohabiting relationships were at significantly higher risk of PTSD. Depression and anxiety were particularly strong correlates; depression was associated with a 15.6-fold increase in probable PTSD (OR = 15.55), and anxiety with a 9.8-fold increase (OR = 9.78). However, no association was found between probable PTSD and risky alcohol use.

Although the study did not evaluate diagnostic recognition or EMR data, the authors inferred that a substantial portion of PTSD cases likely remains undetected in general practice. They emphasized the utility of routine screening (e.g., PC-PTSD) and advocated for enhanced practitioner training and care pathways to trauma-focused treatments. The study had a moderate risk of bias due to regional sampling, missing data managed through zero-fill techniques, and the use of a brief screening tool that was not calibrated to subthreshold symptoms or DSM-5 criteria. Despite these limitations, it provides important UK-based prevalence data and supports the inclusion of PTSD screening in general practice.

Jiang et al. [78] conducted a psychometric validation study of the PCL-5 in a sample of 348 Chinese stroke patients, using the CAPS-5 as the diagnostic reference. The study aimed to assess the reliability, validity, and optimal cut-off scores of the PCL-5 in a non-psychiatric clinical population. Receiver operating characteristic analysis yielded an area under the curve of 0.96, demonstrating excellent diagnostic performance. A cut-off score of 37 optimized sensitivity (0.95) and specificity (0.89), demonstrating high internal consistency (Cronbach’s alpha = 0.95) and test-retest reliability (intraclass correlation coefficient = 0.87).

Although the study confirms the diagnostic precision of structured PTSD tools like the PCL-5 and CAPS-5, it also highlights the challenges of implementing time-intensive assessments in routine practice. The authors noted that clinicians often rely on brief screeners, which may miss more complex or subtle cases.

The use of stroke patients as the validation sample may limit generalizability to broader primary care populations, and the clinical reference standard (CAPS-5) was administered in a single setting, which could affect external validity.

Gillock et al. [36] conducted a cross-sectional study in a civilian primary care setting in the US to assess the prevalence and clinical recognition of PTSD. A total of 529 adult patients were recruited from an urban general medical clinic, and 232 completed all assessments and were included in the final analysis. Participants were screened using structured questionnaires based on DSM-IV criteria. Results showed that 9% met the full criteria for current PTSD, and an additional 25% met partial criteria, indicating a high burden of trauma-related distress in this population. Despite this, PTSD was not documented in participants’ medical records, and those with PTSD were found to use significantly more medical services, report greater physical symptom severity, and exhibit higher levels of psychological distress. The authors concluded that trauma symptoms were likely being treated somatically without psychiatric recognition, which led to systematic underdiagnosis and missed opportunities for MH. The study had a moderate risk of bias due to its single-site sample and the absence of a clinical documentation review to directly assess provider recognition.

The study is limited by its single-site design, modest sample size, reliance on a paper-based diagnostic tool rather than structured clinical interviews such as CAPS or SCID, and the absence of clinician interviews to verify diagnostic blind spots. These factors contribute to a moderate risk of bias.

Nakash et al. [79] conducted an observational validation study that examined diagnostic practices during 122 intake sessions at 4 community MH clinics in Israel. Clinicians conducted unstructured DSM-IV-based evaluations, while participants completed SCIDs with independent assessors. The study found that clinicians frequently underutilized the DSM framework and often failed to collect sufficient information to meet the diagnostic criteria. This resulted in the under-recognition of psychiatric conditions. Although PTSD-specific diagnostic accuracy was not isolated, the general pattern of insufficient symptom inquiry extended across multiple disorders. The authors concluded that structured assessment tools or focused symptom probes are necessary to improve diagnostic accuracy in standard clinical care.

This study has a moderate risk of bias, primarily due to its focus on overall diagnostic accuracy rather than on disorder-specific metrics such as PTSD. However, the use of both clinician-conducted and independently structured interviews adds methodological strength.

Together, these 16 studies highlight the widespread under-recognition of PTSD in clinical settings. Primary contributing factors include the inconsistent use of validated tools, structural time constraints, diagnostic overshadowing by comorbidities, and limited trauma training. This evidence underscores the need for standardized screening protocols and trauma-informed care models across all healthcare sectors.

### Meta-analysis

As shown in Table 1, the weighted mean PTSD diagnostic accuracy in specialized MH settings (Group 1), calculated from 20 of the included studies, was 55.6% of the total number of participants with true PTSD in each study. In contrast, the weighted mean diagnostic accuracy in primary healthcare settings (Group 2), based on 11 studies and weighted by the total number of participants with true PTSD 48.2% (Table 2). Finally, 2 studies were included to assess clinicians’ PTSD-related knowledge and their ability to accurately diagnose PTSD despite contextual biases (Group 3). Based on the weighted mean from these studies, the estimated potential PTSD diagnostic accuracy was 80.0%. This estimate reflects two complementary methodologies: (a) clinicians’ ability to identify PTSD in hypothetical patients presenting with identical symptomatology but differing contextual backgrounds (e.g., foster care vs. biological family), and (b) their domain-specific knowledge of PTSD symptoms and diagnostic criteria (Table 3). The “Total” columns on all 3 tables reflect only those participants who completed the full diagnostic confirmation process. This excludes individuals who were initially recruited or screened but did not undergo diagnostic verification, ensuring accuracy and consistency across all studies.

**Table 1.**
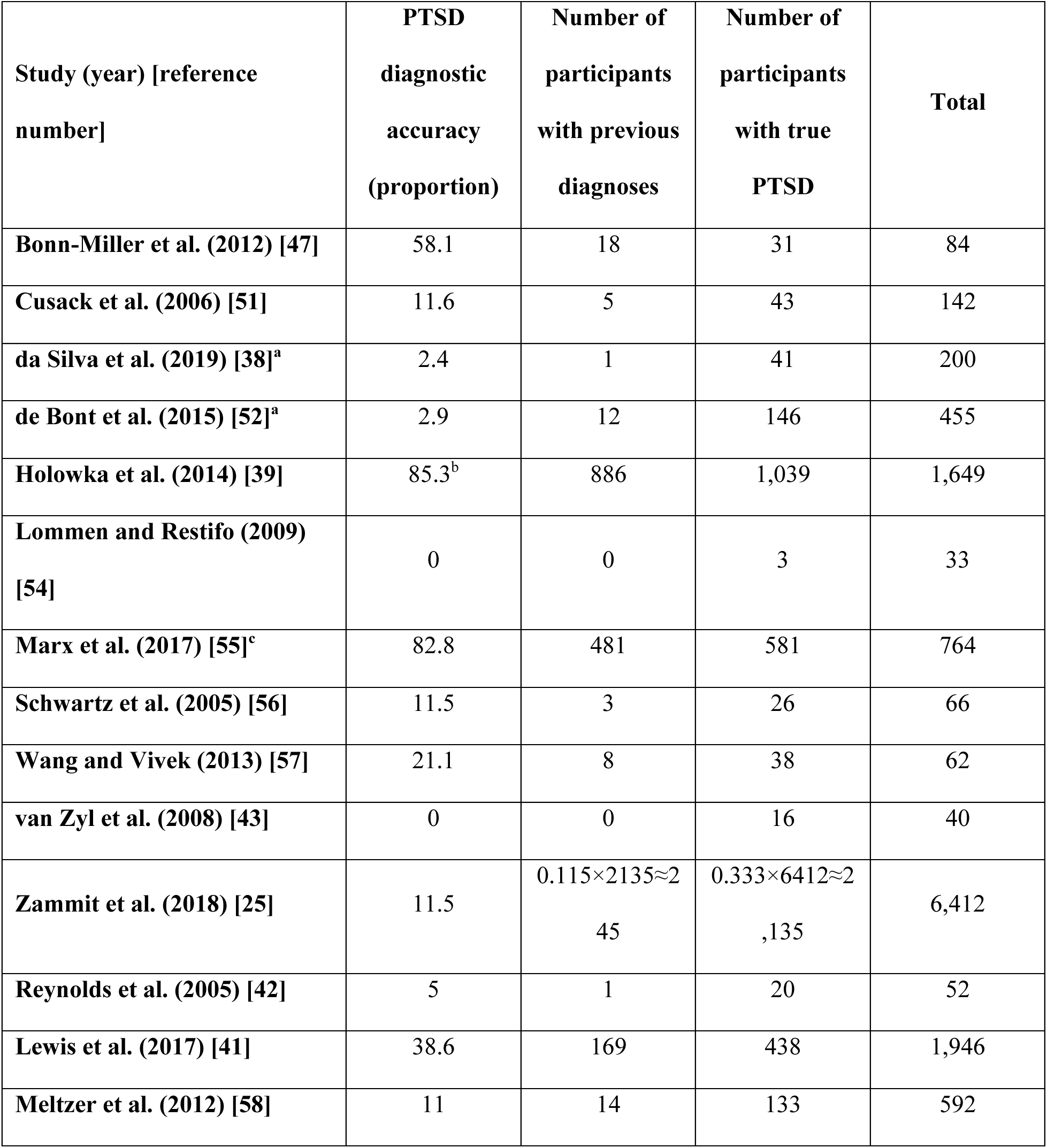
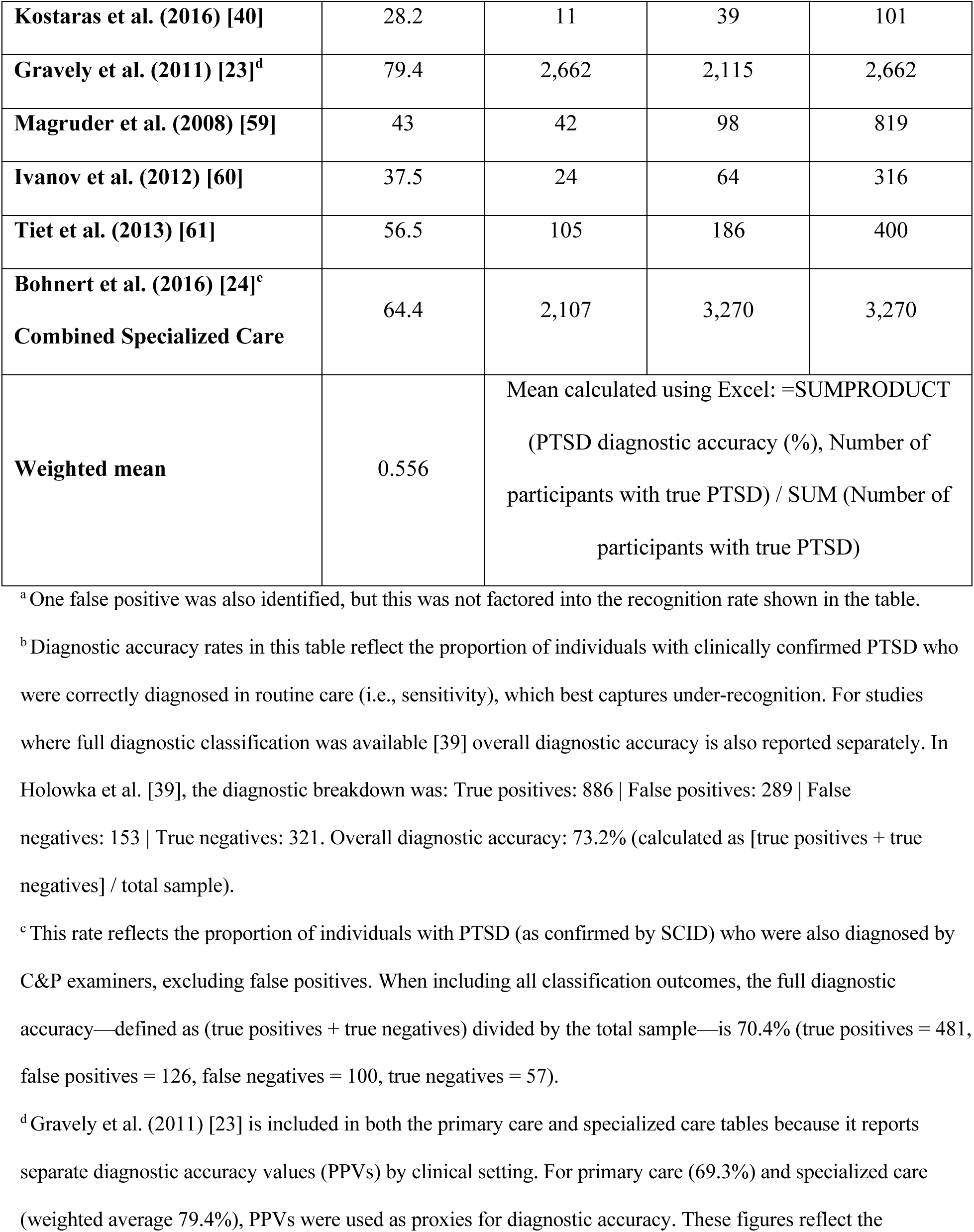
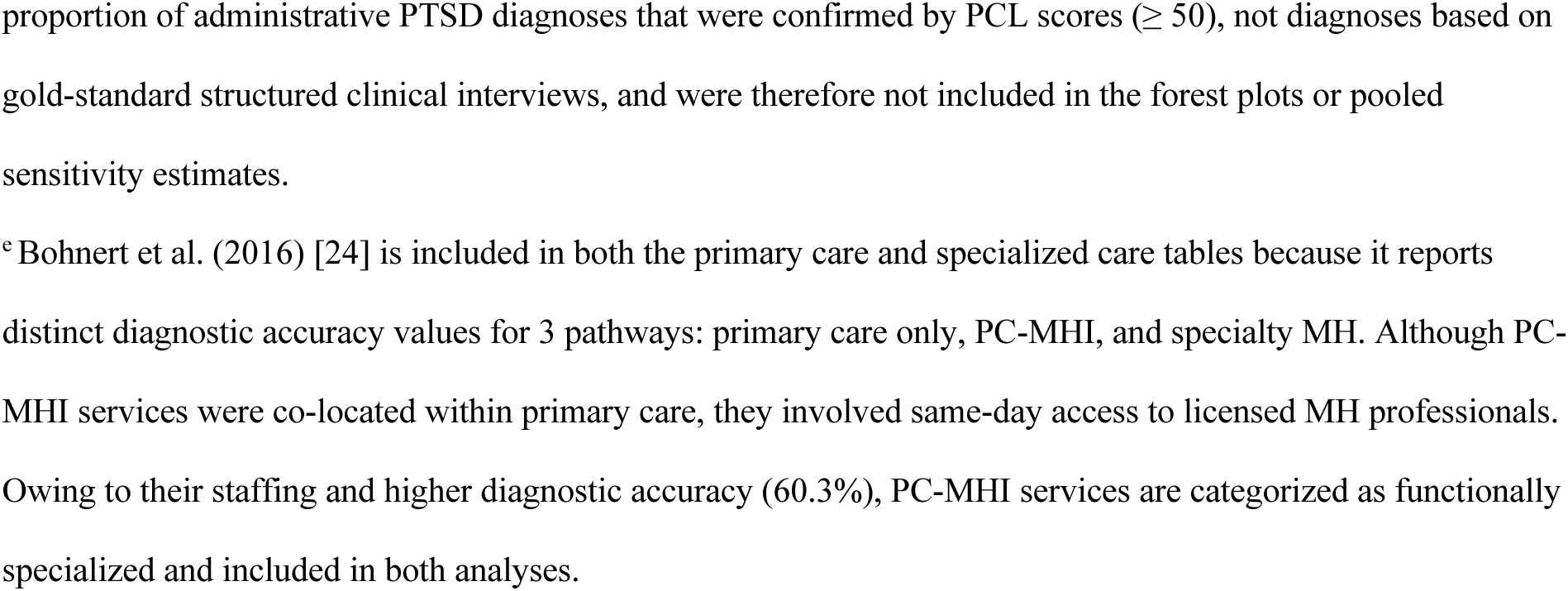
PTSD diagnostic accuracy/recognition rate in mental health settings.

**Table 2.** PTSD diagnostic accuracy/recognition rate in primary care settings.

| Study (year) [reference number] | PTSD diagnostic accuracy (proportion) | Number of participants with previous diagnoses | Number of participants with true PTSD | Total |
| --- | --- | --- | --- | --- |
| Gravely et al. (2011) [23]<sup>a</sup> | 69.3 | 2,071 | 1,435 | 2,071 |
| Bohnert et al. (2016) [24]<sup>b</sup><br>Primary Care Only | 47.9 | 8,703 | 18,157 | 18,157 |
| Meltzer-Brody et al. (2004) [62] | 12<sup>c</sup> | 3 | 25 | 32 |
| Prins et al. (2003) [29] | 39.1 | 11 | 28 | 134 |
| Taubman-Ben-Ari et al. (2001) [30]<sup>d</sup> | 2.40 | 6 | 247 | 2,975 |
| Graves et al. (2011) [31] | 30.8 | 28 | 91 | 375 |
| Liebschutz et al. (2007) [32] | 11.1 | 13 | 117 | 597 |
| **Lu et al. (2023) [63]** | 23.9 | 16 | 67 | 135 |
| **Carey et al. (2003) [33]** | 0 | 0 | 40 | 201 |
| **Seal et al. (2008) [64]** | 24<sup>e</sup> | 41 | 172 | 338 |
| **Kimerling et al. (2006) [34]** | 38.2 | 13 | 34 | 134 |
| **Weighted mean** | 0.482 | Mean calculated using Excel: =SUMPRODUCT<br>(PTSD diagnostic accuracy (%), Number of<br>participants with true PTSD) / SUM (Number of<br>participants with true PTSD) |  |  |

<sup>a</sup> Gravely et al. (2011) [23] is included in both the primary care and specialized care tables because it reports separate diagnostic accuracy values (PPVs) by clinical setting. For primary care (69.3%) and specialized care (weighted average 79.4%), PPVs were used as proxies for diagnostic accuracy. These figures reflect the proportion of administrative PTSD diagnoses that were confirmed by PCL scores ( $\geq 50$ ), not diagnoses based on gold-standard structured clinical interviews, and were therefore not included in the forest plots or pooled sensitivity estimates.
<sup>b</sup> Bohnert et al. (2016) [24] is included in both the primary care and specialized care tables because it reports distinct diagnostic accuracy values for 3 pathways: primary care only, PC-MHI, and specialty MH. Although PC-MHI services were co-located within primary care, they involved same-day access to licensed MH professionals. Owing to their staffing and higher diagnostic accuracy (60.3%), PC-MHI services are categorized as functionally specialized and included in both analyses.
<sup>c</sup> Estimation based on the proportion of PTSD-positive women receiving psychiatric care at assessment. This measure does not confirm documented PTSD diagnoses or specific PTSD treatment and is therefore less precise and likely overestimates true diagnostic accuracy compared to other studies using structured clinical interviews. This limitation is accounted for in the review's analysis and interpretation.
<sup>d</sup> Eight false positives were also identified, but these were not factored into the recognition rate shown in the table.
<sup>e</sup> Diagnostic accuracy is estimated from follow-up mental health appointments after positive PTSD screening rather than structured clinical interviews. This proxy measure is less reliable than gold-standard diagnostics but
offers an approximate indication of PTSD recognition and referral in routine care.

**Table 3.** Average MH specialists’ ability to accurately diagnose PTSD.

| Study (year) [reference number] | PTSD diagnostic competence:<br>Knowledge and contextual accuracy (proportion) | Number of professionals |
| --- | --- | --- |
| McGuire et al. (2022) [66] | 22.7 | 270 |
| McKenzie and Smith (2006) [65] | 74 | 154 |
| **Weighted mean** | 0.4133 | Mean calculated using Excel<br>SUMPRODUCT: =<br>SUMPRODUCT (Accuracy %, N professionals)/SUM (N professionals) |

The meta-analysis was conducted using the following steps. Calculation of standard errors (Table 4): Standard errors (SEs) associated with diagnostic accuracy rates were calculated for each study based on the number of participants who met PTSD criteria using a structured diagnostic interview or a validated screener.

**Table 4.** Standard errors for each of the included studies.

| Study (year) [reference number] | Standard errors |
| --- | --- |
| **Bonn-Miller et al. (2012) [47]** | $\sqrt{\frac{0.581*(1-0.581)}{31}} = 0.0886$ |
| **Cusack et al. (2006) [51]** | $\sqrt{\frac{0.116*(1-0.116)}{43}} = 0.0488$ |
| **da Silva et al. (2019) [38]** | $\sqrt{\frac{0.024*(1-0.024)}{41}} = 0.0239$ |
| **de Bont et al. (2015) [52]** | $\sqrt{\frac{0.029*(1-0.029)}{146}} = 0.0139$ |
| **Holowka et al. (2014) [39]** | $\sqrt{\frac{0.853*(1-0.853)}{1039}} = 0.011$ |
| **Lommen and Restifo (2009) [54]** | $\sqrt{\frac{0*(1-0)}{3}} = 0.0$ |
| **Marx et al. (2017) [55]** | $\sqrt{\frac{0.828*(1-0.828)}{581}} = 0.0157$ |
| **Schwartz et al. (2005) [56]** | $\sqrt{\frac{0.0115*(1-0.0115)}{26}} = 0.0209$ |
| **Wang and Vivek (2013) [57]** | $\sqrt{\frac{0.211*(1-0.211)}{38}} = 0.0662$ |
| **van Zyl et al. (2008) [43]** | $\sqrt{\frac{0*(1-0)}{16}} = 0.0$ |
| **Zammit et al. (2018) [25]** | $\sqrt{\frac{0.115*(1-0.115)}{2135}} = 0.0069$ |
| **Reynolds et al. (2005) [42]** | $\sqrt{\frac{0.05*(1-0.05)}{20}} = 0.0487$ |
| **Lewis et al. (2017) [41]** | $\sqrt{\frac{0.386*(1-0.386)}{438}} = 0.0233$ |
| **Meltzer et al. (2012) [58]** | $\sqrt{\frac{0.11*(1-0.11)}{133}} = 0.0271$ |
| **Kostaras et al. (2016) [40]** | $\sqrt{\frac{0.282*(1-0.282)}{39}} = 0.0721$ |
| **Gravely et al. (2011) [23] – Specialized Care** | $\sqrt{\frac{0.794*(1-0.794*)}{2115}} = 0.0088$ |
| **Gravely et al. (2011) [23] – Primary Care** | $\sqrt{\frac{0.693*(1-0.693)}{1435}} = 0.0122$ |
| **Magruder et al. (2008) [59]** | $\sqrt{\frac{0.43*(1-0.43)}{98}} = 0.0500$ |
| **Ivanov et al. (2012) [60]** | $\sqrt{\frac{0.375*(1-0.375)}{64}} = 0.0605$ |
| **Tiet et al. (2013) [61]** | $\sqrt{\frac{0.565*(1-0.565)}{186}} = 0.0364$ |
| **Bohnert et al. (2016) [24] - Combined**<br>**Specialized Care** | $\sqrt{\frac{0.644*(1-0.644)}{3270}} = 0.0084$ |
| **Bohnert et al. (2016) [24] – Primary Care**<br>**Only** | $\sqrt{\frac{0.479*(1-0.479)}{18157}} = 0.00371$ |
| **Meltzer-Brody et al. (2004) [62]** | $\sqrt{\frac{0.12*(1-0.12)}{25}} = 0.0650$ |
| **Prins et al. (2003) [29]** | $\sqrt{\frac{0.391*(1-0.391)}{28}} = 0.0922$ |
| **Taubman-Ben-Ari et al. (2001) [30]** | $\sqrt{\frac{0.024*(1-0.024)}{247}} = 0.0097$ |
| **Graves et al. (2011) [31]** | $\sqrt{\frac{0.308*(1-0.308)}{91}} = 0.0484$ |
| **Liebschutz et al. (2007) [32]** | $\sqrt{\frac{0.111*(1-0.111)}{117}} = 0.0290$ |
| **Lu et al. (2023) [63]** | $\sqrt{\frac{0.239*(1-0.239)}{67}} = 0.0521$ |
| **Carey et al. (2003) [33]** | $\sqrt{\frac{0.0*(1-0.0)}{40}} = 0.0$ |
| Seal et al. (2008) [64] | $\sqrt{\frac{0.24*(1-0.24)}{172}} = 0.0326$ |
| Kimerling et al. (2006) [34] | $\sqrt{\frac{0.382*(1-0.382)}{34}} = 0.0833$ |
| McGuire et al. (2022) [66] | $\sqrt{\frac{0.227*(1-0.227)}{270}} = 0.0255$ |
| McKenzie and Smith (2006) [65] | $\sqrt{\frac{0.74*(1-0.74)}{154}} = 0.0353$ |

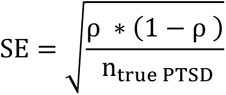

where p represents the diagnostic accuracy rate and n_true PTSD_ is the number of participants who met PTSD criteria according to the reference standard.

Calculation of CIs (Table 5): 95% CIs were calculated using the following formula:

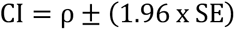

**Table 5.** Confidence intervals for each of the included studies.

| Study (year) [reference number] | Estimate | 95% Confidence intervals |
| --- | --- | --- |
| Bonn-Miller et al. (2012) [47] | 0.581 | $0.581 \pm 1.96 * 0.0886 = [0.4073, 0.7547]$ |
| Cusack et al. (2006) [51] | 0.116 | $0.116 \pm 1.96 * 0.0488 = [0.0204, 0.2116]$ |
| da Silva et al. (2019) [38] | 0.024 | $0.024 \pm 1.96 * 0.0239 = [-0.0228, 0.0708]$ |
| de Bont et al. (2015) [52] | 0.029 | $0.029 \pm 1.96 * 0.0139 = [0.0018, 0.0562]$ |
| Holowka et al. (2014) [39] | 0.853 | $0.853 \pm 1.96 * 0.011 = [0.8314, 0.8746]$ |
| Lommen and Restifo (2009) [54] | 0 | $0.0 \pm 1.96 * 0.0 = [0, 0]$ |
| **Marx et al. (2017) [55]** | 0.828 | $0.828 \pm 1.96 * 0.0157 = [0.7972, 0.8588]$ |
| **Schwartz et al. (2005) [56]** | 0.115 | $0.115 \pm 1.96 * 0.0209 = [0.074, 0.156]$ |
| **Wang and Vivek (2013) [57]** | 0.211 | $0.211 \pm 1.96 * 0.0662 = [0.0812, 0.3408]$ |
| **van Zyl et al. (2008) [43]** | 0.0 | $0.0 \pm 1.96 * 0.0 = [0, 0]$ |
| **Zammit et al. (2018) [25]** | 0.115 | $0.115 \pm 1.96 * 0.0069 = [0.1015, 0.1285]$ |
| **Reynolds et al. (2005) [42]** | 0.05 | $0.05 \pm 1.96 * 0.0487 = [-0.046, 0.146]$ |
| **Lewis et al. (2017) [41]** | 0.386 | $0.386 \pm 1.96 * 0.0233 = [0.340, 0.432]$ |
| **Meltzer et al. (2012) [58]** | 0.11 | $0.11 \pm 1.96 * 0.0271 = [0.0569, 0.1631]$ |
| **Kostaras et al. (2016) [40]** | 0.282 | $0.282 \pm 1.96 * 0.0721 = [0.1407, 0.4233]$ |
| **Gravely et al. (2011) [23] – Specialized Care** | 0.794 | $0.794 \pm 1.96 * 0.0088 = [0.7768, 0.8112]$ |
| **Gravely et al. (2011) [23] – Primary Care** | 0.693 | $0.693 \pm 1.96 * 0.0122 = [0.6691, 0.7169]$ |
| **Magruder et al. (2008) [59]** | 0.43 | $0.43 \pm 1.96 * 0.05 = [0.332, 0.528]$ |
| **Ivanov et al. (2012) [60]** | 0.375 | $0.375 \pm 1.96 * 0.0605 = [0.2564, 0.4936]$ |
| **Tiet et al. (2013) [61]** | 0.565 | $0.565 \pm 1.96 * 0.0364 = [0.4937, 0.6363]$ |
| **Bohnert et al. (2016) [24] - Combined Specialized Care** | 0.644 | $0.644 \pm 1.96 * 0.0084 = [0.6275, 0.6605]$ |
| **Bohnert et al. (2016) [24] - Primary Care** | 0.479 | $0.479 \pm 1.96 * 0.00371 = [0.472, 0.486]$ |
| **Meltzer-Brody et al. (2004) [62]** | 0.12 | $0.12 \pm 1.96 * 0.0650 = [-0.0074, 0.2474]$ |
| **Prins et al. (2003) [29]** | 0.391 | $0.391 \pm 1.96 * 0.0922 = [0.2103, 0.5717]$ |
| **Taubman-Ben-Ari et al. (2001) [30]** | 0.024 | $0.024 \pm 1.96 * 0.0097 = [0.0050, 0.0430]$ |
| **Graves et al. (2011) [31]** | 0.308 | $0.308 \pm 1.96 * 0.0484 = [0.2131, 0.4029]$ |
| **Liebschutz et al. (2007) [32]** | 0.111 | $0.111 \pm 1.96 * 0.0290 = [0.0542, 0.1678]$ |
| **Lu et al. (2023) [63]** | 0.239 | $0.239 \pm 1.96 * 0.0521 = [0.1369, 0.3411]$ |
| **Carey et al. (2003) [33]** | 0.0 | $0.0 \pm 1.96 * 0.0 = [0, 0]$ |
| **Seal et al. (2008) [64]** | 0.24 | $0.24 \pm 1.96 * 0.0326 = [0.1761, 0.3039]$ |
| **Kimerling et al. (2006) [34]** | 0.382 | $0.382 \pm 1.96 * 0.0833 = [0.219, 0.545]$ |
| **McGuire et al. (2022) [66]** | 0.227 | $0.227 \pm 1.96 * 0.0255 = [0.177, 0.277]$ |
| **McKenzie and Smith (2006) [65]** | 0.74 | $0.74 \pm 1.96 * 0.0353 = [0.6708, 0.8092]$ |

These study-level CIs were calculated using the normal approximation formula for descriptive purposes and may differ from the CIs displayed in the forest plots, which were derived from the random-effects meta-analysis model with logit transformation and inverse-variance weighting.

Random-effects meta-analysis: For each group of studies, a random-effects meta-analysis of proportions was conducted using logit transformation and inverse-variance weighting to estimate pooled diagnostic accuracy and corresponding 95% CIs. Between-study heterogeneity was quantified using the I² and τ² statistics, where I² represents the proportion of total variation due to heterogeneity rather than chance, and τ² estimates the between-study variance in effect sizes. Pooled estimates and forest plots were generated in R version 4.2.0 [80] using the meta package [81] and the metaprop() function. Between-study variance (τ²) was estimated using the DerSimonian–Laird method, the default estimator in metaprop() unless otherwise specified.

Plotting forest plots: Forest plots were generated to visualize study-level diagnostic accuracy rates, their 95% CIs, and the pooled estimate with heterogeneity statistics.

For Gravely et al. (2011) [23], which reported diagnostic accuracy as the PPV based on PTSD administrative codes and PCL screening, the same SE formula was applied using the number of participants with PTSD diagnoses in administrative records n_diagnosed PTSD_ as the denominator instead of n_(true PTSD).

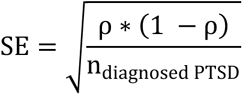

However, because PPV is not directly comparable to sensitivity measures, this study was excluded from all pooled meta-analyses and forest plots.

For studies in Group 3, which evaluated clinicians’ diagnostic accuracy under controlled or hypothetical conditions, the SEs were calculated using the number of clinician participants as the denominator. These studies assessed clinician decision-making accuracy in response to case vignettes or simulated patients rather than actual patient populations. The SE formula was adapted as follows:

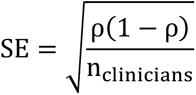

where p denotes the reported diagnostic accuracy rate and n_clinicians_ is the number of clinicians evaluated in the study.

Fig. 2, displayed as a forest plot, illustrates the proportion of individuals accurately diagnosed with PTSD across studies conducted in specialized MH settings (Group 1). Estimates were pooled using a random-effects meta-analysis model. Each box represents the sensitivity estimate, while the whiskers denote the corresponding 95% CI. Holowka et al. [39] reported the highest sensitivity (0.853), while Lommen and Restifo [54] and van Zyl et al. [43] reported the lowest (0.00).

**Fig 2.**
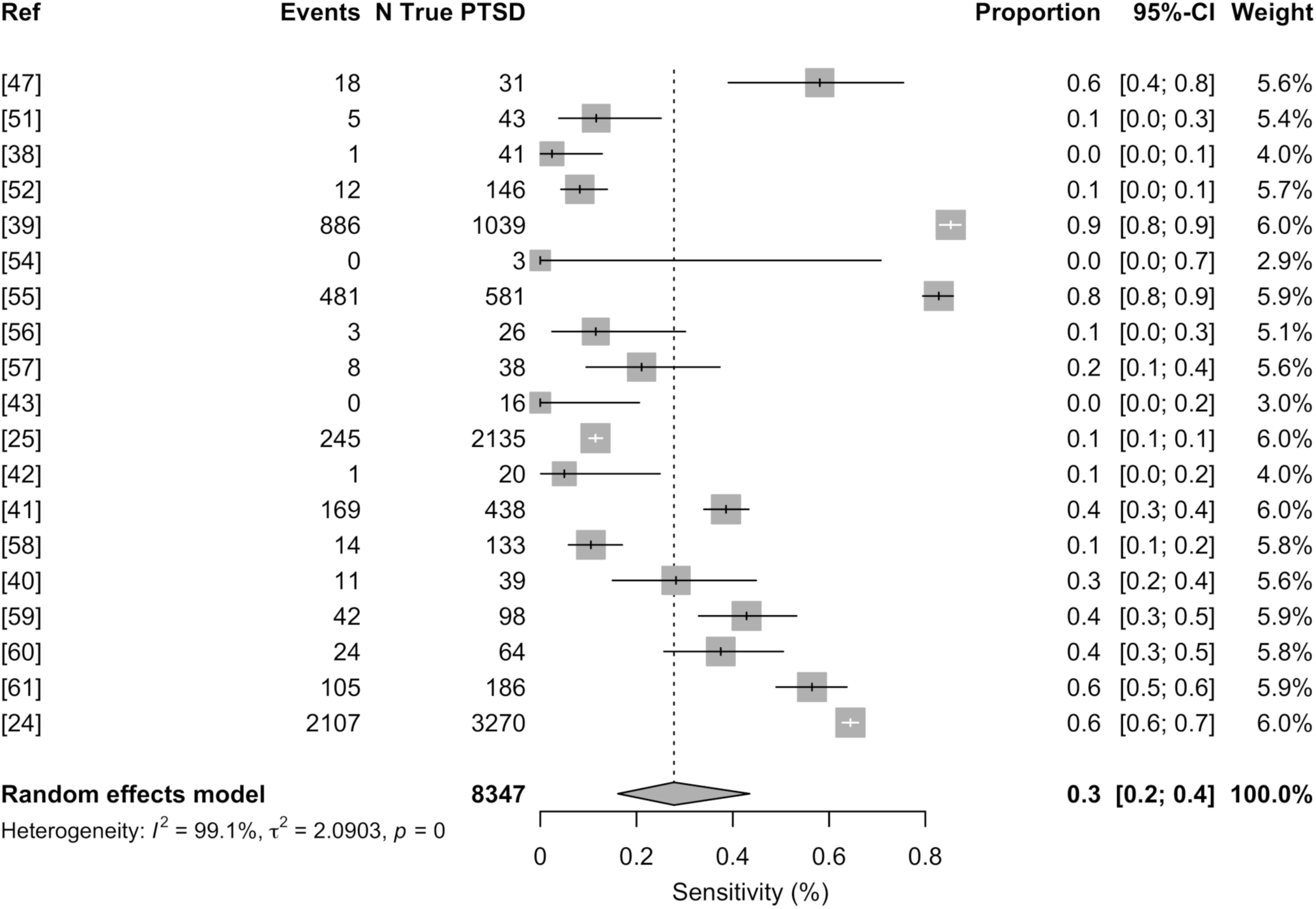
PTSD diagnostic accuracy/recognition rate in mental health settings. PTSD = post-traumatic stress disorder.

Fig. 3, also shown as a forest plot, presents the proportion of individuals accurately diagnosed with PTSD across studies conducted in primary care settings (Group 2). Estimates were pooled using a random-effects meta-analysis model. As in Fig. 2, boxes represent sensitivity estimates, and whiskers denote the 95% CI. Gravely et al. [23] was excluded from this analysis because it reported positive predictive values rather than sensitivity. Among included studies, Bohnert et al. [24] reported the highest sensitivity (0.48), while Carey et al. [33] reported the lowest (0.00).

**Fig 3.**
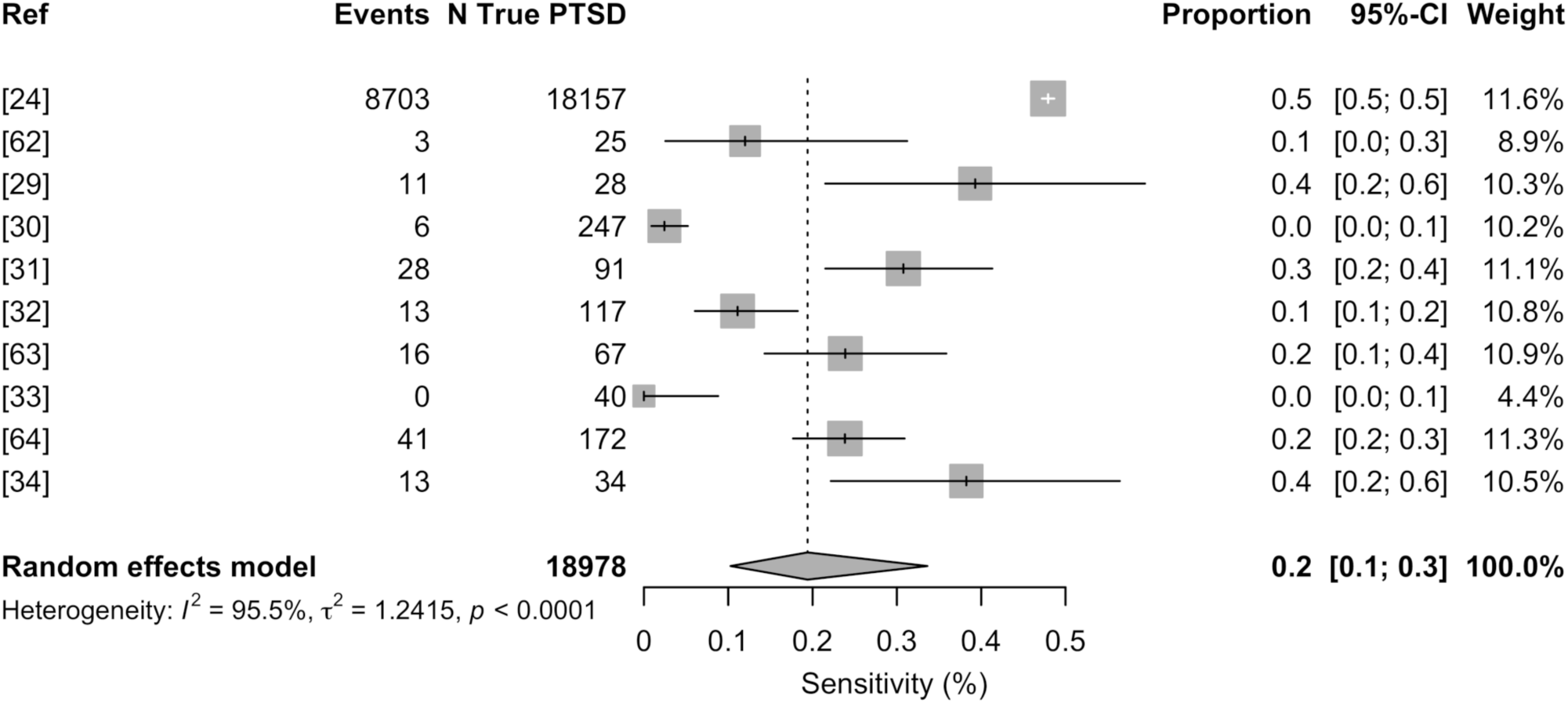
PTSD diagnostic accuracy/recognition rate in primary care settings.

Fig. 4 displays forest plots of sensitivity estimates and 95% CIs from 2 studies in Group 3. McKenzie and Smith [65] assessed clinicians’ ability to accurately identify PTSD based on knowledge and training, while McGuire et al. [66] evaluated the influence of diagnostic expectation biases during simulated diagnostic decision-making. Due to the inclusion of only 2 studies, heterogeneity statistics should be interpreted with caution.

**Fig 4.**
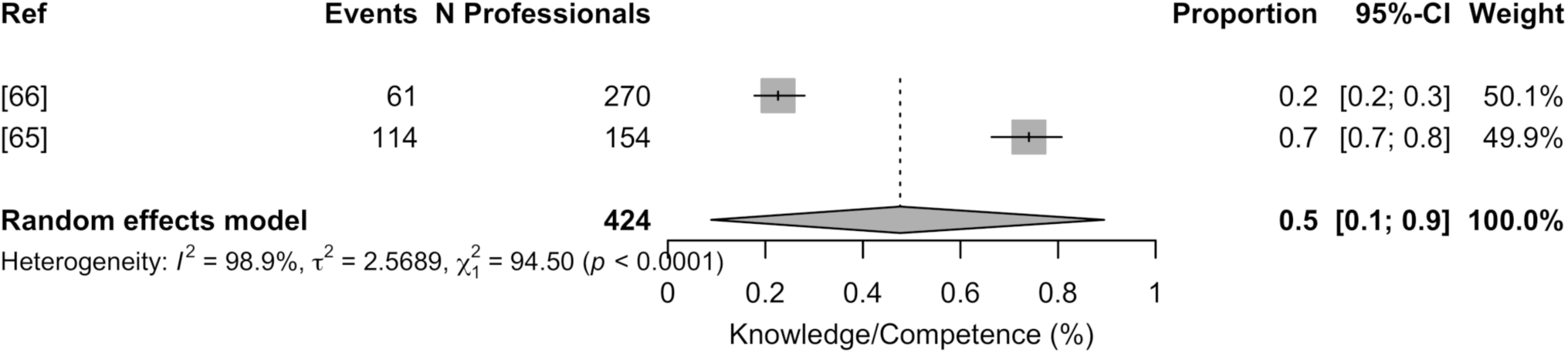
Clinician PTSD diagnostic competence and bias resistance.

#### Sensitivity and Heterogeneity Analyses

To evaluate the robustness of the synthesized results and address potential sources of methodological heterogeneity, both descriptive and model-based meta-analyses were conducted. A quantitative analysis was performed using weighted means and standard error calculations across the subset of studies reporting sufficient statistical data. One study [23] reporting diagnostic accuracy solely as a positive predictive value (PPV) was included in the descriptive analysis due to available contextual data but was excluded from the random-effects meta-analysis to avoid heterogeneity introduced by incompatible outcome measures.

In studies reporting diagnostic estimates for both primary care and specialized mental health settings [23, 24], setting-specific results were included in both corresponding subgroups. Studies were thus grouped by clinical context to allow exploration of heterogeneity across diagnostic environments. A distinct group of studies was also identified based on whether they assessed clinician-level diagnostic behavior or bias, and these were analyzed separately to evaluate the potential impact of provider-related factors.

Consistency between the descriptive and model-based pooled estimates supported the reliability of the findings and suggested that methodological stratification reduced unexplained heterogeneity. Residual heterogeneity is expected given the limited number of studies reporting PTSD diagnostic accuracy in real-world settings, but appropriate statistical models and subgroup stratification reduced its influence. Sixteen studies that lacked sufficient statistical data (e.g., numerators, denominators, or structured diagnostic outcomes) were excluded from the quantitative synthesis but retained in the qualitative review. Their exclusion minimized potential bias due to incomplete data and reinforced the methodological integrity of the pooled analysis.

#### Certainty of Evidence

The overall certainty in the quantitatively pooled diagnostic accuracy estimates was judged to be moderate to low, based on the GRADE [45] criteria. This assessment was based on several limiting factors, including substantial between-study heterogeneity (as indicated by I² and τ²), methodological variation across studies, and variability in diagnostic reference standards and clinical settings. Although many studies used validated diagnostic tools, others relied on administrative records or screening measures, reducing comparability. Only studies providing sufficient data for meta-analysis were included in this certainty assessment. To ensure consistency, outcome definitions were carefully reviewed, and where multiple diagnostic accuracy measures were reported, data corresponding to sensitivity were extracted whenever possible. Pooled estimates (both descriptive and model-based) were derived exclusively from sensitivity data, with one exception: a single study reporting only positive predictive value (PPV) [23] was excluded from the meta-analysis but included in the descriptive synthesis due to contextual relevance. Stratification by setting and methodological focus was used to explore and partially mitigate remaining sources of variation. However, the limited number of studies within certain subgroups (e.g., Group 3) and wide confidence intervals in several small-sample studies further lowered confidence in the pooled estimates.

## Discussion

### Consequences of PTSD misdiagnosis

The misdiagnosis of PTSD carries serious consequences for individual outcomes, healthcare efficiency, and broader access to MH services. Individuals who are misdiagnosed often receive inappropriate treatment, such as antidepressants for presumed depression, while the underlying trauma remains unaddressed [32, 58]. This can lead to prolonged psychological distress, exacerbation of symptoms, and an increased risk of secondary conditions such as substance use disorders, suicidal ideation, and social withdrawal [31]. Additionally, the misdiagnosis of PTSD contributes to significant economic strain by increasing the frequency of healthcare visits, inappropriate psychiatric medication use, and extended therapy sessions that do not address the primary disorder [68, 78].

### PTSD diagnostic accuracy in specialized MH

The meta-analysis findings indicate that PTSD diagnostic accuracy in specialized MH settings is higher than in primary care but it remains variable. A weighted mean diagnostic accuracy of 55.6% in these settings suggests that even trained MH professionals may face challenges in correctly identifying PTSD [68]. Contributing factors may include reliance on patient self-reporting, inconsistencies in clinician training, and symptom overlap with other psychiatric conditions [25, 39]. Further, although structured diagnostic instruments for PTSD are available, they are frequently underutilized due to time constraints and the demands of clinician workloads [41].

### PTSD diagnostic accuracy in primary healthcare

Primary healthcare settings show an even lower PTSD diagnostic accuracy rate, with a weighted mean of 48.2%. PTSD remains substantially underdiagnosed due to the absence of structured screening protocols and limited clinician training in trauma assessment [26, 31, 32]. Many PCPs misattribute PTSD symptoms to depression, anxiety, or somatic complaints, resulting in delayed or inappropriate treatment [62]. Additionally, underreporting by individuals—particularly those unaware of the psychological impact of trauma exposure further complicates diagnostic accuracy [42, 62].

### The impact of clinician training deficiencies

A key finding across studies is the lack of standardized training in assessing traumatic exposure and PTSD symptomatology among both general and MH professionals. Although structured diagnostic interviews such as CAPS-5 [2] and SCID are considered the gold-standard, they are rarely implemented in routine clinical practice due to time constraints and limited familiarity among clinicians [39]. Many healthcare providers receive insufficient training in distinguishing PTSD symptoms from those of frequently comorbid disorders, which further contributes to misdiagnosis and underdiagnosis [25, 41]. Nakash et al. [79] found that during real-world intake sessions, clinicians frequently failed to gather the information necessary to apply DSM diagnostic criteria accurately, which contributed to diagnostic inconsistency and under-recognition of PTSD. Quintana et al. [82] reported a 26.9% misclassification rate when comparing the CIDI to the SCID, noting especially low sensitivity for avoidance and impairment symptoms, which are critical for a PTSD diagnosis. These findings underscore how structured interviews, while essential for diagnostic accuracy, are often bypassed in real-world clinical care due to resource limitations. To address these barriers, several studies advocate for using brief, validated screeners. For example, the PC-PTSD-5 has demonstrated excellent diagnostic accuracy in primary care settings, with area under the curve values exceeding 0.93 in validation studies [83, 84]. The integration of such tools into routine care offers a promising solution to improve detection while accommodating real-world time constraints.

### Major points of agreement

Several key areas of agreement emerge across the reviewed studies:

1. **PTSD is substantially underdiagnosed in clinical practice.** All included studies consistently found that many individuals who met the criteria for PTSD based on structured diagnostic tools were not recognized as such in clinical records. Recognition rates were particularly low in public psychiatric settings and among individuals with severe mental illness or substance use disorders. For example, Lommen and Restifo [54] reported a 0% recognition rate in a Dutch sample of people with psychosis, while da Silva et al. [38] found a 2.4% rate in a Brazilian outpatient clinic, despite much higher rates based on structured assessments.
2. **Structured diagnostic tools consistently reveal higher PTSD rates.** All studies that compared structured tools (e.g., CAPS, MINI, SCID) or validated screeners (e.g., TSQ, PCL) to routine diagnoses found that the structured assessments identified significantly more PTSD cases. For instance, de Bont et al. [52] found that 32% of participants met the PTSD criteria on the CAPS, but most were clinically undiagnosed.
3. **Comorbid psychiatric conditions can mask PTSD symptoms:** Many studies have found that PTSD symptoms are often misattributed to more visible or stigmatized diagnoses, such as depression, psychosis, or substance use disorders. This was highlighted in Reynolds et al. [42], where PTSD was underdiagnosed among patients with substance use disorders, and in Lommen and Restifo [54], where psychotic symptoms obscured PTSD recognition.
4. **Lack of clinician training contributes to missed diagnoses:** Several studies have noted that many clinicians, especially in general psychiatric or primary care settings, lack sufficient training to recognize trauma-related symptoms. This contributes to frequent misdiagnoses or the omission of PTSD from diagnostic formulations [55, 66].
5. **Importance of comprehensive assessments:** The use of structured clinical interviews such as CAPS-5 [2] and SCID is widely recognized as critical to improving diagnostic accuracy. However, their routine implementation remains limited due to time constraints and deficits in clinician training [25, 41, 43]. Nakash et al. [79] demonstrated that clinicians often miss key diagnostic criteria during unstructured intake assessments.
6. **Comorbid psychiatric disorders often obscure PTSD:** In many clinical settings, PTSD is overshadowed by co-occurring diagnoses such as depression, psychosis, or substance use disorders, which are more readily identified or prioritized in treatment planning [40, 42, 54].
7. **Clinicians often do not inquire about trauma:** Multiple studies have found that the failure to systematically assess trauma history contributes to underdiagnosis. Even when trauma was clearly present in a patient’s history, it was frequently undocumented or unexplored [43, 57].
8. **PTSD is especially likely to be overlooked in vulnerable or high-risk populations:** The presentation of PTSD symptom varies across cultural contexts, highlighting the need for culturally sensitive diagnostic approaches to improve detection in diverse populations [31]. Individuals with psychotic disorders, substance use disorders, and those from marginalized groups were particularly likely to have PTSD go undetected, despite high rates of trauma exposure [47, 52].

### Major points of disagreement

Despite a broad consensus on the underdiagnosis of PTSD, several areas of disagreement remain:

1. **Extent of Underdiagnosis Across Settings:** While all studies agreed that PTSD is underdiagnosed, they varied considerably in the degree to which this underdiagnosis occurs. For example, some studies reported complete diagnostic failure (e.g., Lommen & Restifo [54] with 0% recognition), while others found partial recognition, with up to half of PTSD cases being identified depending on the clinical setting, diagnostic method, and population studied (e.g., Reynolds et al. [42]). These differences suggest variability in system effectiveness, clinician expertise, and patient disclosure across contexts.
2. **Impact of Comorbid Depression on PTSD Recognition:** There was inconsistency in how comorbid depression affected PTSD diagnosis. In some studies (e.g., Meltzer et al. [58]), depression increased the likelihood of PTSD being recognized due to symptom overlap. In contrast, others (e.g., Kostaras et al. [40]; Reynolds et al. [42]) found that depressive symptoms often masked PTSD, leading to diagnostic overshadowing.
3. **Utility and Interpretation of Self-Report vs. Clinician-Administered Tools:** Disagreement exists regarding the relative effectiveness of self-report tools (e.g., PCL, TSQ) compared to structured interviews (e.g., CAPS, SCID). Some studies support brief self-report screeners for their practicality and acceptable sensitivity (de Bont et al. [52]), while others caution that they may overestimate PTSD or miss complex symptom presentations in populations with comorbidities or psychosis (Cusack et al. [51]).
4. **Influence of Clinical Setting on Diagnostic Accuracy:** Different studies have drawn contrasting conclusions about where PTSD is most likely to be identified. For example, Meltzer et al. [58] found that PTSD was more likely to be diagnosed in primary care settings, potentially due to overlapping depression screening. In contrast, Van Zyl et al. [43] found zero recognition of PTSD in psychiatric outpatient clinics despite high rates reported in structured interviews, suggesting inconsistent practices across settings.
5. **Effectiveness of Screening in Routine Practice:** While most studies supported brief screening tools, there was disagreement about their standalone reliability in real-world settings. Some authors advocated for the routine use of tools like the PC-PTSD and TSQ (e.g., Prins et al. [29]; de Bont et al. [52]), while others warned that without follow-up assessment or adequate training, such tools might produce false positives or underdiagnosis (e.g., Lewis et al. [41]).
6. **Cultural and Contextual Interpretations of PTSD:** There was some disagreement among the included studies regarding the necessity of cultural adaptation in PTSD diagnostics. McGuire et al. [66] emphasized the importance of culturally sensitive diagnostic approaches, noting that symptom presentation and help-seeking behaviors vary across populations and that standard tools may overlook trauma expressions shaped by culture. In contrast, Lewis et al. [41], Reynolds et al. [42], and Van Zyl et al. [43] applied standardized diagnostic tools (e.g., PCL-C, SCID, CAPS) across diverse or non-Western populations, including immigrant communities, substance use clinics, and South African psychiatric settings, without significant adaptation or discussion of cultural limitations. These studies did not report poor performance of the tools due to cultural mismatch, implying that standard instruments were considered sufficient or at the very least, functionally acceptable within those contexts.

### Unanswered questions and future directions

While this review identified several consistent challenges in PTSD diagnosis across clinical settings, it also highlighted important gaps that remain unresolved both within and beyond the current evidence base:

1. **Genetic and biological contributions:** Despite growing interest, few studies have integrated biological mechanisms into PTSD diagnostic research. Genetic variants in *SLC6A4*, *FKBP5*, and *ADCYAP1R1* have been associated with 20–40% of PTSD heritability [85, 86]. Recent studies have expanded on this by identifying epigenetic markers such as CpG methylation in genes like *NRG1*, *HGS*, and *AHRR*, as well as altered miRNA expression patterns involved in stress regulation [87, 88, 89]. These biomarkers hold promise for distinguishing PTSD from comorbid psychiatric conditions and enabling precision diagnostics. Future work should integrate genomic, epigenetic, and transcriptomic signatures into diagnostic frameworks to enhance personalized care.
2. **Socioeconomic and structural inequities:** Poverty, systemic racism, limited access to MH, and insurance instability remain major drivers of the underdiagnosis of PTSD. African American adults [31], veterans without VA access [90], and people experiencing homelessness [91] consistently face structural barriers to diagnosis and treatment. Further, neurobiological responses to trauma may be shaped by lived experiences of inequality, as evidenced by altered amygdala connectivity patterns in marginalized racial groups [92]. Women exposed to intimate partner violence are another underdiagnosed population, due to both social stigma and lack of screening infrastructure [93]. Research should prioritize trauma-informed models of care that actively address social determinants of health and correct structural inequities in access.
3. **Longitudinal impact of diagnostic errors:** Most studies rely on cross-sectional data, leaving the long-term effects of PTSD misdiagnosis underexplored. However, longitudinal research shows that undiagnosed or misdiagnosed PTSD can result in persistent symptoms, reduced quality of life, increased suicide risk, and worsened physical health outcomes over time [94, 95, 96, 97, 98]. For example, unrecognized PTSD in general psychiatric settings has been associated with poorer treatment outcomes and higher comorbidity rates [99]. Future research should track the chronic impact of diagnostic failures and evaluate interventions that promote long-term clinical recovery.
4. **Innovation through artificial intelligence (AI):** AI-based systems are being developed to improve PTSD screening by analyzing electronic health records, voice features, and self-reported outcomes. These tools could enhance early detection in primary care and underserved settings [93]. However, ethical, logistical, and technical challenges remain. Studies have warned about algorithmic bias, inadequate cultural adaptation, and lack of transparency in AI-based tools [100, 101]. Additionally, many systems are trained on non-representative datasets, which risks poor generalizability. Future development should emphasize explainable AI, equitable access, and robust validation across diverse populations.
5. **Cultural adaptation and global relevance:** Current diagnostic tools may fail to capture trauma-related distress in culturally diverse populations. In many non-Western settings, symptoms are expressed through culturally specific idioms of distress that do not align with DSM-5 criteria. This mismatch can lead to underdiagnosis and inappropriate treatment [64, 92]. Research should focus on validating screening instruments in diverse cultural contexts and integrating community-based concepts of trauma and resilience into diagnostic frameworks.
6. **Gaps in healthcare infrastructure:** In rural and underserved regions, limited MH infrastructure, broadband connectivity, and clinician shortages impede access to PTSD diagnoses. Telepsychiatry and collaborative care models have improved access and outcomes in some settings [78, 79], but implementation gaps persist. Studies emphasize the need for scalable training programs for primary care providers, investment in telehealth infrastructure, and adaptations for low-resource environments.
7. **Ethical and Privacy Concerns in Digital Tools:** As digital tools are integrated into PTSD care, ethical challenges such as data privacy, consent, and algorithmic accountability should be addressed. AI systems often lack transparency and may reinforce biases when trained on non-inclusive datasets [100, 102]. Concerns also include unclear data governance, insufficient patient consent mechanisms, and lack of human oversight. Researchers have called for interdisciplinary frameworks that ensure the fairness, explainability, and safety of digital diagnostics.

### Contributions to the global literature

This systematic review synthesizes evidence on the diagnostic accuracy of PTSD tools across both primary care and specialized MH settings, highlighting marked variability in accuracy depending on the care context, population, and instrument used. It identifies systemic barriers to accurate PTSD identification, including inconsistent screening application, limited clinician training, and inequities in access to MH evaluation. By documenting diagnostic disparities—particularly under-recognition in general medical settings and among marginalized populations—this review underscores the importance of integrating standardized, trauma-informed screening protocols into routine care. Although AI tools are not yet widely implemented in current studies, this review also outlines their future potential to support equitable and efficient PTSD diagnosis. Together, these findings contribute to a structured evidence base that informs diagnostic reform and guides the development of more inclusive and context-sensitive screening practices globally.

### Limitations and future directions

Despite efforts to ensure methodological rigor, this study has some limitations. First, the review process was primarily conducted by a single author, which may have introduced selection or interpretation bias and limited inter-rater reliability [103]. Second, by excluding studies that did not report formal diagnostic accuracy metrics, the review may have missed valuable insights into the implementation challenges and contextual factors affecting PTSD underdiagnosis [104]. Third, heterogeneity in study populations, diagnostic instruments, and outcome definitions precluded direct comparisons and limited opportunities for meta-regression analysis [105]. Although random-effects meta-analyses were used to account for between-study variability, substantial heterogeneity remained, and pooled estimates should be interpreted cautiously, particularly in subgroups with few included studies. This heterogeneity was largely unavoidable, as only a limited number of studies reported formal diagnostic accuracy metrics in real-world clinical contexts. Such variation reflects the diversity of PTSD diagnostic practice across settings, instruments, and populations. To minimize its impact, random-effects models, subgroup analyses, and sensitivity analyses were applied, which improved consistency between descriptive and pooled estimates. Publication bias may also be present, as studies with null or unfavorable diagnostic findings are often less likely to be published or indexed [106]. Detailed limitations of the included studies are documented in the supplemental material (Table S2).

These limitations highlight important directions for future research. PTSD screening tools should be validated in diverse real-world care settings, particularly those with limited resources [107]. There is also a need to develop culturally adapted instruments that reflect local idioms of distress and improve diagnostic equity among underserved populations [108]. As AI and digital technologies continue to expand into psychiatric care, their use in PTSD diagnosis should be guided by rigorous evaluation and ethical safeguards to ensure transparency, accuracy, and fairness [109]. Future systematic reviews should consider broader inclusion criteria, dual-reviewer screening, and mixed-method synthesis to capture a more comprehensive picture of diagnostic challenges and innovations.

## Supporting information

PRISMA Checklist 1

PRISMA Checklist 2

Initial Review PRISMA Flow Diagram

R Code Mental Health Settings Meta-analysis

R Code Primary Settings Meta-analysis

R Code Clinician Bias/Knowledge Meta-analysis

Supplemental Text - Reconstructed Search Strategy

## Data Availability

All data produced in the present work are contained in the manuscript submission as suplemental files.

## Acknowledgments

Support with baseline data collection for a few included studies and supervision of the initial report were provided by Brad Strasser, RAC – Vice President of Clinical, Quality, and Regulatory Affairs at Senseye, Inc.

Taylor & Francis Editing Services (https://www.tandfeditingservices.com/) provided scientific editing of the final manuscript.

Supporting Information files include data extraction tables (S1–S3 Tables), PRISMA checklists (S4–S5), the original PRISMA flow diagram (S6), the full dataset used for meta-analysis (S7), and the complete R code used for statistical analysis and figure generation (S8–S10). A detailed search strategy is also provided (S11). All files will be available upon publication at (<u>osf.io/xzyhp</u>; osf.io/umwxd) [110, 111] for full transparency and reproducibility.

## Supporting information

**S1 Table.** Characteristics of each of the included studies

**S2 Table.** Risk of bias assessments of the included studies

**S3 Table.** IRB approvals for each of the included studies

**S4 Checklist.** PRISMA 2020 for abstracts checklist

**S5 Checklist.** PRISMA 2020 checklist

**S6 Figure.** Original PRISMA 2020 flow diagram used during the initial search phase in May–June 2023.

**S7 Dataset.** Excel file with three sheets listing included studies for: (1) specialized mental healthcare, (2) primary care, and (3) clinician diagnostic competence. Includes original Excel forest plots (not based on meta-analysis); final plots appear in main manuscript figures.

**S8 Code.** R script for random-effects meta-analysis and forest plot: specialized mental healthcare.

**S9 Code.** R script for random-effects meta-analysis and forest plot: primary care.

**S10 Code.** R script for random-effects meta-analysis and forest plot: clinician diagnostic competence.

**S11 Text.** Documentation of the initial and updated search phases, including databases, search terms, date ranges, and additional methods.

## References

1. American Psychiatric Association. Diagnostic and statistical manual of mental disorders. 5th ed. Washington, DC: American Psychiatric Publishing; 2013. doi: 10.1176/appi.books.97808904255962.

2. Weathers FW, Blake DD, Schnurr PP, Kaloupek DG, Marx BP, Keane TM. The Clinician-Administered PTSD Scale for DSM-5 (CAPS-5). 2013 [cited 2024 October 19]. Available from: https://www.ptsd.va.gov3.

3. Nordgaard J, Sass LA, Parnas J. The psychiatric interview: validity, structure, and subjectivity. Eur Arch Psychiatry Clin Neurosci. 2013;263(4): 353–364. doi: 10.1007/s00406-012-0366-z.

4. Bovin MJ, Marx BP, Weathers FW, Gallagher MW, Rodriguez P, Schnurr PP, et al. Psychometric properties of the PTSD Checklist for Diagnostic and Statistical Manual of Mental Disorders–Fifth Edition (PCL-5) in veterans. Psychol Assess. 2016;28(11): 1379–1391. doi: 10.1037/pas0000254.

5. Blake DD, Weathers FW, Nagy LM, Kaloupek DG, Gusman FD, Charney DS, et al. The development of a Clinician-Administered PTSD Scale. J Trauma Stress. 1995;8(1): 75–90.

6. First MB, Williams JBW, Karg RS, Spitzer RL. Structured Clinical Interview for DSM-5 Disorders: Clinician Version (SCID-5-CV). Washington, DC: American Psychiatric Association Publishing; 2016.

7. Weathers FW, Bovin MJ, Lee DJ, Sloan DM, Schnurr PP, Kaloupek DG, et al. The Clinician-Administered PTSD Scale for DSM-5 (CAPS-5): Development and initial psychometric evaluation in military veterans. Psychol Assess. 2018;30(3): 383–395. doi: 10.1037/pas0000486.

8. Engelhard IM, Arntz A, van den Hout MA. Low specificity of symptoms on the post-traumatic stress disorder (PTSD) symptom scale: A comparison of individuals with PTSD, individuals with other anxiety disorders, and individuals without psychopathology. Br J Clin Psychol. 2007;46(4): 449–456. doi: 10.1348/014466507X206883.

9. Shalev AY, Peri T, Canetti L, Schreiber S. Predictors of PTSD in injured trauma survivors: A prospective study. Am J Psychiatry. 1996;153(2): 219–225. doi: 10.1176/ajp.153.2.219.

10. Boscarino JA, Kirchner HL, Hoffman SN, Sartorius J, Adams RE, Figley CR. A brief screening tool for assessing psychological trauma in clinical practice: Development and validation of the New York PTSD Risk Score. Gen Hosp Psychiatry. 2011;33(5): 489–500. doi: 10.1016/j.genhosppsych.2011.06.001.

11. Dimauro J, Carter SP, Folk JB, Kashdan TB. A historical review of trauma-related diagnoses to reconsider the heterogeneity of PTSD. J Anxiety Disord. 2014;28(8): 774–786. doi: 10.1016/j.janxdis.2014.09.002.

12. Hyland P, Shevlin M, Brewin CR, Cloitre M, Downes A, Jumbe S, et al. Validation of post-traumatic stress disorder (PTSD) and complex PTSD using the International Trauma Questionnaire. Acta Psychiatr Scand. 2017;136: 313–322. doi: 10.1111/acps.12771.

13. May CL, Wisco BE. Defining trauma: How level of exposure and proximity affect risk for posttraumatic stress disorder. Psychol Trauma. 2016;8(2): 233–240. doi: 10.1037/tra0000077.

14. Center for Substance Abuse Treatment (U.S.). Exhibit 1.3-4, DSM-5 diagnostic criteria for PTSD. Trauma-Informed Care in Behavioral Health Services. Substance Abuse and the Mental Health Service Administration; [no date].

15. American Psychiatric Association. Diagnostic and statistical manual of mental disorders. 5th ed. Text rev. Washington, DC: American Psychiatric Association; 2022.

16. Hinton DE, Lewis-Fernández R. The cross-cultural validity of posttraumatic stress disorder: implications for DSM-5. Depress Anxiety. 2011;28(9): 783–801.

17. Friedman MJ. Finalizing PTSD in DSM-5: getting here from there and where to go next. J Trauma Stress. 2013;26(5): 548–556.

18. Galatzer-Levy IR, Bryant RA. 636,120 ways to have posttraumatic stress disorder. Perspect Psychol Sci. 2013;8(6): 651–662.

19. Merten JW, Cwik MF, Margraf J, Schneider S. Overreliance on symptom checklists in PTSD assessment: a review of problems and potential solutions. Clin Psychol Rev. 2016;44: 13–24.

20. Australian Government National Health and Medical Research Council. NHMRC levels of evidence and grades for recommendations for developers of guidelines. 2009 [cited 2024 October 19]. Available from: https://www.nhmrc.gov.au/sites/default/files/images/NHMRC%20Levels%20and%20Grades%20(2009).pdf

21. Page MJ, McKenzie JE, Bossuyt PM, Boutron I, Hoffmann TC, Mulrow CD, et al. The PRISMA 2020 statement: An updated guideline for reporting systematic reviews. PLOS Med. 2021;18(3): e1003583. doi: 10.1371/journal.pmed.1003583.

22. U.S. Department of Veterans Affairs. PTSD: National Center for PTSD [Internet]. Washington, DC: U.S. Department of Veterans Affairs; [cited 2025 April 20]. Available from: https://www.ptsd.va.gov/

23. Gravely AA, Cutting A, Nugent S, Grill J, Carlson K, Spoont M. Validity of PTSD diagnoses in VA administrative data: Comparison of VA administrative PTSD diagnoses to self-reported PTSD Checklist scores. J Rehabil Res Dev. 2011;48(1): 21–30. doi: 10.1682/JRRD.2009.08.0116.

24. Bohnert KM, Sripada RK, Mach J, McCarthy JF. Same-day integrated mental health care and PTSD diagnosis and treatment among VHA primary care patients with positive PTSD screens. Psychiatr Serv. 2016;67(1): 94–100. doi: 10.1176/appi.ps.201500035.

25. Zammit S, Lewis C, Dawson S, Colley H, McCann H, Piekarski A, et al. Undetected post-traumatic stress disorder in secondary-care mental health services: systematic review. Br J Psychiatry. 2018;212(1): 11–18. doi: 10.1192/bjp.2017.8.

26. Greene T, Neria Y, Gross R. Prevalence, detection and correlates of PTSD in the primary care setting: A systematic review. J Clin Psychol Med Settings. 2016;23(2): 160–180. doi: 10.1007/s10880-016-9449-8.

27. Spottswood M, Davydow DS, Huang H. The prevalence of posttraumatic stress disorder in primary care: A systematic review. Harv Rev Psychiatry. 2017;25(4): 159–169.

28. Covidence Systematic Review Software, Veritas Health Innovation. Covidence systematic review software. 2023 [cited 2024 October 19]. Available from: https://www.covidence.org

29. Prins A, Ouimette PC, Kimerling R, Cameron RP, Hugelshofer DS, Shaw-Hegwer J, et al. The Primary Care PTSD Screen (PC-PTSD): Development and operating characteristics. Prim Care Psychiatry. 2003;9(1): 9–14. doi: 10.1185/135525703125002360.

30. Taubman-Ben-Ari O, Rabinowitz J, Feldman D, Vaturi R. Post-traumatic stress disorder in primary-care settings: prevalence and physicians’ detection. Psychol Med. 2001;31(3): 555–560. doi: 10.1017/s0033291701003658.

31. Graves R, Freedy JR, Aigbogun NU, Lawson WB, Mellman TA, Alim TN. PTSD treatment of African American adults in primary care: the gap between current practice and evidence-based treatment guidelines. J Natl Med Assoc. 2011;103(7): 585–593. doi: 10.1016/s0027-9684(15)30384-9.

32. Liebschutz J, Saitz R, Brower V, Keane TM, Lloyd-Travaglini C, Averbuch T, et al. PTSD in urban primary care: high prevalence and low physician recognition. J Gen Intern Med. 2007;22(6): 719–726. doi: 10.1007/s11606-007-0161-0.

33. Carey PD, Stein DJ, Zungu-Dirwayi N, Seedat S. Trauma and posttraumatic stress disorder in an urban Xhosa primary care population: prevalence, comorbidity, and service use patterns. J Nerv Ment Dis. 2003;191(4): 230–236. doi: 10.1097/01.NMD.0000061143.66146.A8.

34. Kimerling R, Ouimette P, Prins A, Nisco P, Lawler C, Cronkite R, et al. Brief report: utility of a short screening scale for DSM-IV PTSD in primary care. J Gen Intern Med. 2006;21(1): 65–67. doi: 10.1111/j.1525-1497.2005.00292.x.

35. Neria Y, Olfson M, Gameroff MJ, et al. Trauma exposure and posttraumatic stress disorder among primary care patients with anxiety disorders. Psychiatr Serv. 2008;59(4): 460–465.

36. Gillock KL, Zayfert C, Hegel MT, Ferguson RJ. Posttraumatic stress disorder in primary care: prevalence and relationships with physical symptoms and medical utilization. Gen Hosp Psychiatry. 2005;27(6): 392–399. doi: 10.1016/j.genhosppsych.2005.09.004.

37. EndNote. EndNote: the best citation & reference management tool. 2024 [cited 2024 October 19]. Available from: https://endnote.com/

38. da Silva HC, Furtado da Rosa MM, Berger W, Luz MP, Mendlowicz M, Coutinho ESF, et al. PTSD in mental health outpatient settings: highly prevalent and under-recognized. Rev Bras Psiquiatr. 2019;41(3): 213–217. doi: 10.1590/1516-4446-2017-0025.

39. Holowka DW, Marx BP, Gates MA, Litman HJ, Ranganathan G, Rosen RC, et al. PTSD diagnostic validity in Veterans Affairs electronic records of Iraq and Afghanistan veterans. J Consult Clin Psychol. 2014;82(4): 569–579. doi: 10.1037/a0036347.

40. Kostaras P, Bergiannaki JD, Psarros C, Ploumbidis D, Papageorgiou C. Posttraumatic stress disorder in outpatients with depression: still a missed diagnosis. J Trauma Dissociation. 2016;18(2): 233–247. doi: 10.1080/15299732.2016.1237402.

41. Lewis C, Raisanen L, Bisson JI, Jones I, Zammit S. Trauma exposure and undetected posttraumatic stress disorder among adults with a mental disorder. Depress Anxiety. 2017;35(2): 178–184. doi: 10.1002/da.22707.

42. Reynolds M, Mezey G, Chapman M, Wheeler M, Drummond C, Baldacchino A. Co-morbid post-traumatic stress disorder in a substance misusing clinical population. Drug Alcohol Depend. 2005;77(3): 251–258. doi: 10.1016/j.drugalcdep.2004.08.017.

43. van Zyl M, Oosthuizen PP, Seedat S. Post traumatic stress disorder: undiagnosed cases in a tertiary inpatient setting. Afr J Psychiatry. 2008;11(2): 119–122. doi: 10.4314/ajpsy.v11i2.30263.

44. Sterne J.A.C., Higgins J.P.T., Reeves B.C., et al. (2024). ROBINS-I V2: Risk Of Bias In Non-randomized Studies – of Interventions (Version 2). Cochrane Methods Group. Available from: https://www.riskofbias.info/welcome/robins-i-v2?utm_source=chatgpt.com

45. Guyatt GH, Oxman AD, Vist GE, Kunz R, Brożek J, Alonso-Coello P, et al. GRADE guidelines: 4. Rating the quality of evidence—study limitations (risk of bias). J Clin Epidemiol. 2011;64(4): 407–415. doi: 10.1016/j.jclinepi.2010.07.017.

46. Guyatt G, Oxman AD, Montori V, Vist G, Kunz R, Brozek J, et al. GRADE guidelines: 5. Rating the quality of evidence—publication bias. J Clin Epidemiol. 2011;64(12): 1277–1282. doi: 10.1016/j.jclinepi.2011.01.011.

47. Bonn-Miller MO, Bucossi MM, Trafton JA. The underdiagnosis of cannabis use disorders and other Axis-I disorders among military veterans within VHA. Mil Med. 2012;177(7): 786–788. doi: 10.7205/milmed-d-12-00052.

48. First MB, Spitzer RL, Gibbon M, Williams JBW. Structured Clinical Interview for DSM-IV Axis I Disorders, Clinician Version (SCID-CV). Washington, DC: American Psychiatric Press; 1996.

49. American Psychiatric Association. Diagnostic and statistical manual of mental disorders. 4th ed. Washington, DC: American Psychiatric Publishing; 1994.

50. U.S. Department of Veteran Affairs. Veterans Affairs Health Care System. Veterans Health Administration. [cited 2024 October 19]. Available from: https://www.va.gov/health/

51. Cusack KJ, Grubaugh AL, Knapp RG, Frueh BC. Unrecognized trauma and PTSD among public mental health consumers with chronic and severe mental illness. Community Ment Health J. 2006;42(5): 487–500. doi: 10.1007/s10597-006-9049-4.

52. de Bont PAJM, van den Berg DPG, van der Vleugel BM, de Roos C, de Jongh A, van der Gaag M, et al. Predictive validity of the Trauma Screening Questionnaire in detecting post-traumatic stress disorder in patients with psychotic disorders. Br J Psychiatry. 2015;206(5): 408–416. doi: 10.1192/bjp.bp.114.148486.

53. U.S. Department of Veterans Affairs. Project VALOR Registry [Internet]. Washington, DC: Cooperative Studies Program Epidemiology Center; [cited 2025 April 20]. Available from: https://www.vacsp.research.va.gov/CSPEC/Studies/INVESTD-R/Project-VALOR-Registry.asp

54. Lommen MJJ, Restifo K. Trauma and posttraumatic stress disorder (PTSD) in patients with schizophrenia or schizoaffective disorder. Community Ment Health J. 2009;45(6): 485–496. doi: 10.1007/s10597-009-9248-x.

55. Marx BP, Engel-Rebitzer E, Bovin MJ, Parker-Guilbert KS, Moshier S, Barretto K, et al. The influence of veteran race and psychometric testing on veterans affairs posttraumatic stress disorder (PTSD) disability exam outcomes. Psychol Assess. 2017;29(6): 710–719. doi: 10.1037/pas0000378.

56. Schwartz AC, Bradley RL, Sexton M, Sherry A, Ressler KJ. Posttraumatic stress disorder among African Americans in an inner city mental health clinic. Psychiatr Serv. 2005;56(2): 212–215. doi: 10.1176/appi.ps.56.2.212.

57. Wang B, Vivek S. Survey of posttraumatic stress disorder (PTSD) with PTSD Checklist–Civilian (PCL-C) Questionnaire on outpatients at two mental health clinics in New York City. J Depress Anxiety. 2013;4: 7.

58. Meltzer EC, Averbuch T, Samet JH, Saitz R, Jabbar K, Lloyd-Travaglini C, et al. Discrepancy in diagnosis and treatment of post-traumatic stress disorder (PTSD): treatment for the wrong reason. J Behav Health Serv Res. 2012;39(2): 190–201. doi: 10.1007/s11414-011-9263-x.

59. Magruder KM, Frueh BC, Knapp RG, Davis L, Hamner MB, Martin RH, et al. Prevalence of posttraumatic stress disorder in Veterans Affairs primary care clinics. Gen Hosp Psychiatry. 2008;27(3): 169–179. doi: 10.1016/j.genhosppsych.2004.11.001.

60. Ivanov I, Yehuda R, Silverman JM, Siever LJ. Clinical and diagnostic characteristics of trauma-exposed patients in a psychiatric emergency setting: a preliminary report. J Trauma Dissociation. 2012;13(2): 207–221. doi: 10.1080/15299732.2011.608780.

61. Tiet QQ, Schutte KK, Leyva YE, Wierwille LA. Diagnostic accuracy of brief PTSD screening instruments in substance use disorder patients. J Subst Abuse Treat. 2013 August;45(2): 134–142. doi: 10.1016/j.jsat.2013.02.005.

62. Meltzer-Brody S, Hartmann K, Miller WC, Scott J, Garrett J, Davidson J. A brief screening instrument to detect posttraumatic stress disorder in outpatient gynecology. Obstet Gynecol. 2004;104(4): 770–776. doi: 10.1097/01.AOG.0000140683.43272.85.

63. Lu W, Bullock D, Ruszczyk L, Srijeyanthan J, Ettinger S, Caldwell B, et al. Positive PTSD screening in patients with HIV in urban primary care settings and its health correlates. J Psychosoc Nurs Ment Health Serv. 2023. doi: 10.3928/02793695-20231206-03.

64. Seal KH, Maguen S, Cohen B, Gima KS, Metzler TJ, Ren L, Bertenthal D, Marmar CR. Getting beyond “Don’t ask; don’t tell”: an evaluation of US Veterans Administration postdeployment mental health screening of veterans returning from Iraq and Afghanistan. Am J Public Health. 2011;101(5): 873–878. doi: 10.2105/AJPH.2010.300027.

65. McKenzie KJ, Smith DI. Posttraumatic stress disorder: examination of what clinicians know. Clin Psychol. 2006;10(2): 78–85. doi: 10.1080/13284200600693705.

66. McGuire R, Halligan SL, Meiser-Stedman R, Durbin L, Hiller RM. Differences in the diagnosis and treatment decisions for children in care compared to their peers: an experimental study on post-traumatic stress disorder. Br J Clin Psychol. 2022;61(4): 1075–1088. doi: 10.1111/bjc.12379.

67. Bovin MJ, Kimerling R, Weathers FW, Prins A, Marx BP, Post EP, et al. Diagnostic accuracy and acceptability of the Primary Care PTSD Screen for DSM-5 among US veterans. JAMA Netw Open. 2021;4(2): e2036733. doi: 10.1001/jamanetworkopen.2020.36733.

68. Kosowan L, Bélanger C, El-Gabalawy R. Limitations of electronic medical records in detecting PTSD: a machine learning approach. J Med Internet Res. 2022;24(7): e23456. doi: 10.2196/23456.

69. Singer JL, McCormack RM, Davis AL. PTSD diagnosis and treatment delays in primary care: a retrospective analysis. BMC Prim Care. 2024;25(1): 9–19. doi: 10.1186/s12875-023-02045-7.

70. Ehlers A, Gene-Cos N, Perrin S. Clinicians’ estimates of PTSD prevalence and recognition of trauma-related symptoms. Psychol Trauma. 2009;1(1): 34–45. doi: 10.1037/a0018856.

71. Della Porta MA. The impact of symptom presentation on PTSD detection in primary care settings. J Fam Med. 2017;66(5): 359–370. doi: 10.3122/jabfm.2017.05.170157.

72. Kaltman S, Pauk J, Alter CL. Meeting the mental health needs of low-income immigrants in primary care: a community adaptation of an evidence-based model. Am J Orthopsychiatry. 2011;81(4): 543–551.

73. Cook JM, Simiola V, McCarthy E, Ellis A, Thompson R. A brief mental health intervention for primary care patients with trauma exposure: a pilot randomized clinical trial. Gen Hosp Psychiatry. 2017;44: 58–62.

74. Bruce SE, Weisberg RB, Dolan RT, Machan JT, Kessler RC, Manchester G, et al. Trauma exposure and posttraumatic stress disorder in primary care patients. J Gen Intern Med. 2001;16(10): 625–630. doi: 10.1046/j.1525-1497.2001.016010625.x.

75. Neria Y, Gross R, Olfson M, Gameroff MJ, Wickramaratne P, Das A, et al. Posttraumatic stress disorder in primary care one year after the 9/11 attacks. Gen Hosp Psychiatry. 2006;28(3): 213–222. doi: 10.1016/j.genhosppsych.2006.02.002.

76. Sareen J, Cox BJ, Stein MB, et al. Physical and mental comorbidity, disability, and suicidal behavior associated with posttraumatic stress disorder in a large community sample. Psychosom Med. 2007;69(3): 242–248.

77. Cowlishaw S, Howard L, Dewey ME. Prevalence of probable PTSD among general practice attendees in England. J Gen Pract. 2020;70(1): 1–9. doi: 10.1017/jgp.2020.021.

78. Jiang C, Xue G, Yao S, Zhang X, Chen W, Cheng K, et al. Psychometric properties of the post-traumatic stress disorder checklist for DSM-5 (PCL-5) in Chinese stroke patients. BMC Psychiatry. 2023;23(1):16. doi: 10.1186/s12888-022-04493-y

79. Nakash O, Nagar M, Kanat-Maymon Y. Clinical use of the DSM categorical diagnostic system during the mental health intake session. Int J Methods Psychiatr Res. 2015;24(3): 206–215. doi: 10.1002/mpr.1463.

80. R Core Team. R: A language and environment for statistical computing [Internet]. Vienna (Austria): R Foundation for Statistical Computing; 2022 [cited 2025 Jul 8]. Available from: https://www.R-project.org/

81. Balduzzi S, Rücker G, Schwarzer G. How to perform a meta-analysis with R: a practical tutorial. Evid Based Ment Health. 2019;22(4):153–160. doi:10.1136/ebmental-2019-300117

82. Quintana MI, Andreoli SB, Jorge MR, Gastal FL, Miranda CT. Validity of the Composite International Diagnostic Interview (CIDI) for diagnosing post-traumatic stress disorder in Brazil. Rev Bras Psiquiatr. 2012;34(3): 247–253. doi: 10.1016/j.rbp.2012.01.003.

83. Prins A, Bovin MJ, Smolenski DJ, Marx BP, Kimerling R, Jenkins-Guarnieri MA, et al. The Primary Care PTSD Screen for DSM-5 (PC-PTSD-5): development and evaluation within a veteran primary care sample. J Gen Intern Med. 2016;31(10): 1206–1211. doi: 10.1007/s11606-016-3703-5.

84. Williamson V, Stickley A, Romppanen J, Van Bortel T. Diagnostic accuracy of the Primary Care PTSD Screen for DSM-5 (PC-PTSD-5): a systematic review and meta-analysis. J Affect Disord. 2022;310: 246–257. doi: 10.1016/j.jad.2022.04.048.

85. Zhu W, Li Y, Ma X, Yang H, Wang Z, Shi R, et al. Bibliometric analysis of post-traumatic stress disorder in forensic medicine: research trends, hot spots, and prospects. Front Psychol. 2023;13: 1074999. doi: 10.3389/fpsyg.2022.1074999.

86. Golubeva E, Zeltser A, Zorkina Y, Ochneva A, Tsurina A, Andreyuk D, et al. Epigenetic alterations in post-traumatic stress disorder: comprehensive review of molecular markers. Complex Psychiatry. 2024;10(1-4): 71–107. doi: 10.1159/000541822.

87. Uddin M, Ratanatharathorn A, Koenen KC, Aiello AE, Bromet EJ, Galea S, et al. Epigenetic meta-analysis across three civilian cohorts identifies NRG1 and HGS as blood-based biomarkers for post-traumatic stress disorder. Epigenomics. 2018;10(12): 1585–1601. doi: 10.2217/epi-2018-0070.

88. Sustretov A, Kuznetsov V. Epigenetic contributors to PTSD: a comprehensive review of DNA methylation and gene expression studies. Neurosci Biobehav Rev. 2024;157: 105511. doi: 10.1016/j.neubiorev.2024.105511.

89. Kang HJ, Yoon S, Kim H, Park JY, Lee HK, Kim SH, et al. FKBP5-associated miRNA signature as a putative biomarker for PTSD. J Affect Disord. 2020;263: 266–274. doi: 10.1016/j.jad.2019.11.146.

90. Holder N, Ranney RM, Bernhard PA, Holliday R, Vogt D, Hoffmire CA, et al. Which veterans with PTSD are most likely to report being told of their diagnosis? J Psychiatr Res. 2023;170: 158–166. doi: 10.1016/j.jpsychires.2023.03.026.

91. Christey-Reid P, Thompson R. Auditing the treatment of post-traumatic stress disorder in patients experiencing inner-city homelessness. BJPsych Open. 2023;9(S184): e72. doi: 10.1192/bjo.2023.431.

92. Kohrt BA, Mendenhall E, Brown PJ, Caldéron M, Krause KR, Lovell A, et al. Cultural concepts and the DSM-5: looking at posttraumatic stress disorder. Front Psychiatry. 2019;10: 439. doi: 10.3389/fpsyt.2019.00439.

93. McFarlane J, Maddoux J, Paulson RM, Symes L, Jouriles E. An evidence-based assessment tool for estimating future post-traumatic stress disorder: a 7-year follow-up study. J Womens Health (Larchmt). 2020;29(4): 508–514. doi: 10.1089/jwh.2019.7707.

94. Harnett NG, Fani N, Michopoulos V, Yehuda R, Ressler KJ. Structural inequities contribute to racial/ethnic differences in amygdala resting-state functional connectivity associated with PTSD. Neuropsychopharmacology. 2023;48(1): 127–135. doi: 10.1038/s41386-022-01427-1.

95. Karchoud J, Haagsma J, Karaban I, Hoeboer C, van de Schoot R, Olff M, et al. Long-term PTSD prevalence and associated adverse psychological, functional, and economic outcomes: a 12–15 year follow-up of adults with suspected serious injury. Eur J Psychotraumatol. 2024;15: 2401285. doi: 10.1080/20008066.2024.2401285.

96. Al-Saffar S, Borgå P, Hällström T. Long-term consequences of unrecognised PTSD in general outpatient psychiatry. Soc Psychiatry Psychiatr Epidemiol. 2002;37(12): 580–585. doi: 10.1007/s00127-002-0586-z.

97. Ford JD, Hawke J, Alessi SM, Ledgerwood D, Petry NM. Psychological trauma and PTSD symptoms as predictors of substance dependence treatment outcomes. Behav Res Ther. 2007;45(10): 2417–2431. doi: 10.1016/j.brat.2007.04.001.

98. Steinert C, Hofmann M, Leichsenring F, Kruse J. The course of PTSD in naturalistic long-term studies: high variability of outcomes. A systematic review. Nord J Psychiatry. 2015;69(7): 483–496. doi: 10.3109/08039488.2015.1005023.

99. Handley TE, Kelly BJ, Lewin TJ, Coleman C, Stain HJ, Weaver N, et al. Long-term effects of lifetime trauma exposure in a rural community sample. BMC Public Health. 2015;15: 2490. doi: 10.1186/s12889-015-2490-y.

100. Wu Y, Mao Y, Wang Y, Yan J, Cao H, Yuan T-F. Systematic review of machine learning in PTSD studies for outcome prediction: a roadmap for future research. J Affect Disord. 2023;325: 274–284. doi: 10.1016/j.jad.2022.12.243.

101. Eyo-Udo U, Apeh ET. Review of ethical considerations and dilemmas in the field of AI in mental health. AI Ethics. 2025;2(1): 23–34. doi: 10.1007/s43681-025-00297-9.

102. Jha A, Durak G, Harrell M. A conceptual framework for applying ethical principles of AI in healthcare. Front Digit Health. 2025;3: 121. doi: 10.3389/fdgth.2025.00121.

103. McKenzie JE, Brennan SE, Ryan RE, Thomson HJ, Johnston RV, Thomas J. Chapter 3: Defining the criteria for including studies and how they will be grouped for the synthesis. In: Higgins JPT, Thomas J, Chandler J, et al., editors. Cochrane Handbook for Systematic Reviews of Interventions. 2nd ed. Chichester, UK: John Wiley & Sons; 2019.

104. Tricco AC, Lillie E, Zarin W, O’Brien KK, Colquhoun H, Levac D, et al. PRISMA extension for scoping reviews (PRISMA-ScR): checklist and explanation. Ann Intern Med. 2018;169(7): 467–473. doi: 10.7326/M18-0850.

105. Marinova Z, Maercker A. Diagnostic and methodological issues in epidemiological studies of PTSD. In: Beck JG, Sloan DM, editors. The Oxford Handbook of Traumatic Stress Disorders. Oxford University Press; 2012.

106. Song F, Parekh S, Hooper L, Loke YK, Ryder J, Sutton AJ, et al. Dissemination and publication of research findings: an updated review of related biases. Health Technol Assess. 2010;14(8): iii, ix–xi, 1-193. doi: 10.3310/hta14080.

107. Fortney JC, Pyne JM, Kimbrell TA, Hudson TJ, Robinson DE, Schneider R, et al. Telemedicine-based collaborative care for posttraumatic stress disorder: a randomized clinical trial. JAMA Psychiatry. 2015;72(1): 58–67. doi: 10.1001/jamapsychiatry.2014.1575.

108. Kohrt BA, Mendenhall E, Brown PJ, Caldéron M, Krause KR, Lovell A, et al. Cultural concepts and the DSM-5: looking at posttraumatic stress disorder. Front Psychiatry. 2019;10: 439. doi: 10.3389/fpsyt.2019.00439.

109. Wu Y, Mao Y, Wang Y, Yan J, Cao H, Yuan T-F. Systematic review of machine learning in PTSD studies for outcome prediction: a roadmap for future research. J Affect Disord. 2023;325: 274–284. doi: 10.1016/j.jad.2022.12.243.

110. Gow A. H. Post-traumatic stress disorder diagnostic accuracy rates in clinical settings: a systematic review and meta-analysis [Internet]. OSF; 2025. Available from: osf.io/umwxd

111. Gow A. H. Post-traumatic stress disorder diagnostic accuracy rates in clinical settings: a systematic review and meta-analysis [Internet]. OSF; 2025. Available from: osf.io/xzyhp

