## Supplemental Table 1 for "Post-traumatic stress disorder diagnostic accuracy rates in clinical settings: a systematic review and meta-analysis"

### TABLE S1: CHARACTERISTICS OF THE INCLUDED STUDIES

***Note:*** *This supplementary document shares references with the main manuscript, which ends at reference [111].*

Table S1.1 Studies on PTSD diagnostic accuracy in specialized MH (included in the meta-analysis).

| Study ID | Type of facility | Initial PTSD diagnosis | Number of participants | True PTSD diagnosis | PTSD diagnostic accuracy | Reasons for accuracy level |
|---|---|---|---|---|---|---|
| **Bonn-Miller et al. (2012) [47]** | Mental health clinics, including specialty Substance Use Disorder (SUD) clinics at a VA Medical Center, USA. | PTSD diagnosis documented in VA electronic medical records (EMRs). | 84 military veterans with cannabis use disorder. | PTSD diagnosed using Clinician-Administered PTSD Scale (CAPS); 31 (36.9%) met DSM-IV criteria for PTSD. | Diagnostic accuracy 58.1% (18/31 CAPS-confirmed PTSD cases documented in EMRs). | Moderate under-recognition of PTSD among veterans with co-occurring cannabis use disorder. Underdiagnosis possibly due to lack of structured assessments in routine care and clinical focus on substance use symptoms. Structured interviews were audio recorded and reviewed to ensure reliability. Sample |

| | | | | | | |
|---|---|---|---|---|---|---|
| | | | | | | mostly older male veterans, limiting generalizability. Emphasizes need for regular structured assessments in VA MH services. |
| **Cusack et al. (2006) [51]** | Community mental health centre (psychosocial rehabilitation program), U.S. public mental health system. | PTSD documented in clinical charts based on routine care diagnosis. | 142 adults with serious mental illness (SMI) and psychiatric hospitalization history. | PTSD symptoms assessed with Trauma Assessment for Adults Self-Report Version (TAA) and PTSD Checklist (PCL); 19-30% met PTSD criteria depending on scoring method. | Diagnostic accuracy 11.6%: only 5 of 43 PTSD cases identified by DSM-based PCL method had a documented diagnosis in charts. | High trauma exposure (87%) and high PTSD prevalence (19–30%) based on self-report screening, but extremely low clinical recognition. Underdiagnosis attributed to limited clinician recognition and documentation despite active trauma initiatives at the centre. Use of validated self-report tools without structured clinical interviews limits diagnostic |

| | | | | | | |
|---|---|---|---|---|---|---|
| | | | | | | confirmation. High functional impairment and complex comorbidity in this SMI population may further complicate detection. Moderate risk of bias due to self-report reliance and lack of gold-standard interviews. |
| **da Silva et al. (2019) [38]** | University outpatient psychiatric clinic, Institute of Psychiatry, Universidade Federal do Rio de Janeiro, Brazil. | Medical records review for PTSD diagnosis made by psychiatrists in training | 200 psychiatric outpatients; mean age 48 years, 59% women. | PTSD module of the SCID-IV used to assess current PTSD; 41 (20.5%) met criteria. | Diagnostic accuracy 2.4%: only 1 of 41 patients meeting PTSD criteria had prior official diagnosis; underdiagnosis rate 97.6%. | Diagnostic complexity and symptom overlap with other disorders complicate recognition; patients’ avoidance and underreporting of trauma further hinder diagnosis; diagnosis supervised by clinical experts though no formal inter-rater reliability reported; |

| | | | | | | |
|---|---|---|---|---|---|---|
| | | | | | | moderate risk of bias mainly due to selection and detection limitations. |
| **de Bont et al. (2015) [52]** | Secondary or tertiary mental healthcare across 13 long-term MH organizations in the Netherlands | *"[...] Clinical charts of all 2608 participants, either as the primary, secondary, or tertiary Axis 1 diagnosis."* [41, p. 412] | 2608 screened; subsample of 455 assessed with CAPS + MINI-Plus | CAPS used for PTSD diagnosis in subsample of 455; 146 (32.1%) met PTSD criteria by CAPS. | Diagnostic recognition rate of approximately 8.2%: only 12 of 146 CAPS-confirmed PTSD cases were documented in charts (true positives among CAPS cases). Chart PTSD diagnosis prevalence was 0.5% (13 of 2608) overall. | Under-recognition of PTSD is substantial; reasons include hierarchical diagnostic systems (psychotic disorder diagnosis prioritized), clinician hesitance to discuss trauma with psychotic patients, and challenges in routine PTSD assessment. TSQ screening showed good sensitivity (78.8%) and specificity (75.6%) at optimal cut-off (6). Methodological notes include moderate selection bias due to multiple imputation |

| | | | | | | |
|---|---|---|---|---|---|---|
| | | | | | | and logistic regression for prevalence estimation; low reporting and performance bias; unclear detection and attrition bias due to limited blinding/dropout data. |
| **Holowka et al. (2014) [39]** | Veterans Affairs (VA) mental health system, Iraq and Afghanistan veterans from Project VALOR registry, U.S. | PTSD diagnoses extracted from VA Problem List and Encounter data in the National Patient Care Database (EMR-based). | 1,649 veterans. | PTSD module of SCID administered by blinded, trained doctoral-level clinicians; 1,039 (63%) met criteria for current PTSD by SCID. | Diagnostic accuracy of 73.2% based on Problem List EMR data compared to SCID (886 true positives, 289 false positives, 153 false negatives, 321 true negatives). Recognition rate was 85.3% (true positives | Overreliance on informal clinical assessments led to 26.8% misclassification (under- or over-diagnosis). False negatives linked to lower combat exposure, fewer avoidance symptoms, less impairment, and lower panic disorder rates. False positives associated with emotional treatment seeking and lower |

| | | | | | | |
|---|---|---|---|---|---|---|
| | | | | | / SCID PTSD cases). Concordance rate 73.2% for Problem List, 72.3% for encounter data. | symptom severity. Limitations include retrospective data, *interviewer* variance, and generalizability restricted to VA veteran populations. Moderate risk of bias due to potential EMR coding misclassification and reliance on single structured interview. Emphasizes importance of structured assessments to improve PTSD diagnosis accuracy in clinical practice. |
| **Lommen and Restifo (2009) [54]** | Outpatient psychiatric clinic in the Netherlands, | PTSD diagnosis as documented in medical charts. | 33 adult outpatients. | PTSD assessed by self-report instruments: Trauma History | Diagnostic accuracy (recognition rate) 0%: None of the participants meeting | Underdiagnosis attributed to clinician focus on psychotic symptoms, patient reluctance to disclose trauma, and symptom |

| | | | | | |
|---|---|---|---|---|---|
| | patients with schizophrenia (n=23) or schizoaffective disorder (n=10). | | Questionnaire-Revised (THQ-R), PTSD Symptom Scale–Self-Report (PSS-SR), and Post-Traumatic Cognitions Inventory (PTCI). Using the most conservative DSM-IV scoring (including criterion A1 and symptom threshold ≥ 2), 9.1% met PTSD criteria. Liberal criteria gave 39.4% prevalence. | PTSD criteria had a PTSD diagnosis documented in their charts. | overlap between schizophrenia and PTSD. High correlation between negative post-traumatic cognitions (PTCI) and PTSD symptom severity (r=0.74). Emphasizes need for trauma screening and cognitive models of PTSD in psychotic disorders. Moderate risk of bias due to small sample size, reliance on clinician referrals, and self-report measures rather than structured interviews. |

| | | | | | | |
|---|---|---|---|---|---|---|
| **Marx et al. (2017) [55]** | Veterans Affairs (VA) disability examination, Project VALOR, U.S. veterans. | PTSD diagnoses from Compensation & Pension (C&P) examiners and VA PTSD service-connection status. | 764 veterans. | PTSD assessed by Structured Clinical Interview for DSM-IV (SCID) for current and lifetime PTSD. | Diagnostic concordance: 70.4% current PTSD, 77.7% lifetime PTSD; overall diagnostic accuracy 70.4%. True positives 62.9%, false positives 16.4%, false negatives 13.1%, true negatives 7.4%. Veterans diagnosed by C&P examiners were over 3 times more likely to meet SCID criteria (OR = 3.39, 95% CI [2.25–5.15]). | Racial disparities noted: Black veterans more likely false negatives, White veterans more likely false positives, especially without psychometric testing. Psychometric testing, used in 24.2% of exams, reduced disparities. Study strengths include blinded interviewers, standardized instruments, and consideration of confounders. Low risk of bias overall. |

| | | | | | | |
|---|---|---|---|---|---|---|
| **Schwartz et al. (2005) [56]** | Urban community mental health clinic, U.S., primarily African American outpatients. | Clinical PTSD diagnoses extracted from medical charts. | 184 outpatients recruited and surveyed; of these, 72 completed all assessments including the SCID-I; 66 medical charts were available for review. | PTSD diagnosis confirmed via SCID-I (Structured Clinical Interview for DSM-IV Axis I Disorders) on subsample of 72; 29 (40%) met PTSD criteria by SCID-I; 83% of total sample reported trauma meeting DSM-IV Criterion A. | Diagnostic accuracy estimated at 11.5%: among 26 SCID-positive participants with available medical charts, only 3 had documented PTSD diagnoses; large under-recognition evident. | High prevalence of trauma and PTSD in this underserved population. PTSD-positive participants had significantly higher rates of comorbidities (e.g., major depression, nonschizophrenic psychotic disorders). Limitations include reliance on subsample for diagnostic confirmation, incomplete chart availability (66/72), single-site recruitment, retrospective chart review, and potential selection bias due to attrition. Use of validated self-report tools and blinded interviewers enhanced internal validity. |

| | | | | | | |
|---|---|---|---|---|---|---|
| **Wang and Vivek (2013) [57]** | Two inner-city mental health clinics in Queens, New York, U.S. | Clinical chart review for PTSD diagnoses documented during routine care. | 62 psychiatric outpatients voluntarily recruited. | PTSD screening via PTSD Checklist–Civilian Version (PCL-C), cut-off score = 44; 38 participants (61%) screened positive for PTSD. Trauma exposure assessed by Traumatic Life Events Questionnaire (88% reported trauma). | Diagnostic accuracy ~21%: 8 participants (13%) had PTSD diagnosis documented in charts out of 38 positive by PCL-C (8/38). | Significant under-recognition of both PTSD and trauma exposure in clinical records (22% recorded trauma vs. 88% reported). Comorbidities such as major depressive disorder and SSRI prescriptions more common in PTSD-positive group but differences not statistically significant due to small sample size. Limitations include absence of gold-standard structured interviews (e.g., CAPS), potential selection bias from voluntary participation, and routine clinical diagnostic inaccuracies. Highlights urgent need for standardized screening |

| | | | | | | |
|---|---|---|---|---|---|---|
| | | | | | | and diagnostic assessment in outpatient settings. |
| **van Zyl et al. (2008) [43]** | Neuroclinic C, a therapeutic inpatient unit specializing in mood and anxiety disorders at Stikland Hospital, Bellville, Cape Town, South Africa. | PTSD diagnosis at admission and discharge recorded in routine clinical care. | 40 consenting inpatients (82.5% female; mean age 35.65 years). | PTSD diagnosis based on Clinician-Administered PTSD Scale (CAPS) for DSM-IV, administered during study; 16 (40%) met criteria for PTSD. | Diagnostic accuracy 0%: none of the 16 CAPS-confirmed PTSD cases were diagnosed with PTSD on admission; among all 293 patients admitted during study period, 30 were diagnosed with PTSD at discharge (including the 16 study cases), indicating under-recognition. | Under-recognition attributed to clinicians not routinely inquiring about trauma exposure and patient reluctance to disclose trauma without prompting. Cannabis use was significantly associated with PTSD ($p = 0.01$). PTSD prevalence higher among women and mixed ethnicity but not statistically significant. Moderate risk of bias due to small sample size, lack of blinding of participants and evaluators, and unclear allocation procedures. CAPS |

| | | | | | | |
|---|---|---|---|---|---|---|
| | | | | | | ensured diagnostic rigor. |
| **Zammit et al. (2018) [25]** | Secondary-care mental health services worldwide; included 29 peer-reviewed studies across multiple countries (USA, UK, Netherlands, Australia, Germany, South Africa, Spain, Turkey, others). | Clinical diagnoses recorded in medical/clinical records reviewed across studies. | Total sample of 6,412 individuals with various psychiatric diagnoses based on DSM or ICD criteria. | PTSD identified using validated self-report questionnaires or structured interviews (varied by study); median PTSD prevalence on screening: 33.3% (IQR 23.4–40.0%). | Median diagnostic recognition rate (accuracy) 11.5% (IQR 2.8–19.4%) across studies; median prevalence of PTSD documented clinically was 2.3% (IQR 1.1–4.5%); median undetected PTSD proportion 28.6% (IQR 18.2–38.6%). | High prevalence of PTSD on screening contrasted with very low clinical recognition globally, especially in USA, inpatient samples, and psychotic disorder populations. Substantial heterogeneity across studies ($I^2 > 90\%$). Validated screening tools showed good specificity; self-report tools may slightly overestimate PTSD but were well validated. Most studies did not mask assessors, possibly introducing bias. Findings consistent across subgroup analyses. Call for routine PTSD |

| | | | | | | |
|---|---|---|---|---|---|---|
| | | | | | | screening trials in secondary care. Moderate to high risk of bias in many studies due to selection and detection factors, but robust overall findings. |
| **Reynolds et al. (2005) [42]** | Specialist inpatient addiction services unit, South West Thames, UK. | PTSD diagnoses documented in medical case notes from routine clinical care. | 52 substance use disorder (SUD) inpatients interviewed and had case notes reviewed. | PTSD diagnosis established via validated structured instruments: Trauma History Questionnaire (THQ), PTSD Symptom Scale–Interview (PSS-I), and Addiction Severity Index (ASI). 20 participants | Diagnostic accuracy 5%: only 1 participant had a documented PTSD diagnosis in their medical notes and was referred for treatment, despite 38.5% meeting criteria. | High under-recognition attributed to PTSD symptoms being embedded in broader psychiatric presentations and routinely overlooked in documentation. PTSD-positive patients showed significantly more trauma-related distress, impaired social and occupational functioning, and higher medical/psychiatric issues on ASI. Significant associations |

| | | | | | | |
|---|---|---|---|---|---|---|
| | | | | (38.5%) met DSM-IV criteria for current PTSD; 27 (51.9%) met criteria for lifetime PTSD. | | between trauma and substance use coping were noted. Moderate risk of bias due to small sample size and reliance on retrospective chart review, balanced by rigorous use of validated diagnostic tools and in-depth assessments. |
| **Lewis et al. (2017) [41]** | UK, National Centre for Mental Health cohort; adults with diagnosed mental disorders recruited from primary and secondary care | Self-reported clinician diagnosis of PTSD. | 1,946 adults with current or past mental disorders who completed the Trauma Screening Questionnaire (TSQ). | PTSD assessed using TSQ anchored to DSM-5 qualifying traumatic events; 438 (23%) screened positive for probable current PTSD. | Diagnostic accuracy ~38.6%: 169 participants both screened positive on TSQ and self-reported clinician diagnosis; 271 (13.9%) had undetected PTSD | High prevalence of trauma exposure (40.9%) and undetected PTSD (13.9%) across broad mental health population. Undetected PTSD more common among females, younger age at first psychiatric contact, and lower socioeconomic status (income, |

| | | | |
|---|---|---|---|
| | and community settings. | (screened positive but no clinician diagnosis). | education). Trauma types associated with undetected PTSD included childhood abuse, sexual assault, and domestic violence. Undetected PTSD prevalence was highest in personality disorders, anxiety, bipolar, and psychotic disorders. Limitations include reliance on self-reported clinician diagnoses, lack of structured clinical interviews, potential underreporting of trauma, and screening limited to current PTSD. Emphasizes need for routine PTSD screening in psychiatric services to improve recognition and treatment. |

| | | | | | | |
|---|---|---|---|---|---|---|
| **Meltzer et al. (2012) [58]** | Urban safety-net primary care hospital serving patients receiving mental health treatment for various disorders (Boston Medical Center, U.S.). | PTSD diagnoses documented in EMRs from patients utilizing mental health services. | 592 adult primary care patients enrolled; 133 met PTSD criteria by diagnostic interview (Composite International Diagnostic Interview [CIDI]). | PTSD diagnosis via Composite International Diagnostic Interview (CIDI); comorbid depression assessed with PHQ-9. | Diagnostic accuracy ~11.5%: only 14 of 133 patients with interview-confirmed PTSD had PTSD documented in their EMRs. | Study measured under-recognition of PTSD among patients receiving treatment for other mental health disorders in a primary care setting. Despite high comorbidity with depression, PTSD was rarely documented, possibly due to diagnostic overshadowing, therapeutic overlap, and lack of trauma-focused inquiry. Findings highlight the need for improved PTSD screening and diagnosis within mental health treatment contexts, even when delivered in primary care. Moderate risk of bias due to cross-sectional design and |

| | | | | | | |
|---|---|---|---|---|---|---|
| | | | | | | reliance on EMR data. |
| **Kostaras et al. (2016) [40]** | Psychiatric outpatient units, University Hospital of Athens, Greece. | PTSD diagnosis documented in medical records by treating clinicians. | 101 outpatients with Major Depressive Disorder (MDD). | PTSD diagnosed using the Mini-International Neuropsychiatric Interview (MINI); trauma assessed with Life Events Checklist. | Diagnostic accuracy 28.2%: 11 of 39 patients with PTSD diagnosis via MINI had PTSD documented in their medical records. | High prevalence of MDD-PTSD comorbidity (38.6% lifetime, 26.7% current); all PTSD patients also had MDD. PTSD often preceded MDD onset (mean 6.2 years earlier). Under-recognition likely due to clinicians' reluctance to assess trauma/PTSD symptoms, lack of routine trauma screening, and patient avoidance in disclosing trauma. Chronic depression, prolonged/repeated trauma, male gender, younger symptom onset, lower education, and reduced functioning were predictors of |

| | | | | | | |
|---|---|---|---|---|---|---|
| | | | | | | MDD-PTSD. PTSD diagnosis documented only in 28.2%, implying substantial underdiagnosis and delayed/inadequate treatment. Moderate risk of bias due to retrospective design and sample size. |
| **Gravely et al. (2011) [23]** | Veterans Affairs (VA) health system; stratified by clinic type: Primary care clinics and specialized mental health | PTSD diagnosis codes recorded in VA administrative data (ICD-9 code 309.81), based on routine clinical assessments. | 4,777 veterans with at least one new PTSD diagnosis. | PTSD screening using self-reported PTSD Checklist (PCL) with cutoff ≥50 as gold standard. | PPV for ≥1 diagnosis: Primary care: 69.3% (2,071 veterans diagnosed), Non-PTSD MH clinics: 79.9% (2,122 veterans), PTSD specialty clinics: 77.6% (540 | PTSD diagnoses in specialized MH clinics are more likely true cases than those in primary care. Nearly 31% of primary care diagnoses may be false positives. Lower PPV among older veterans. Using ≥2 diagnoses improves accuracy but reduces sample size by |

| | | | | | | |
|---|---|---|---|---|---|---|
| | clinics (including PTSD specialty clinics) in the U.S. | | | | veterans). For veterans ≥65 years old, PPV was 61.4% for ≥1 diagnosis, improving to 76.1% for ≥2 diagnoses. Overall PPV was 74.8%; for ≥2 diagnoses, 81.8%. | ~40%. Study only includes diagnosed cases; false negatives and sensitivity not assessed. Findings are generalizable to real-world VA clinical settings. Included in both PC and MH subgroups due to stratified data. |
| **Magruder et al. (2008) [59]** | VA primary care clinics at four southeastern U.S. VA medical centers. | PTSD diagnosis documented by providers via ICD-9 code (309.81) in VA electronic medical records. | 819 veterans who completed structured diagnostic assessments. | PTSD diagnosed using Clinician-Administered PTSD Scale (CAPS) via telephone within 2 months of primary care visit; 98 (12%) met DSM-IV PTSD | Recognition rate (diagnostic accuracy) 43%: 42 of 98 CAPS-confirmed PTSD cases had documented PTSD diagnosis in EMRs. | Substantial under-recognition of PTSD in primary care prior to routine screening. Higher recognition associated with age 50–64, war-zone service (OR=3.0), musculoskeletal pain diagnoses (OR=3.8), and prior substance use disorder |

| | | | |
|---|---|---|---|
| | | criteria. | (OR=9.91). PTSD symptom clusters B (reexperiencing) and D (hyperarousal) linked to higher recognition; avoidance/numbing symptoms less likely recognized. Emotional impairment increased detection likelihood. No significant effect of race, sex, education, employment. Moderate bias risk due to retrospective EMR review and missing provider/system data. Highlights need for provider education and structured screening to improve PTSD detection. |

| | | | | | | |
|---|---|---|---|---|---|---|
| **Ivanov et al. (2012) [60]** | Psychiatric emergency unit, University Hospital Lausanne, Switzerland. | PTSD diagnosis recorded in routine clinical charts (historical sample). | 403 enrolled (316 analyzed for MINI assessment). | PTSD diagnosed using the PTSD module of the MINI; 64 (20.3%) met criteria for current PTSD based on MINI. | Diagnostic accuracy 3.47%: only 21 PTSD diagnoses documented in charts vs. an estimated 605 expected cases based on MINI prevalence. | Severe under-recognition of PTSD in psychiatric emergency settings. Only 37.5% of MINI-confirmed PTSD cases had PTSD documented in charts in the prospective sample. Higher prevalence among refugees, migrants, and those with multiple psychiatric comorbidities. Barriers include time constraints, limited follow-up, and impact on therapeutic relationships. Moderate risk of bias from lack of blinding, exclusion of some patients, and reliance on unstructured clinical diagnoses. |

| | | | | | | |
|---|---|---|---|---|---|---|
| **Tiet et al. (2013) [61]** | VA outpatient specialty clinics: Substance Use Disorder (SUD) and General Mental Health (MH) clinics. | PTSD diagnosis documented in VA electronic medical records (9 months before/after structured interview). | 400 veterans (158 SUD, 242 MH). | PTSD diagnosed using the Computerized Diagnostic Interview Schedule for DSM-IV (C-DIS-IV): 58 (36.7%) SUD, 128 (52.9%) MH. | Diagnostic accuracy: 48.3% in SUD clinics (28/58 cases recognized), 60.2% in MH clinics (77/128 cases recognized), combined accuracy 56.5%. | Under-recognition likely due to symptom overlap with other psychiatric disorders, clinical workflow issues, and possible under-documentation. Use of validated structured interview as reference standard enhances accuracy estimate. Screening tools (PCL-Bliese-4, PC-PTSD, etc.) showed good psychometric properties, suggesting routine use may improve detection. Moderate risk of bias due to non-clinician interviewers and timing differences between screening and diagnostic assessments. |

Table S1.2 Studies on PTSD diagnostic accuracy in primary healthcare (included in the meta-analysis).

| Study ID | Type of facility | Initial PTSD diagnosis | Number of participants | True PTSD diagnosis | PTSD diagnostic accuracy | Reasons for accuracy level |
|---|---|---|---|---|---|---|
| **Bohnert et al. (2016) [24]** | Veterans Health Administration (VHA) primary care, PC-MHI, specialty MH clinics. | PTSD diagnosis documented in EMRs via ICD-9 code 309.81 within 1 year of positive PC-PTSD screen. | 21,427 veterans with positive PC-PTSD screen (18,157 primary care only; 1,507 PC-MHI; 1,763 specialty MH). | PTSD diagnosis inferred from medical records within 1 year of screening; no structured diagnostic interview used. | Diagnostic accuracy: Primary care only 47.9%, PC-MHI 60.3%, Specialty MH 68.0%; Combined MH 64.4%. | Integrated mental health services (PC-MHI) and specialty MH care significantly increased PTSD diagnosis compared to primary care only. Use of administrative data introduces moderate bias risk (no structured interview, potential misclassification). Diagnosis rates include any within 1 year post-screen, not just same-day diagnosis. Treatment initiation also higher |

| | | | | | | |
|---|---|---|---|---|---|---|
| | | | | | | with PC-MHI and specialty care. Findings support integrated care models to improve PTSD recognition and treatment in veteran populations. |
| **Meltzer-Brody et al. (2004) [62]** | University-affiliated outpatient gynecology primary care clinic, U.S. | PTSD diagnosis based on clinical records compared to MINI structured clinical interview. | 292 women screened; 32 completed diagnostic interview. | PTSD diagnosed using MINI structured clinical interview; 25 of 32 (78%) met criteria for PTSD. | <u>Diagnostic accuracy 12%: 3 of 25 PTSD-positive women documented as receiving psychiatric care</u> | Undetected PTSD (general). |
| **Prins et al. (2003) [29]** | VA general and women's primary care clinics (Palo | PTSD diagnosis documented in VA medical records (chart | 188 veterans (134 with available medical | PTSD diagnosed using CAPS structured clinical interview; 46 | Real-world diagnostic accuracy 39% (52/134 CAPS-confirmed PTSD | Under-recognition of PTSD in primary care VA settings; many cases missed during routine care. PC-PTSD |

| | | | | | | |
|---|---|---|---|---|---|---|
| | Alto and Menlo Park, CA, USA). | review). | records). | (24.5%) met PTSD criteria by CAPS. | cases documented in charts). | screening tool showed strong correlation with CAPS and outperformed the PTSD Checklist (PCL) on most diagnostic metrics. PC-PTSD preferred in primary care for brevity and ease of use. Sensitivity and specificity varied by sex. Moderate risk of bias due to sample attrition, limited record availability, volunteer bias, and single-site design. The study focuses on non-specialized, fast-paced primary care clinics within the VA system where brief screening tools are necessary. |

| | | | | | | |
|---|---|---|---|---|---|---|
| **Taubman-Ben-Ari et al. (2001) [30]** | National sample of 26 primary care clinics in Israel operated by the largest national health provider. | Primary care physician clinical assessment based on blinded clinical impression forms. | 2,975 primary care patients. | PTSD prevalence based on PTSD Inventory (DSM-III criteria) self-report: 247/2,975 (8.3%) PTSD cases. | Recognition rate (sensitivity) 2.4%: Only 6 of 247 PTSD-positive cases identified by physicians; PPV 42.9%. | Extremely low PTSD recognition rate in primary care. Physicians identified only a small fraction of PTSD cases despite a relatively high prevalence. Diagnostic accuracy metric inflated by the large number of non-PTSD cases correctly not diagnosed (91.6%), which is less meaningful clinically. Use of self-report PTSD inventory rather than structured clinical interview, and DSM-III criteria limit diagnostic precision. Large, representative sample and physician blinding strengthen study validity. |

| | | | | | | |
|---|---|---|---|---|---|---|
| | | | | | | Moderate to high risk of bias due to self-report measures and older diagnostic criteria. |
| **Graves et al. (2011) [31]** | Academically affiliated urban primary care clinics, U.S. | PTSD diagnosis documented by providers via clinical records or evidence of mental health treatment (medication or psychotherapy). | 375 participants who completed structured interviews (SCID and CAPS). | PTSD diagnosed using SCID and CAPS structured diagnostic interviews; 91 (24.3%) met current PTSD criteria. | Diagnostic accuracy 30.8%: 28 of 91 PTSD-positive individuals recognized/treated in real-world care. | High under-recognition and treatment gap in primary care among African American adults. Many PTSD-positive patients never received mental health treatment or disclosed symptoms to providers. Contributing factors include stigma, clinician training gaps, patient presentation with physical rather than psychological symptoms, and systemic barriers. High comorbidity rates complicated |

| | | | | | | |
|---|---|---|---|---|---|---|
| | | | | | | recognition. Sample limited by size and convenience sampling. |
| **Liebschutz et al. (2007) [32]** | Hospital-based primary care internal medicine clinic, Boston Medical Center, U.S. | PTSD documentation in EMRs (diagnosis, medical notes, ICD-9 codes) | 636 recruited, 609 completed screening, 597 included in diagnostic analysis (complete CIDI data). | PTSD diagnosed using Composite International Diagnostic Interview (CIDI); 117 (19.6%) met criteria for PTSD (based on 597 analyzed). | Diagnostic accuracy 11.1%: only 13 of 117 PTSD-positive patients had PTSD documented in their medical records. | Although 636 patients were recruited and 609 completed screening, only 597 had complete CIDI diagnostic data, which was necessary for reliable prevalence and diagnostic accuracy calculations. This analytic subset aligns with the methodological rigor of the study. Under-recognition was significant, with high comorbidity potentially leading to diagnostic overshadowing. The study was limited by |

| | | | | | | |
|---|---|---|---|---|---|---|
| | | | | | | single-site recruitment and English-language requirement, which may affect generalizability. |
| **Lu et al. (2023) [63]** | Urban primary care HIV clinic, Infectious Diseases Unit. | PTSD documented by clinicians in EMRs based on retrospective chart review. | 135 primarily African American HIV patients. | Probable PTSD via PC-PTSD-5 screening (cut-off ≥ 3); 67 screened positive (49.6%). | Diagnostic accuracy 23.9% (16 of 67 probable PTSD cases documented in EMRs). | Significant underdiagnosis consistent with previous literature on low PTSD detection rates in HIV care. Diagnosis based on EMR documentation only; screening tool (PC-PTSD-5) used as reference standard rather than structured clinical interview, limiting diagnostic precision. Small, single-site sample primarily African American limits generalizability. |

| | | | | | | |
|---|---|---|---|---|---|---|
| | | | | | | Retrospective chart review may introduce documentation bias. Strong correlations found between PTSD symptoms and depression, insomnia, anxiety, and alcohol use, indicating co-occurring conditions that complicate clinical recognition. Supports need for trauma-informed care and routine screening in high-risk urban HIV primary care settings. Moderate risk of bias due to methodological limitations and sample size. |
| **Carey et al. (2003) [33]** | Primary healthcare | PTSD diagnosis based on | 201 adults attending | PTSD diagnosed via standardized | Diagnostic accuracy 0%: None of the 40 | High prevalence of trauma and PTSD, with significant |

| | | | | | | |
|---|---|---|---|---|---|---|
| | clinic in a South African township. | retrospective chart review in medical records. | primary care clinic. | diagnostic interviews and instruments; 40 (19.9%) met current PTSD criteria. | PTSD cases were documented in medical records. | comorbidity including major depression (75%), somatization (35%), and panic disorder (25%). Despite this, clinicians failed to identify any PTSD cases or trauma exposure. Low psychotropic medication prescription (1%) indicates near-total lack of PTSD recognition and treatment. Moderate risk of bias due to single-site design and retrospective chart review. Highlights a critical gap in PTSD recognition in primary care in this setting. |
| **Seal et al.** | VA primary | PTSD screening | 750 post-9/11 | No structured | 24% of screen- | Significant gap between PTSD |

| | | | | | | |
|---|---|---|---|---|---|---|
| **(2008) [64]** | care and community-based outpatient clinics in the U.S. | positive using Primary Care PTSD Screen (PC-PTSD) from post-deployment screening program. | veterans; 338 screened. | clinical interview; data based on positive screeners with follow-up MH clinic visits. | positive veterans completed a VA MH appointment within 90 days; 73% completed mental health appointment eventually. | symptom detection and timely MH referral. Screening increased MH clinic attendance but many positive cases lacked follow-up. No gold-standard diagnostic confirmation; reliance on chart data for follow-up MH visits; lack of data on non-VA MH treatment. Moderate risk of bias due to single-site design and absence of formal PTSD diagnosis. Demonstrates real-world challenges in linkage from screening to treatment in primary care settings. |
| **Kimerling et** | VA primary | PTSD diagnosis | 134 veterans | PTSD diagnosed | Recognition rate | Under-recognition of PTSD |

| | | | | | | |
|---|---|---|---|---|---|---|
| **al. (2006) [34]** | care clinics (general internal medicine and women's health outpatient clinics). | documented in VA electronic medical records (EMRs) via chart review. | recruited; 34 (25.4%) met PTSD criteria on CAPS, used as gold standard. | using Clinician-Administered PTSD Scale (CAPS) by blinded psychologists. | 38.2%: 13 of 34 CAPS-confirmed PTSD cases had documented PTSD diagnosis in EMRs. | despite integrated healthcare system. Brief self-report Breslau 7-item screen showed high sensitivity (85%) and specificity (84%) at cutoff 4. Underdiagnosis attributed to provider discomfort, competing demands, lack of structured screening. Study limited by small sample size and single-site VA setting, but rigorous methodology supports findings. Highlights need for routine use of brief validated PTSD screeners in primary care to improve detection. |

Table S1.3 Studies on clinicians' ability to diagnose PTSD accurately under various scenarios and the impact of diagnostic biases (included in the meta-analysis).

| Study ID | Type of facility | Initial PTSD diagnosis | Number of participants | True PTSD diagnosis | PTSD diagnostic accuracy | Reasons for accuracy level |
|---|---|---|---|---|---|---|
| **McGuire et al. (2022) [66]** | UK child and adolescent mental health services (various sectors including NHS, charities, social care). | Online survey responses from 270 MH professionals asked to assign diagnosis based on vignettes of a teenage boy with PTSD symptoms, one in foster care and one living | 270 MH professionals: randomized 1:1 to two vignette groups — 135 saw foster care vignette, 135 saw biological mother vignette. | Not applicable—diagnosis based on vignette assessment rather than real patients. | PTSD diagnosis selected as primary diagnosis by 22.7% overall; 14.5% in foster care group and 31.0% in biological mother group. | Contextual bias influenced clinician PTSD diagnostic decisions; children in foster care less likely to be diagnosed with PTSD despite identical symptom presentation. Only ~33.9% recommended NICE-approved PTSD treatments, with significant group difference (27.2% foster care vs 40.4% biological mother). Moderate risk of bias due to vignette |

| | | | | | | |
|---|---|---|---|---|---|---|
| | | with biological mother. | | | | design, lack of blinding, and use of hypothetical scenarios. Highlights diagnostic and treatment biases related to social context. |
| **McKenzie and Smith (2006) [65]** | Mental health professionals in Victoria, Australia (GPs, psychologists, psychiatrists). | PTSD Knowledge Questionnaire assessing clinician knowledge across 8 domains. | 154 MH professionals (59 GPs, 56 psychologists, 39 psychiatrists). | Not a diagnostic accuracy study; assessed clinician knowledge on PTSD, beliefs, and knowledge gaps. | Clinician PTSD knowledge assessment score: 74% average correct responses (GPs 68%, psychologists 77%, psychiatrists 79%) | Significant variation between professions; GPs showed lower knowledge and greater uncertainty, potentially impairing PTSD recognition. Poor self-awareness of knowledge gaps observed. Moderate risk of bias from sample selection and response. |

Table S1.4 Evidence of PTSD underdiagnosis in clinical settings (not included in the meta-analysis – systematic review moved to the qualitative synthesis section due to study overlap)

| Study ID | Type of facility | Initial PTSD diagnosis | Number of participants | True PTSD diagnosis | PTSD diagnostic accuracy | Reasons for accuracy level |
|---|---|---|---|---|---|---|
| **Greene et al. (2016) [26]** | Primary care clinics (various international settings). | Detection by primary care physicians using medical records and clinical judgment, compared to structured assessments (CAPS, SCID, CIDI). | 23,941 (across 27 studies). | Prevalence ranged 2% to 39% depending on population and trauma exposure; structured assessments identified substantially more PTSD cases than clinical judgment alone. | Detection rates ranged 0% to 52%, weighted average ~15.46% across studies. | Underdiagnosis due to reliance on clinical judgment without standardized tools; patients often present with somatic complaints rather than psychiatric symptoms; high psychiatric comorbidity complicates diagnosis; lack of routine screening and provider training. |

Table S1.5 Evidence of PTSD underdiagnosis in clinical settings (not included in the meta-analysis)

| Study ID | Type of facility / setting | Type of data collected | Sample specifications | Main findings on PTSD recognition | Key conclusions on PTSD under-recognition |
|---|---|---|---|---|---|
| **Bovin et al. (2021) [67]** | VA primary care clinics (U.S.). | Cross-sectional diagnostic validation study; PC-PTSD-5 vs CAPS-5. | 396 veterans attending VA primary care; mixed sex; no direct clinical coding data. | 17.2% met CAPS-5 criteria for PTSD; PC-PTSD-5 showed excellent screening accuracy (AUC=0.927); no direct data on provider recognition; most PTSD cases not engaged in specialty mental health treatment. | Significant under-recognition implied by low specialty mental health engagement despite high PTSD prevalence; screening tools need sex-specific cut-offs for optimal detection; supports systematic screening in primary care. |
| **Kosowan et al. (2022) [68]** | Pan-Canadian primary care EMR network (CPCSSN). | Retrospective diagnostic coding analysis using ICD-9-CM codes and free-text fields. | 289,523 primary care patients in Manitoba, Canada; adults; large EMR dataset with coding variability. | PTSD diagnosis rate based on coding was very low (1.3%), highlighting substantial underestimation; sensitivity and specificity of case definitions varied, with best method achieving 91.1% sensitivity and | PTSD prevalence is substantially underestimated in primary care EMRs due to reliance on diagnostic codes and documentation variability; improved detection requires advanced methods like natural |

| | | | | | |
|---|---|---|---|---|---|
| | | | | 99.1% specificity. | language processing to extract trauma data from free text notes. |
| **Singer et al. (2024) [69]** | Primary care clinics across 7 Canadian provinces. | Retrospective EMR data analysis. | 689,301 primary care patients from 1,574 providers in 253 clinics in 7 provinces (BC, AB, MB, ON, QC, NS, NL). Diverse adult population from Canadian community primary care settings. | PTSD diagnosis recorded in EMRs was 1.3% (n=8,817). PTSD was strongly associated with comorbid depression, anxiety, alcohol and drug use disorders. PTSD was often documented years after other mental health conditions, suggesting delayed or missed recognition. Patients with PTSD had higher healthcare utilization and resided more often in materially and socially deprived areas. | PTSD is substantially under-recognized in Canadian primary care EMRs. Delays in PTSD documentation relative to other MH diagnoses indicate PTSD often remains a hidden or secondary diagnosis. Enhanced screening and resource allocation are recommended to improve identification and management in primary care, especially for socially and medically complex patients. |
| **Ehlers et al. (2009) [70]** | Primary care (GP clinics in South London). | Cross-sectional survey of GPs' self-reported | 129 GPs surveyed (18% response rate); practices | GPs reported a median estimated PTSD prevalence of 0.2%, much lower than epidemiological data | PTSD is severely under-recognized in primary care with inconsistent adherence to treatment guidelines. |

| | | | | | |
|---|---|---|---|---|---|
| | | estimates and treatment practices. | covering ~4612 patients each; data based on self-report from GPs. | (1.5–3.6%). 27% of GPs had not seen any adult PTSD cases in 3 months; only 15% had read NICE PTSD guidelines; 52% prescribed SSRIs often contrary to NICE recommendations. Trauma-focused CBT/EMDR rarely offered (11%). Significant under-recognition and suboptimal treatment noted. | Low recognition is driven by limited training, time constraints, patient reluctance to disclose trauma, and limited availability/access to recommended trauma-focused therapies. Dissemination of knowledge and improved access (e.g., IAPT services) are needed. |
| **Della Porta (2017) [71]** | Primary care physicians in the U.S. (vignette study). | Experimental vignette-based survey. | 144 primary care physicians randomly assigned to 2 PTSD vignettes (straightforward PTSD and PTSD with physical comorbidity). | 90.3% overall correctly identified PTSD (PTSD-S: 91.0%, PTSD-P: 89.1%). Years of clinical experience and PTSD knowledge scores did not predict recognition accuracy. No significant difference between vignette types. | Primary care physicians can accurately identify PTSD when symptoms are clearly presented; under-recognition in clinical practice likely due to symptom presentation, absence of structured tools, or competing priorities rather than lack of diagnostic skill. |

| | | | | | |
|---|---|---|---|---|---|
| **Kaltman et al. (2011) [72]** | Primary care clinics serving uninsured, low-income immigrant women in the U.S. | Prospective cohort data with structured interviews and medical record review | 138 uninsured, low-income immigrant women from Central and South America attending two primary care clinics; 67.7% screened positive for PTSD in the collaborative care clinic, 64.4% in the on-site therapist clinic. | Approximately 67% of participants screened positive for PTSD via structured interviews, yet PTSD was under-documented in EMRs. More than 50% of participants received no care or care below minimal adequacy thresholds. Collaborative care model was associated with greater PTSD symptom reduction compared to standard co-located care. Many comorbid PTSD and depression cases went untreated. | PTSD is substantially under-recognized and undertreated among low-income immigrant women in primary care, exacerbated by language barriers, cultural stigma, and limited provider training. Collaborative care models improve detection and symptom outcomes. Limitations include small sample size and reliance on self-reports without clinician confirmation, limiting generalizability. |
| **Cook et al. (2017) [73]** | Five large civilian healthcare systems in the | Retrospective administrative claims and EMR data | 5,256 patients diagnosed with PTSD in 2014 across 5 integrated | 16.6% of PTSD diagnoses were made in primary care or non-mental health clinics, while 83.4% were diagnosed by specialty mental health | Significant disparities exist in PTSD detection and treatment quality between primary care and specialty mental health care. Primary care |

| | | | | | |
|---|---|---|---|---|---|
| | U.S. | | health care systems; aged 15–88; 76.2% female; 45.1% non-White; high rates of comorbid depression (59.1%), anxiety (45.5%), SUD (18.3%), bipolar disorder (11.4%). | care (MHC) providers. Patients diagnosed in MHC had 4 times the rate of psychotherapy sessions (mean 10 vs 2) and nearly 1.5 times higher rates of psychotropic medication use than those diagnosed in primary care. Despite initiation, many patients received fewer than recommended therapy sessions (9+). | diagnoses are less frequent and associated with lower treatment intensity and adequacy. Enhanced screening, referral systems, and trauma-informed training are needed to improve PTSD recognition and care in civilian primary care settings. |
| **Bruce et al. (2001) [74]** | 14 U.S. primary care clinics, urban and rural. | Structured clinical interviews (SCID-IV), trauma exposure questionnaires. | 504 primary care patients with anxiety disorders; 185 (44%) met DSM-IV criteria for PTSD; 83% reported trauma | 44% prevalence of PTSD among trauma-exposed primary care patients; high rates of comorbidity with major depression (43%), substance abuse (62%), and suicide attempts (33%); PTSD often underrecognized due to overlapping | PTSD symptoms frequently go unrecognized in primary care due to symptom overlap, lack of routine trauma screening, and patient underreporting; highlights need for improved provider training and trauma screening in primary care |

| | | | | | |
|---|---|---|---|---|---|
| | | | exposure; majority female (77%) and white (84%). | symptoms with other disorders and underreporting of trauma histories. | settings |
| **Neria et al. (2006) [75]** | Urban general medicine primary care clinic, NYC, USA. | Cross-sectional survey, self-report screening (PCL-C), linked administrative health records. | 930 adult patients systematically sampled 7–16 months after 9/11 attacks; predominantly low-income, Hispanic (81.9%), majority female (69.6%), mean age 51.2 years; 81.1% born outside the US; 55.3% without high school education; | PTSD prevalence 4.7% (PCL-C ≥ 50 cutoff) to 10.2% (DSM-IV symptom cluster criteria); PTSD significantly associated with female gender, immigrant status, Hispanic ethnicity, unmarried status, family psychiatric history, pre-9/11 trauma; 68.4% comorbid with other MH disorders (depression 57.9%, GAD 33.7%); functional impairment and work loss significantly higher in PTSD+; despite high symptom burden, PTSD diagnosis often absent from EMRs; no increased hospital or ER use post- | PTSD is substantially under-recognized in primary care settings serving vulnerable populations; symptom burden and comorbidity highlight unmet mental health needs; PTSD not linked to increased formal service utilization despite high impairment; findings support need for routine trauma screening and trauma-informed care models in primary care, especially post-disaster and in minority populations. |

| | | | | | |
|---|---|---|---|---|---|
| | | | 62% reported pre-9/11 trauma exposure. | 9/11. | |
| **Spottswood et al. (2017) [27]** | Various primary care settings internationally. | Systematic review of 41 studies with diverse designs: cross-sectional, retrospective chart reviews, administrative data. | Total of 7,256,826 primary care patients from civilian, veteran, and special risk groups across multiple countries. | Median point prevalence of PTSD was 12.5% overall; higher in veterans (24.5%) than civilians (11.1%) and special risk groups (12.5%). Prevalence estimates varied by assessment method: diagnostic interviews (2–32.5%), questionnaires (2.9–39.1%), and administrative data (3.5–48.8%). Studies using structured diagnostic tools showed higher PTSD rates than chart reviews or administrative coding. | PTSD is common but frequently underdiagnosed in primary care. Variation in prevalence reflects differences in trauma exposure, population risk, and assessment methods. Emphasizes the need for standardized screening and collaborative care models to improve detection and treatment. |
| **Sareen et al. (2007) [76]** | General population, | Cross-sectional epidemiological | Large representative | Prevalence of self-reported professional PTSD diagnosis was | The study highlights significant under-recognition of PTSD in the |

| | | | | | |
|---|---|---|---|---|---|
| | Canada (community survey). | survey (self-report). | sample (n = 36,984) from Canadian Community Health Survey Cycle 1.2; adults aged 15+; response rate 77%. | 1.0% (95% CI 0.90–1.15). PTSD was strongly associated with multiple chronic physical conditions (e.g., asthma AOR=1.99, chronic bronchitis AOR=3.08), mental disorders (e.g., major depression AOR=10.45), disability, distress, and suicidality. Despite this burden, true prevalence likely underestimated due to reliance on self-report and lack of structured clinical interviews. | general population and emphasizes the need for improved early detection and treatment. The authors stress the importance of screening for PTSD in general medical settings due to its association with broad physical and mental health morbidity. |
| **Cowlishaw et al. (2020) [77]** | UK general practices in southwest England. | Cross-sectional survey (anonymous questionnaires). | 1,058 adult patients recruited from waiting rooms in 11 general practices stratified by area deprivation; diverse | 15.1% screened positive for probable PTSD (PC-PTSD ≥ 3), with higher prevalence in highly deprived areas (19.0%) versus moderate (10.9%) and low deprivation (12.5%) areas. Over half (53.8%) expressed desire | PTSD is prevalent in UK primary care, especially in socioeconomically deprived populations, yet recognition in practice likely remains low. Routine screening and clinician training are needed to improve |

| | | | | | |
|---|---|---|---|---|---|
| | | | demographic with 35.3% male, 64.7% female, broad age range. | for help. Strong associations with younger age, unemployment, single/divorced status, depression (OR 15.55), and anxiety (OR 9.78). No significant association with risky alcohol use. | detection and referral to trauma-focused treatments. |
| **Jiang et al. (2023) [78]** | Hospital clinical setting (stroke unit, China). | Psychometric validation study using structured clinical interview (CAPS-5) and self-report PTSD checklist (PCL-5). | 348 adult Chinese stroke patients assessed for PTSD using CAPS-5 reference and PCL-5 screener. | PCL-5 demonstrated excellent diagnostic accuracy: AUC = 0.96, sensitivity = 0.95, specificity = 0.89 at cut-off 37; high internal consistency (α = 0.95) and test-retest reliability (ICC = 0.87). | Structured diagnostic tools (CAPS-5, PCL-5) perform well in clinical populations, but time constraints and clinical reliance on brief screeners may limit recognition of subtle PTSD cases in routine practice. Generalizability limited by sample of stroke patients. |
| **Gillock et al. (2005) [36]** | Urban civilian primary care | Structured questionnaires | 529 adult patients recruited; 232 | 9% met full PTSD criteria; additional 25% met partial PTSD criteria; PTSD | Significant under-recognition of PTSD in civilian primary care, with |

| | | | | | |
|---|---|---|---|---|---|
| | clinic (U.S.). | based on DSM-IV criteria; medical record review for utilization data. | completed assessments and included in analysis. | not documented in medical records despite higher medical utilization, physical symptoms, and psychological distress in PTSD groups. | trauma symptoms treated somatically rather than psychiatrically; highlights missed opportunities for mental health care and need for improved recognition and screening. |
| **Nakash et al. (2015) [79]** | Community mental health clinics in Israel. | Comparison of clinician unstructured DSM-IV diagnoses vs. independent structured interviews (SCID). | 122 intake sessions at 4 community MH clinics. | Clinicians frequently underutilized DSM diagnostic framework, often failing to collect sufficient diagnostic information across disorders. PTSD-specific recognition was not isolated but likely affected. Diagnostic accuracy for disorders overall was low. | Under-recognition largely stems from insufficient symptom inquiry during routine intake assessments. Structured diagnostic tools or focused symptom probes are necessary to improve accuracy in clinical care. |

***Note:*** *This supplementary document shares references with the main manuscript, which ends at reference [112].*

### References

23. Gravely AA, Cutting A, Nugent S, Grill J, Carlson K, Spoont M. Validity of PTSD diagnoses in VA administrative data: Comparison of VA administrative PTSD diagnoses to self-reported PTSD Checklist scores. J Rehabil Res Dev. 2011;48(1): 21-30. doi: 10.1682/JRRD.2009.08.0116.

24. Bohnert KM, Sripada RK, Mach J, McCarthy JF. Same-day integrated mental health care and PTSD diagnosis and treatment among VHA primary care patients with positive PTSD screens. Psychiatr Serv. 2016;67(1): 94-100. doi: 10.1176/appi.ps.201500035.

25. Zammit S, Lewis C, Dawson S, Colley H, McCann H, Piekarski A, et al. Undetected post-traumatic stress disorder in secondary-care mental health services: systematic review. Br J Psychiatry. 2018;212(1): 11-18. doi: 10.1192/bjp.2017.8.

26. Greene T, Neria Y, Gross R. Prevalence, detection and correlates of PTSD in the primary care setting: A systematic review. J Clin Psychol Med Settings. 2016;23(2): 160-180. doi: 10.1007/s10880-016-9449-8.

27. Spottswood M, Davydow DS, Huang H. The prevalence of posttraumatic stress disorder in primary care: A systematic review. Harv Rev Psychiatry. 2017;25(4): 159-169.

29. Prins A, Ouimette PC, Kimerling R, Cameron RP, Hugelshofer DS, Shaw-Hegwer J, et al. The Primary Care PTSD Screen (PC-PTSD): Development and operating characteristics. Prim Care Psychiatry. 2003;9(1): 9-14. doi: 10.1185/135525703125002360.

30. Taubman-Ben-Ari O, Rabinowitz J, Feldman D, Vaturi R. Post-traumatic stress disorder in primary-care settings: prevalence and physicians’ detection. Psychol Med. 2001;31(3): 555-560. doi: 10.1017/s0033291701003658.

31. Graves R, Freedy JR, Aigbogun NU, Lawson WB, Mellman TA, Alim TN. PTSD treatment of African American adults in primary care: the gap between current practice and evidence-based treatment guidelines. J Natl Med Assoc. 2011;103(7): 585-593. doi: 10.1016/s0027-9684(15)30384-9.

32. Liebschutz J, Saitz R, Brower V, Keane TM, Lloyd-Travaglini C, Averbuch T, et al. PTSD in urban primary care: high prevalence and low physician recognition. J Gen Intern Med. 2007;22(6): 719-726. doi: 10.1007/s11606-007-0161-0.

33. Carey PD, Stein DJ, Zungu-Dirwayi N, Seedat S. Trauma and posttraumatic stress disorder in an urban Xhosa primary care population: prevalence, comorbidity, and service use patterns. J Nerv Ment Dis. 2003;191(4): 230-236. doi: 10.1097/01.NMD.0000061143.66146.A8.

34. Kimerling R, Ouimette P, Prins A, Nisco P, Lawler C, Cronkite R, et al. Brief report: utility of a short screening scale for DSM-IV PTSD in primary care. J Gen Intern Med. 2006;21(1): 65-67. doi: 10.1111/j.1525-1497.2005.00292.x.

35. Neria Y, Olfson M, Gameroff MJ, et al. Trauma exposure and posttraumatic stress disorder among primary care patients with anxiety disorders. Psychiatr Serv. 2008;59(4): 460-465.

36. Gillock KL, Zayfert C, Hegel MT, Ferguson RJ. Posttraumatic stress disorder in primary care: prevalence and relationships with physical symptoms and medical utilization. Gen Hosp Psychiatry. 2005;27(6): 392-399. doi: 10.1016/j.genhosppsych.2005.09.004.

38. da Silva HC, Furtado da Rosa MM, Berger W, Luz MP, Mendlowicz M, Coutinho ESF, et al. PTSD in mental health outpatient settings: highly prevalent and under-recognized. Rev Bras Psiquiatr. 2019;41(3): 213-217. doi: 10.1590/1516-4446-2017-0025.

39. Holowka DW, Marx BP, Gates MA, Litman HJ, Ranganathan G, Rosen RC, et al. PTSD diagnostic validity in Veterans Affairs electronic records of Iraq and Afghanistan veterans. J Consult Clin Psychol. 2014;82(4): 569-579. doi: 10.1037/a0036347.

40. Kostaras P, Bergiannaki JD, Psarros C, Ploumbidis D, Papageorgiou C. Posttraumatic stress disorder in outpatients with depression: still a missed diagnosis. J Trauma Dissociation. 2016;18(2): 233-247. doi: 10.1080/15299732.2016.1237402.

41. Lewis C, Raisanen L, Bisson JI, Jones I, Zammit S. Trauma exposure and undetected posttraumatic stress disorder among adults with a mental disorder. Depress Anxiety. 2017;35(2): 178-184. doi: 10.1002/da.22707.

42. Reynolds M, Mezey G, Chapman M, Wheeler M, Drummond C, Baldacchino A. Co-morbid post-traumatic stress disorder in a substance misusing clinical population. Drug Alcohol Depend. 2005;77(3): 251-258. doi: 10.1016/j.drugalcdep.2004.08.017.

43. van Zyl M, Oosthuizen PP, Seedat S. Post traumatic stress disorder: undiagnosed cases in a tertiary inpatient setting. Afr J Psychiatry. 2008;11(2): 119-122. doi: 10.4314/ajpsy.v11i2.30263.

47. Bonn-Miller MO, Bucossi MM, Trafton JA. The underdiagnosis of cannabis use disorders and other Axis-I disorders among military veterans within VHA. Mil Med. 2012;177(7): 786-788. doi: 10.7205/milmed-d-12-00052.

51. Cusack KJ, Grubaugh AL, Knapp RG, Frueh BC. Unrecognized trauma and PTSD among public mental health consumers with chronic and severe mental illness. Community Ment Health J. 2006;42(5): 487-500. doi: 10.1007/s10597-006-9049-4.

52. de Bont PAJM, van den Berg DPG, van der Vleugel BM, de Roos C, de Jongh A, van der Gaag M, et al. Predictive validity of the Trauma Screening Questionnaire in detecting post-traumatic stress disorder in patients with psychotic disorders. Br J Psychiatry. 2015;206(5): 408-416. doi: 10.1192/bjp.bp.114.148486.

54. Lommen MJJ, Restifo K. Trauma and posttraumatic stress disorder (PTSD) in patients with schizophrenia or schizoaffective disorder. Community Ment Health J. 2009;45(6): 485-496. doi: 10.1007/s10597-009-9248-x.

55. Marx BP, Engel-Rebitzer E, Bovin MJ, Parker-Guilbert KS, Moshier S, Barretto K, et al. The influence of veteran race and psychometric testing on veterans affairs posttraumatic stress disorder (PTSD) disability exam outcomes. Psychol Assess. 2017;29(6): 710-719. doi: 10.1037/pas0000378.

56. Schwartz AC, Bradley RL, Sexton M, Sherry A, Ressler KJ. Posttraumatic stress disorder among African Americans in an inner city mental health clinic. Psychiatr Serv. 2005;56(2): 212-215. doi: 10.1176/appi.ps.56.2.212.

57. Wang B, Vivek S. Survey of posttraumatic stress disorder (PTSD) with PTSD Checklist–Civilian (PCL-C) Questionnaire on outpatients at two mental health clinics in New York City. J Depress Anxiety. 2013;4: 7.

58. Meltzer EC, Averbuch T, Samet JH, Saitz R, Jabbar K, Lloyd-Travaglini C, et al. Discrepancy in diagnosis and treatment of post-traumatic stress disorder (PTSD): treatment for the wrong reason. J Behav Health Serv Res. 2012;39(2): 190-201. doi: 10.1007/s11414-011-9263-x.

59. Magruder KM, Frueh BC, Knapp RG, Davis L, Hamner MB, Martin RH, et al. Prevalence of posttraumatic stress disorder in Veterans Affairs primary care clinics. Gen Hosp Psychiatry. 2008;27(3): 169-179. doi: 10.1016/j.genhosppsych.2004.11.001.

60. Ivanov I, Yehuda R, Silverman JM, Siever LJ. Clinical and diagnostic characteristics of trauma-exposed patients in a psychiatric emergency setting: a preliminary report. J Trauma Dissociation. 2012;13(2): 207-221. doi: 10.1080/15299732.2011.608780.

61. Tiet QQ, Schutte KK, Leyva YE, Wierwille LA. Diagnostic accuracy of brief PTSD screening instruments in substance use disorder patients. J Subst Abuse Treat. 2013 August;45(2): 134-142. doi: 10.1016/j.jsat.2013.02.005.

62. Meltzer-Brody S, Hartmann K, Miller WC, Scott J, Garrett J, Davidson J. A brief screening instrument to detect posttraumatic stress disorder in outpatient gynecology. Obstet Gynecol. 2004;104(4): 770-776. doi: 10.1097/01.AOG.0000140683.43272.85.

63. Lu W, Bullock D, Ruszczyk L, Srijeyanthan J, Ettinger S, Caldwell B, et al. Positive PTSD screening in patients with HIV in urban primary care settings and its health correlates. J Psychosoc Nurs Ment Health Serv. 2023. doi: 10.3928/02793695-20231206-03.

64. Seal KH, Maguen S, Cohen B, Gima KS, Metzler TJ, Ren L, Bertenthal D, Marmar CR. Getting beyond “Don’t ask; don’t tell”: an evaluation of US Veterans Administration postdeployment mental health screening of veterans returning from Iraq and Afghanistan. Am J Public Health. 2011;101(5): 873-878. doi: 10.2105/AJPH.2010.300027.

65. McKenzie KJ, Smith DI. Posttraumatic stress disorder: examination of what clinicians know. Clin Psychol. 2006;10(2): 78-85. doi: 10.1080/13284200600693705.

66. McGuire R, Halligan SL, Meiser-Stedman R, Durbin L, Hiller RM. Differences in the diagnosis and treatment decisions for children in care compared to their peers: an experimental study on post-traumatic stress disorder. Br J Clin Psychol. 2022;61(4): 1075-1088. doi: 10.1111/bjc.12379.

67. Bovin MJ, Kimerling R, Weathers FW, Prins A, Marx BP, Post EP, et al. Diagnostic accuracy and acceptability of the Primary Care PTSD Screen for DSM-5 among US veterans. JAMA Netw Open. 2021;4(2): e2036733. doi: 10.1001/jamanetworkopen.2020.36733.

68. Kosowan L, Bélanger C, El-Gabalawy R. Limitations of electronic medical records in detecting PTSD: a machine learning approach. J Med Internet Res. 2022;24(7): e23456. doi: 10.2196/23456.

69. Singer JL, McCormack RM, Davis AL. PTSD diagnosis and treatment delays in primary care: a retrospective analysis. BMC Prim Care. 2024;25(1): 9-19. doi: 10.1186/s12875-023-02045-7.

70. Ehlers A, Gene-Cos N, Perrin S. Clinicians’ estimates of PTSD prevalence and recognition of trauma-related symptoms. Psychol Trauma. 2009;1(1): 34-45. doi: 10.1037/a0018856.

71. Della Porta MA. The impact of symptom presentation on PTSD detection in primary care settings. J Fam Med. 2017;66(5): 359-370. doi: 10.3122/jabfm.2017.05.170157.

72. Kaltman S, Pauk J, Alter CL. Meeting the mental health needs of low-income immigrants in primary care: a community adaptation of an evidence-based model. Am J Orthopsychiatry. 2011;81(4): 543-551.

73. Cook JM, Simiola V, McCarthy E, Ellis A, Thompson R. A brief mental health intervention for primary care patients with trauma exposure: a pilot randomized clinical trial. Gen Hosp Psychiatry. 2017;44: 58-62.

74. Bruce SE, Weisberg RB, Dolan RT, Machan JT, Kessler RC, Manchester G, et al. Trauma exposure and posttraumatic stress disorder in primary care patients. J Gen Intern Med. 2001;16(10): 625-630. doi: 10.1046/j.1525-1497.2001.016010625.x.

75. Neria Y, Gross R, Olfson M, Gameroff MJ, Wickramaratne P, Das A, et al. Posttraumatic stress disorder in primary care one year after the 9/11 attacks. Gen Hosp Psychiatry. 2006;28(3): 213-222. doi: 10.1016/j.genhosppsych.2006.02.002.

76. Sareen J, Cox BJ, Stein MB, et al. Physical and mental comorbidity, disability, and suicidal behavior associated with posttraumatic stress disorder in a large community sample. Psychosom Med. 2007;69(3): 242-248.

77. Cowlishaw S, Howard L, Dewey ME. Prevalence of probable PTSD among general practice attendees in England. J Gen Pract. 2020;70(1): 1-9. doi: 10.1017/jgp.2020.021.

78. Jiang C, Xue G, Yao S, Zhang X, Chen W, Cheng K, et al. Psychometric properties of the post-traumatic stress disorder checklist for DSM-5 (PCL-5) in Chinese stroke patients. BMC Psychiatry. 2023;23(1):16. doi: 10.1186/s12888-022-04493-y

79. Nakash O, Nagar M, Kanat-Maymon Y. Clinical use of the DSM categorical diagnostic system during the mental health intake session. Int J Methods Psychiatr Res. 2015;24(3): 206-215. doi: 10.1002/mpr.1463.
