## Supplemental Table 2 for "Post-traumatic stress disorder diagnostic accuracy rates in clinical settings: a systematic review and meta-analysis"

### TABLE S2: RISK OF BIAS ASSESSMENTS OF THE INCLUDED STUDIES.

***Note:*** *This supplementary document shares references with the main manuscript, which ends at reference [111]. Three new references are cited only in this document and are numbered [112, 113, 114].*

Table S2.1 Risk of bias assessment of the included observational studies using A Cochrane Risk of Bias Assessment Tool: for Non-Randomized Studies of Interventions (ACROBAT-NRSI, now referred to as ROBINS-I), [44].[1]

**OBSERVATIONAL STUDIES**

| **STUDY ID** | **Risk of bias due to confounding** —This domain assesses | **Risk of bias in classification of interventions** — This | **Risk of bias in selection of participants into the study (or into the** | **Risk of bias due to deviations from intended interventions** | **Risk of bias due to missing data** — This domain | **Risk of bias arising from measurement of the outcome** — | **Risk of bias in selection of the reported result** —This domain | **Overall** |
|---|---|---|---|---|---|---|---|---|

[1] *Note: ChatGPT [112], an AI language model developed by OpenAI, was utilized to assist in the risk of bias assessment of the included studies using A Cochrane Risk of Bias Assessment Tool: for Non-Randomized Studies of Interventions (ACROBAT-NRSI now referred to as ROBINS-I), [44]. Each individual study was provided for review and assessment. The initial prompt used was "Can you conduct a risk of bias assessment of the included observational studies using A Cochrane Risk of Bias Assessment Tool: for Non-Randomized Studies of Interventions (ACROBAT-NRSI)?" This prompt was subsequently revised to explicitly reference ROBINS-I to reflect updated terminology and guidance. AI-generated responses were carefully reviewed, edited, and verified for accuracy before incorporation into the overall evidence synthesis. The AI's responses were reviewed, revised, and ensured for accuracy before incorporation into the overall analysis of the evidence [112].*

| | | | | | | |
|---|---|---|---|---|---|---|
| whether other variables (confounders) could have distorted the relationship between the exposure or intervention and the outcome. | domain evaluates whether interventions or exposures were clearly defined, measured accurately, and classified correctly for all participants. | **analysis)** — This domain considers whether the way participants were included could have introduced systematic differences between groups or influenced results. | — This domain examines whether any departures from planned interventions or exposures occurred and could have affected study outcomes. | assesses whether incomplete or missing data on outcomes, exposures, or confounders could have led to bias. | This domain evaluates whether outcome assessment was reliable, consistent, and blinded when appropriate, reducing measurement errors. | assesses whether analyses or results were selectively chosen or reported based on the findings. |

| | | | | | | | | |
|---|---|---|---|---|---|---|---|---|
| **Bonn-Miller et al. (2012) [47]** | **Moderate** — The selection of only veterans with cannabis use disorder could confound the observed detection rate of PTSD, as this subgroup may differ systematically in trauma exposure or | **Low** — The "interventions" here were the diagnostic classification methods: structured CAPS interview and EMR records. Both were clearly defined and applied consistently to | **Moderate** — Inclusion required participants to be motivated to quit cannabis and consent to assessment, which likely created a non-representative sample of all VA clinic patients. | **Low** — No deviations occurred because the study did not deliver interventions; it only assessed diagnoses in a single cross-sectional visit. | **Low** — All participants completed CAPS assessments and EMR data were available for all cases; no missing data were reported. | **Moderate** — CAPS interviews were standardized, but EMR diagnoses depended on clinician documentation practices, which may have been inconsistent or incomplete, | **Low** — The study reported all planned outcomes (CAPS prevalence and EMR prevalence), and there was no evidence of selective reporting. | **Moderate** — Confounding, participant selection, and EMR measurement limitations likely introduced moderate bias overall. |

| | | | | | | | | |
|---|---|---|---|---|---|---|---|---|
| | diagnosis likelihood compared to other clinical populations. No adjustment for confounding variables (e.g., comorbidities, treatment engagement) was reported. | all participants. | | | | risking misclassification of PTSD. | | |
| **Cusack et al. (2006) [51]** | **Moderate** — The sample | **Moderate** — PTSD | **Low** — All consumers | **Low** — No interventions | **Low** — No missing | **Moderate** — PTSD | **Low** — All planned | **Moderate** — Use of self-report |

| | | | | | | | | |
|---|---|---|---|---|---|---|---|---|
| | included individuals with severe mental illness, which may be associated both with higher trauma exposure and with under-recognition of PTSD. No adjustment for potential confounders | diagnosis was based on a self-report checklist (PCL) without structured diagnostic interviews, raising potential misclassification of PTSD status. EMR documentation may also have | enrolled in the program were invited, with no exclusions reported other than refusal to participate. The sample appears representative of that setting. | were delivered. This was a cross-sectional assessment study. No deviations occurred. | outcome data were reported; all participants completed the PCL and EMR records were available. | measurement relied entirely on self-reported symptoms without clinician confirmation, increasing the risk of over- or underestimation. EMR records may have under-documented | outcomes and analyses were reported. No evidence of selective reporting. | measures and potential confounding factors likely introduced moderate bias overall. |

| | | | | | | | | |
|---|---|---|---|---|---|---|---|---|
| | such as comorbid substance use, symptom severity, or treatment engagement was reported. | been inconsistent. | | | | trauma exposure and PTSD diagnoses. | | |
| **da Silva et al. (2019) [38]** | **Moderate** — Patients were from a tertiary care psychiatric clinic without adjustment for | **Moderate** — PTSD diagnosis by SCID-IV is strong, but lack of inter-rater reliability | **Moderate** — Participation was voluntary, with ~60% nonparticipatio n; refusal was likely higher | **Low** — No interventions or deviations occurred; purely observational. | **Low** — All outcome data were available and accounted for. | **Moderate** — SCID-IV interviews were supervised but had no formal reliability | **Low** — All planned outcomes were reported transparently. | **Moderate** — Confounding, selection bias, and measurement limitations were present but methods were |

| | | | | | | | | |
|---|---|---|---|---|---|---|---|---|
| | comorbidity or severity, increasing potential confounding. | checks and variable EMR documentation by trainees could lead to misclassification. | among trauma-affected patients, introducing selection bias. | | | testing; EMR diagnoses by trainees could be inconsistent. | | otherwise rigorous. |
| **de Bont et al. (2015) [52]** | **Moderate** — Participants were patients with psychotic disorders recruited in long-term mental | **Low** — PTSD was diagnosed with the CAPS (gold standard) and psychotic disorder confirmed by MINI-Plus, | **Moderate** — Participation required consent at multiple stages; 51% of eligible patients either | **Low** — No interventions were delivered—this was a cross-sectional diagnostic | **Low** — All screening and diagnostic data were either available or accounted for, with multiple imputation | **Low** — PTSD was assessed by trained clinicians using the CAPS with established psychometric | **Low** — All prespecified outcomes, including PTSD prevalence, TSQ accuracy, and | **Moderate** — While measurement was rigorous, moderate risk remains due to confounding and selection bias |

| | | | | | | | | |
|---|---|---|---|---|---|---|---|---|
| | healthcare services. PTSD prevalence and detection may be influenced by unmeasured confounders (e.g., illness severity, service engagement, trauma type) not adjusted for in | with validated instruments and clear definitions. | declined or were not reached for interviews, which may systematically exclude less engaged or more symptomatic individuals. | validation study. | used to estimate prevalence. | properties and independent interviews; minimal measurement bias risk. | comparison to EMR documentation , were reported transparently. | from voluntary participation and unadjusted comparisons. |

| | | | | | | | | |
|---|---|---|---|---|---|---|---|---|
| | estimating prevalence. | | | | | | | |
| **Holowka et al. (2014) [39]** | **Moderate** — Participants were oversampled to include those with prior EMR PTSD diagnoses and those without. Although this improves statistical | **Low** — PTSD diagnosis was assessed by SCID (gold standard) with excellent inter-rater reliability (κ = .87–.91) and clear EMR coding criteria. Classifications were clearly defined and | **Moderate** — Recruitment was based on EMR records, including those with ≥2 PTSD diagnoses and a comparison group without. Only ~38% of contacted veterans | **Low** — No intervention was administered —cross-sectional diagnostic validation only. No deviations. | **Low** — Minimal missing data (e.g., 176 participants excluded from final models due to incomplete covariate data), all accounted for and handled | **Low** — SCID interviews were conducted by doctoral-level clinicians blind to EMR status, with high inter-rater reliability. Minimal measurement error risk. | **Low** — All prespecified outcomes (sensitivity, specificity, predictors of concordance) were reported transparently. No evidence of selective reporting. | **Moderate** — Selection bias and residual confounding present, but measurement and reporting were rigorous. |

| | | | | | |
|---|---|---|---|---|---|
| | power, it introduces confounding related to treatment seeking and symptom severity, which were not adjusted for in estimates of concordance. | consistently applied. | completed all assessments, with likely systematic differences in engagement and symptom severity. | | appropriately (multiple imputation sensitivity analyses). |

| | | | | | | | | |
|---|---|---|---|---|---|---|---|---|
| **Lommen and Restifo (2009) [54]** | **Moderate** — Participants were outpatients with schizophrenia or schizoaffective disorder, and no adjustment was made for comorbidity, symptom severity, or treatment | **Moderate** — PTSD status was determined using self-report measures (PSS-SR), not a structured diagnostic interview. Although the scales have good psychometric | **Moderate** — Therapists pre-screened patients and excluded many (e.g., too symptomatic, unable to consent). 75 were selected, and only 33 participated, creating likely systematic exclusion of | **Low** — No interventions were conducted—this was a cross-sectional assessment only. | **Low** — No missing outcome data were reported; all participants who consented completed measures. | **Moderate** — All measures were read aloud to mitigate comprehensio n problems, but reliance solely on self-report (PSS-SR) without clinician confirmation limits accuracy. | **Low** — All prespecified outcomes—trauma prevalence, PTSD rates, negative cognitions—were reported transparently. | **Moderate** — Confounding, self-report measurement, and selection bias were present despite otherwise transparent methods. |

| | | | | | | | | |
|---|---|---|---|---|---|---|---|---|
| | engagement, all of which can affect PTSD prevalence and reporting. | properties, reliance on self-report increases misclassification risk. | those with higher distress or cognitive impairment. | | | Authors note this limitation. | | |
| **Marx et al. (2017) [55]** | **Moderate** — The study adjusted for many demographic and clinical factors (e.g., age, gender, income, | **Low** — SCID-based PTSD diagnosis was conducted by trained doctoral-level assessors with excellent interrater | **Moderate** — Participants were drawn from Project VALOR with oversampling of PTSD cases, and the final analytic | **Low** — No interventions were delivered in this observational comparison of diagnostic outcomes. | **Low** — Missing data were minimal; the study reported exclusions transparently and performed complete-case | **Low** — The SCID was administered by blinded, trained clinicians; C&P diagnoses were extracted | **Low** — All planned outcomes and subgroup analyses were clearly pre-specified and reported transparently | **Moderate** — While measurement quality and reporting were excellent, moderate risk remains due to potential |

| | | | | | | | | |
|---|---|---|---|---|---|---|---|---|
| | combat exposure, education), but important unmeasured confounders (e.g., examiner attitudes, regional policies, time since trauma) could still influence diagnostic concordance | reliability (κ = .91) blinded to C&P outcomes and race; C&P diagnoses were abstracted reliably from EMRs. | sample was restricted to those with available C&P exams and who were White or Black, potentially introducing selection bias affecting representativeness. | | analysis without substantial loss of cases within the analytic sample. | systematically. Discordance is likely due to true differences in assessment approaches rather than measurement error. | (e.g., concordance rates, impact of psychometric testing, racial disparities). | unmeasured confounding and selection processes related to the sampling frame and exclusions. |

| | | | | | | | | |
|---|---|---|---|---|---|---|---|---|
| | and racial disparities. | | | | | | | |
| **Schwartz et al. (2005) [56]** | **Moderate** — Participants were long-term mental health outpatients with very high trauma exposure, high rates of poverty, and comorbid disorders. The | **Moderate** — PTSD was primarily measured by self-report (PSS), with the SCID administered only to a randomly selected subset (72/184). The use of self- | **Moderate** — Participants were selected from a single urban clinic, required long-term engagement (≥2 years), and 8% declined or were ineligible. Selection | **Low** — No interventions were delivered. | **Low** — Missing data were minimal (6 unavailable charts), and all exclusions were accounted for transparently. | **Moderate** — The PSS was administered by interviewers to improve comprehension, but reliance on self-report and inconsistent application of SCID across | **Low** — All prespecified outcomes—PTSD prevalence, trauma exposure, comorbidity, and detection rates—were reported transparently. | **Moderate** — Measurement and confounding bias due to reliance on self-report and unadjusted comparisons, despite transparent reporting and good SCID |

| | | | | | | | | |
|---|---|---|---|---|---|---|---|---|
| | study did not adjust estimates for comorbid substance use or illness severity, which can influence detection and PTSD rates. | report scales for the main prevalence estimate and reliance on chart documentation without standardized diagnostic procedures introduces potential misclassification. | could bias results if more severe cases were excluded due to cognitive impairment or refusal. | | | participants increases measurement risk. | | subsample procedures. |

| | | | | | | | | |
|---|---|---|---|---|---|---|---|---|
| **Wang and Vivek (2013) [57]** | **Moderate** — The study included psychiatric outpatients from low-income, inner-city clinics with high trauma exposure and comorbidities. No adjustment for illness severity, | **Moderate** — PTSD was measured using the PCL-C self-report checklist, not a structured clinical interview (CAPS). The authors explicitly note this limitation and recommend | **Moderate** — Of eligible patients, only 62 consented to participate due to time-consuming consent procedures and voluntary recruitment. Selection likely favored patients more willing to | **Low** — No interventions were delivered; this was a cross-sectional survey and chart review. | **Low** — No missing outcome data were reported; all participants completed both questionnaires and chart reviews. | **Moderate** — The PCL-C was self-administered and interpreted without clinician confirmation. EMR diagnoses relied on routine documentation, which is known to be | **Low** — All planned outcomes (PTSD prevalence, trauma exposure, comparisons to charts) were reported transparently. | **Moderate** — While reporting and completeness were good, reliance on self-report and selection bias likely introduced moderate overall bias. |

| | | | | | | | | |
|---|---|---|---|---|---|---|---|---|
| | comorbidity, or demographic confounders was performed when comparing survey-based PTSD prevalence to EMR documentation . | caution interpreting prevalence estimates. | discuss trauma or PTSD symptoms. | | | inconsistent. This increases risk of outcome misclassificati on. | | |
| **van Zyl et al. (2008) [43]** | **Moderate** — Participants | **Low** — PTSD was diagnosed | **Moderate** — Patients were | **Low** — No interventions | **Low** — No missing | **Low** — CAPS was | **Low** — All planned | **Moderate** — Despite strong |

| | | | | | | | | |
|---|---|---|---|---|---|---|---|---|
| | were inpatients admitted for mood and anxiety disorders, with high trauma exposure and comorbidity. No adjustment was made for illness severity, prior treatment, or other variables influencing | using the CAPS administered by trained clinicians, considered the gold standard. Prior diagnoses were obtained from medical records. Classification was clearly defined and | randomly selected from a therapeutic ward, but only those able to consent and without recent substance dependence were included. This likely excluded more symptomatic or cognitively | were delivered—this was a cross-sectional diagnostic assessment. | outcome data were reported. All 40 participants completed the CAPS assessment and were included in analysis. | administered by trained interviewers following DSM-IV criteria. Minimal measurement bias is likely. | outcomes—PTSD prevalence, demographic comparisons, substance use associations—were reported transparently. | measurement procedures, moderate bias remains due to unadjusted confounding and selection limitations. |

| | | | | | | | | |
|---|---|---|---|---|---|---|---|---|
| | PTSD detection and reporting. | applied consistently. | impaired individuals. | | | | | |
| **Reynolds et al. (2005) [42]** | **Moderate** — No adjustment was made for comorbid psychiatric conditions, trauma severity, or duration of abstinence—all of which could | **Moderate** — PTSD was assessed using the PSS-I (interview version), a validated but brief measure rather than a structured diagnostic interview (e.g., | **Moderate** — Participants were recruited only if they were abstinent for at least 18 days and able to consent, excluding those with severe withdrawal, | **Low** — No interventions were delivered—this was a cross-sectional assessment of trauma and PTSD prevalence. | **Low** — No missing outcome data reported; all consenting participants completed assessments. | **Moderate** — PTSD diagnoses were based on the PSS-I interview without independent clinician verification, and chart diagnoses | **Low** — All planned outcomes—prevalence estimates, trauma characteristics, impact ratings—were transparently reported. | **Moderate** — While reporting and sample documentation were strong, moderate bias arises from selection procedures, unadjusted confounding, and reliance on a |

| | | | | | | | | |
|---|---|---|---|---|---|---|---|---|
| | influence PTSD detection and reporting. The sample consisted of inpatients in detoxification/ rehabilitation, a group with very high psychiatric morbidity. | SCID or CAPS). Authors explicitly note this limitation. EMR diagnoses were based on unstandardized clinician notes. | cognitive impairment, or acute psychosis, likely biasing the sample. | | | were inconsistent. The authors note under-detection in charts and potential misclassification. | | brief PTSD measure rather than a structured diagnostic interview. |
| **Lewis et al. (2017) [41]** | **Moderate** — No | **Moderate** — PTSD was | **Moderate** — Participants | **Low** — No intervention | **Low** — All respondents | **Moderate** — TSQ scores | **Low** — All planned | **Moderate** — While |

| | | | | | | | | |
|---|---|---|---|---|---|---|---|---|
| | multivariate adjustment for severity of primary mental disorders, comorbidities, or treatment status when estimating undetected PTSD rates. Authors note confounding by symptom | assessed using the TSQ, a validated screening tool rather than a structured diagnostic interview (CAPS or SCID). Self-reported prior PTSD diagnoses may underreport true EMR- | were recruited through mixed systematic (NHS) and non-systematic (advertising) approaches. Only 57% returned questionnaires, with likely systematic differences in who responded. | was delivered—this was purely observational. | who returned the TSQ completed all outcome items. The main issue was nonresponse (43% did not return the questionnaires), but this was accounted for in reporting. | relied on self-report anchored to a single traumatic event. No structured interview confirmation. Potential misclassification acknowledged by the authors. | outcomes—prevalence, trauma types, demographic predictors—were transparently reported. | measurement, classification, and response bias were acknowledged and methods were clearly reported, reliance on self-report and selection limitations result in moderate overall risk. |

| | | | | | | | | |
|---|---|---|---|---|---|---|---|---|
| | overlap and demographic factors (e.g., income, gender). | documented diagnoses. | | | | | | |
| **Meltzer et al. (2012) [58]** | **Moderate** — No multivariate adjustment for trauma severity or other psychiatric comorbidities beyond | **Low** — PTSD diagnosis was determined via the CIDI, a structured diagnostic interview administered by trained research | **Moderate** — Participants were recruited in a single urban safety-net primary care setting. Although 81% of eligible patients | **Low** — No interventions were delivered; the study was cross-sectional and observational. | **Low** — Minimal missing data: all 133 participants with PTSD completed diagnostic interviews and | **Low** — PTSD was diagnosed with a validated structured interview (CIDI). EMR documentation was systematic, | **Low** — All pre-specified outcomes (PTSD prevalence, MH treatment receipt, predictors) were reported transparently. | **Moderate** — While measurement was robust, moderate risk arises due to unadjusted confounding and selection limitations |

| | | | | | | | | |
|---|---|---|---|---|---|---|---|---|
| | depressive symptoms, which may affect detection and treatment rates. The authors note that cross-sectional design prevents clarifying causality and | assistants. EMR diagnoses were extracted systematically. | consented, those declining may differ systematically. Additionally, inclusion was limited to English speakers. | | EMR abstraction. | though content of MH visits was not available. Misclassification is unlikely in PTSD diagnosis itself. | | inherent to the study setting. |

| | | | | | | | | |
|---|---|---|---|---|---|---|---|---|
| | residual confounding. | | | | | | | |
| **Kostaras et al. (2016) [40]** | **Moderate** — No multivariate adjustment for illness severity or comorbid disorders when estimating PTSD prevalence in MDD. Although logistic | **Low** — PTSD diagnosis was made using the MINI structured interview administered by a trained psychologist blinded to other assessments. Trauma | **Moderate** — Recruited from two outpatient clinics in a single university hospital. Only about two-thirds of eligible patients consented (112/164), and | **Low** — No interventions were applied—this was a cross-sectional observational assessment. | **Low** — All 101 participants completed the diagnostic interview and trauma assessments. Minimal missing data. | **Low** — PTSD and MDD diagnoses were based on the MINI, administered consistently by trained raters. EMR verification was conducted, and under- | **Low** — All planned outcomes—lifetime and current PTSD prevalence, trauma history, predictors—were reported transparently. | **Moderate** — Although measurement was rigorous, moderate risk arises from unadjusted confounding and selection bias. |

| | | | | | | |
|---|---|---|---|---|---|---|
| | regression was used for predictors of comorbidity, the main prevalence estimates were unadjusted. Authors note that prolonged trauma and chronic depression likely confound | exposure was measured systematically with the Life Events Checklist. | 101 completed all criteria. Selection may favor patients more engaged with services. | | | diagnosis was noted transparently. |

| | | | | | | | | |
|---|---|---|---|---|---|---|---|---|
| | observed associations. | | | | | | | |
| **Magruder et al. (2008) [59]** | **Moderate** — While analyses adjusted for several patient-level covariates (age, sex, race, war-zone service), no adjustment was made for other psychiatric | **Low** — PTSD diagnosis established via the Clinician-Administered PTSD Scale (CAPS), administered by trained blinded interviewers, considered a gold standard. | **Moderate** — Although the sample was randomly selected and oversampled for women, only 73% of invited patients consented and about 56% (819/1,474) | **Low** — No intervention was applied—purely observational. | **Low** — Minimal missing data for main outcomes among participants with usable data (819). Those completing interviews had complete | **Low** — PTSD measured by CAPS, administered blinded to PCL results. Provider recognition determined from systematic EMR coding. Both measures | **Low** — All planned outcomes—recognition rates, predictors of recognition, functional status—were transparently reported. | **Moderate** — While measurement and reporting were rigorous, moderate risk arises from confounding (unadjusted comorbidities) and selection (nonresponse and |

| | | | | | | | | |
|---|---|---|---|---|---|---|---|---|
| | comorbidities (e.g., depression) or severity of trauma exposure in estimating recognition rates. Authors noted unmeasured factors likely contributed to differential recognition. | EMR diagnoses were systematically abstracted. | completed usable data. Nonresponse could be related to PTSD severity or willingness to disclose trauma. | | PTSD assessments. | are standardized and validated. | | exclusion criteria). |

| | | | | | | | | |
|---|---|---|---|---|---|---|---|---|
| **Ivanov et al. (2012) [60]** | **Moderate** — No adjustment for psychiatric comorbidities, trauma severity, or socio-demographic factors influencing PTSD detection and recording. The authors note higher PTSD | **Low** — PTSD assessed with the MINI PTSD module, a structured diagnostic interview administered systematically. Although the French version was not formally validated, it was based on a | **Moderate** — 21.6% of consecutive eligible patients were excluded (resident forgot, patient unable to participate, refusal, incomplete data). This high exclusion rate may | **Low** — No interventions were applied; purely observational. | **Low** — Among included patients (N=316), no missing PTSD outcome data were reported. | **Moderate** — Although the MINI is validated overall, the French version was not formally validated, and interviews were administered by residents with variable training. Some | **Low** — All planned outcomes (prevalence, subgroup rates, clinician perceptions) were transparently reported. | **Moderate** — Despite systematic measurement, moderate risk arises from selection bias (high exclusion) and lack of adjustment for confounding. |

| | | | | | | | | |
|---|---|---|---|---|---|---|---|---|
| | prevalence in subgroups (refugees, migrants) but did not adjust for these factors when estimating overall prevalence or comparison to historical records. | validated instrument and applied consistently. | systematically bias prevalence estimates. | | | clinicians noted discomfort with the tool, possibly affecting consistency. | | |
| **Tiet et al. (2013) [61]** | **Moderate** — No adjustment | **Low** — PTSD was assessed | **Moderate** — Participants | **Low** — No intervention | **Low** — All enrolled | **Low** — The C-DIS-IV | **Low** — All prespecified | **Moderate** — While |

| | | | | | | | | |
|---|---|---|---|---|---|---|---|---|
| | for potential confounders affecting PTSD detection (e.g., psychiatric comorbidities, trauma severity, treatment engagement). The study population was veterans already | using the C-DIS-IV structured diagnostic interview administered uniformly to all participants. VA administrative records were extracted systematically. Classification | were recruited consecutively from VA outpatient mental health clinics. Only those who agreed to participate were included; non-responders may have differed systematically | was applied—purely observational assessment. | participants completed both the index and reference assessments with no missing PTSD outcome data reported. | interview is validated and was administered in a standardized manner. VA records were consistently abstracted for documentation of PTSD diagnosis. | outcomes—including PTSD prevalence, detection rates, and symptom severity—were transparently reported. | measurement and reporting were rigorous, moderate risk arises from unadjusted confounding and potential selection bias due to voluntary participation. |

| | | | | | | | | |
|---|---|---|---|---|---|---|---|---|
| | engaged in mental health care, which may overestimate recognition rates compared to general clinical populations. | was consistent and used validated instruments. | in symptom severity or willingness to report trauma. | | | | | |
| **Bohnert et al. (2016) [24]** | **Moderate** — The study used multivariable regression adjusting for | **Low** — Service setting (primary care only, PC-MHI, specialty | **Low** — All patients meeting inclusion criteria within | **Low** — No intervention was experimentally applied. | **Low** — Administrative data provided full follow-up of diagnosis | **Low** — PTSD diagnosis and treatment initiation were captured via | **Low** — All planned outcomes (diagnosis and treatment | **Low to Moderate** — The study was robustly designed and adjusted for |

| | | | | | | | | |
|---|---|---|---|---|---|---|---|---|
| | demographic characteristics, prior psychiatric diagnosis, prior VHA use, and PTSD screen severity. However, unmeasured confounding remains possible (e.g., PTSD | mental health) was derived from standardized administrative data and clearly defined. | the 30% random national sample were included. No evidence of selective inclusion. | Observational study of existing care patterns. | and treatment initiation over 1 year. No missing outcome data. | ICD-9 codes and procedural codes. These measures are standardized in the VA system and consistently applied. | initiation at multiple time points) were transparently reported. No evidence of selective reporting. | many confounders. Residual confounding remains possible, but measurement and reporting were rigorous. |

symptom
severity
beyond the
PC-PTSD
screen,
clinician
judgment
factors, patient
preference).
Authors
themselves
note potential
residual
confounding

| | | | | | | | | |
|---|---|---|---|---|---|---|---|---|
| | despite adjustment. | | | | | | | |
| **Meltzer-Brody et al. (2004) [62]** | **Moderate** — No adjustment for potential confounding factors (e.g., trauma severity, comorbid psychiatric diagnoses, socioeconomic status) influencing | **Low** — Trauma history was self-reported with a clearly defined question; PTSD diagnosis was determined by a structured diagnostic interview | **Serious** — Only 36% (32 of 88) of patients with trauma history completed the structured interview, introducing substantial selection bias. Nonparticipants may differ | **Low** — No intervention was applied; observational survey and diagnostic interview only. | **Moderate** — High dropout rate from eligible trauma-exposed patients (64% did not complete the psychiatric assessment), creating potential bias | **Low** — PTSD diagnosis used a validated structured interview (MINI) administered blinded to SPAN results. | **Low** — All planned outcomes (PTSD prevalence, SPAN performance, comorbid diagnoses) were reported transparently. | **Moderate to Serious** — Although measurement was rigorous, the combination of high nonparticipation and lack of adjustment for confounding creates |

| | | | | | |
|---|---|---|---|---|---|
| | both likelihood of PTSD and willingness to complete the structured interview. Authors noted that women who completed interviews might have more severe symptoms. | (MINI) administered by a blinded psychiatrist. Classification was consistent and clearly reported. | systematically in symptom severity or readiness to disclose trauma. | in estimating PTSD prevalence and recognition. | substantial risk of bias. |

| | | | | | | | | |
|---|---|---|---|---|---|---|---|---|
| **Prins et al. (2003) [29]** | **Moderate** — No adjustment for potential confounding variables (e.g., trauma severity, psychiatric comorbidities, demographic factors) that could influence both PTSD prevalence and | **Low** — PTSD status determined by CAPS structured interview administered by trained assessors blinded to screening results. VA medical record diagnoses | **Moderate** — 56% of invited patients completed the survey, creating potential selection bias. Among screen-positive participants, 79% completed the CAPS interview, | **Low** — No intervention applied; purely observational. | **Low** — Among the 57 who completed the CAPS, outcome data were complete. Medical record data were available for 71% of the total survey participants, but this was | **Low** — CAPS is a validated gold-standard instrument; interviews were blinded. Chart diagnoses were consistently abstracted. High inter-rater reliability reported. | **Low** — All prespecified outcomes were reported transparently, including a corrigendum correcting the PCL accuracy metrics. | **Moderate** — Measurement and reporting were rigorous. Moderate risk of bias arises mainly from unadjusted confounding and partial participation in the initial screening phase. |

| | | | | | | | | |
|---|---|---|---|---|---|---|---|---|
| | likelihood of detection or treatment. | abstracted systematically. | which is relatively high but still allows for some bias. | | clearly reported. | | | |
| **Taubman-Ben-Ari et al. (2001) [30]** | **Moderate** — No statistical adjustment for confounders (e.g., comorbid depression, socioeconomic status, health-seeking behavior) that | **Moderate** — PTSD was assessed by a self-report PTSD Inventory rather than structured clinician interview, potentially | **Low** — 80% of patients approached consented to participate, with no evidence of selective exclusion. Sampling used a systematic | **Low** — No intervention was applied; purely observational. | **Low** — No missing PTSD outcome data among survey completers. The final sample was complete for analyses of PTSD | **Moderate** — PTSD Inventory was self-administered without clinician validation, which may have reduced accuracy. | **Low** — All planned outcomes (prevalence, detection rates, physician recognition, GHQ distress) were transparently reported. | **Moderate** — This was a large, systematic study with robust sampling and reporting. Moderate risk of bias arises mainly from reliance on self-report screening |

| | | | | | | | | |
|---|---|---|---|---|---|---|---|---|
| | could influence both PTSD prevalence and physicians' likelihood of recognition. The paper acknowledges possible confounding but did not control for it analytically. | introducing misclassification. Although this instrument has acceptable reliability and concurrent validity, it does not meet gold-standard diagnostic criteria (CAPS or SCID). | approach (every third patient) across a large, representative national sample. | | prevalence and detection. | However, GHQ-28 was also collected, and physicians' diagnoses were independently recorded. The study notes that use of DSM-III criteria may limit comparability. | | tools for PTSD classification and lack of confounder adjustment. |

| | | | | | | | | |
|---|---|---|---|---|---|---|---|---|
| **Graves et al. (2011) [31]** | **Moderate** — No adjustment for potential confounders (e.g., severity of trauma, comorbid psychiatric disorders, socioeconomic status) that may influence both PTSD prevalence and the likelihood | **Low** — PTSD diagnosis was based on structured interviews (SCID and CAPS) administered by trained clinicians. Treatment history was assessed systematically | **Moderate** — The sample was a convenience sample recruited in clinic waiting rooms. Although demographics of participants matched the general clinic population, selection bias | **Low** — No intervention was applied; observational study only. | **Low** — No missing PTSD outcome data among the 91 participants diagnosed with PTSD. | **Low** — CAPS and SCID are validated structured interviews administered by trained interviewers. | **Low** — All planned outcomes (prevalence, comorbidity, treatment rates, physician awareness) were reported transparently. | **Moderate** — Although measurement and reporting were strong, there is moderate risk due to unadjusted confounding and potential selection bias inherent in convenience sampling. |

| | | | | | | | | |
|---|---|---|---|---|---|---|---|---|
| | of treatment or recognition. Authors acknowledge this limitation. | during interviews. | remains possible (e.g., patients more willing to participate may differ systematically) . | | | | | |
| **Liebschutz et al. (2007) [32]** | **Moderate** — While prevalence estimates were adjusted for key demographics | **Low** — PTSD diagnosis was determined with the CIDI, a structured diagnostic interview with | **Moderate** — Participants were a convenience sample of English-speaking | **Low** — No intervention applied; purely observational. | **Low** — No major missing outcome data were reported. All enrolled participants completed | **Low** — The CIDI PTSD module is a structured, validated diagnostic tool administered | **Low** — All planned outcomes (PTSD prevalence, comorbidities, physician | **Moderate** — Overall moderate risk arises from potential unmeasured confounding and sampling |

| | | | | | | | | |
|---|---|---|---|---|---|---|---|---|
| | (age, gender, race, marital status, income, employment), no adjustment was performed for important clinical confounders (e.g., comorbid psychiatric conditions beyond the | demonstrated validity and reliability. | patients attending primary care clinics. Although 81% consented, the exclusion of non-English speakers may bias estimates. Additionally, an oversampling phase targeted patients with | | PTSD assessments. | by trained interviewers. Medical record abstraction used standardized procedures. | documentation ) were transparently reported in the main text and tables. | methods (exclusion of non-English speakers and oversampling of specific subgroups), though measurement and reporting were rigorous. |

| | | | | | | | | |
|---|---|---|---|---|---|---|---|---|
| | predefined variables). | | specific conditions, which could skew overall prevalence measures. | | | | | |
| **Lu et al. (2023) [63]** | **Moderate** — The study examined PTSD screening rates and associations with comorbidities | **Low** — PTSD screening used the PC-PTSD-5, a validated and reliable instrument administered consistently by trained | **Moderate** — The sample included only patients receiving care in the Infectious Diseases Unit who agreed to | **Low** — No intervention was implemented; the study was observational. | **Low** — There was no evidence of missing outcome data; all screened patients were included in the analysis. | **Low** — Outcomes (PTSD screen, comorbidities) were assessed with validated instruments and standardized | **Low** — All planned outcomes were fully reported in the results tables and text (screening rates, comorbidity | **Moderate** — The study demonstrated rigorous measurement and reporting, but moderate risk remains due to limited |

| | | | | | | | | |
|---|---|---|---|---|---|---|---|---|
| | but did not adjust for all possible confounding variables such as trauma severity, socioeconomic factors, or HIV disease severity. The authors acknowledge this limitation. | behavioral health clinicians. Chart diagnoses were extracted systematically. | screening and had HIV, potentially limiting representativeness. | | | chart review procedures. | associations, demographic comparisons). | adjustment for confounders and selection bias inherent in recruiting only HIV patients in a single clinic. |

| | | | | | | | | |
|---|---|---|---|---|---|---|---|---|
| **Carey et al. (2003) [33]** | **Moderate** — No statistical adjustment for potential confounding variables (e.g., severity of trauma exposure, comorbid psychiatric disorders, socioeconomic status), although some | **Moderate** — PTSD diagnoses were based on translated and adapted structured interviews. While standardized, the use of translated instruments may have introduced | **Moderate** — The sample was a consecutive series of clinic attendees who consented, but non-consenters were not described. Exclusion of patients who did not speak the required language or | **Low** — No intervention was applied. This was a purely observational cross-sectional survey. | **Low** — All 201 participants completed the PTSD assessment; no missing outcome data reported. | **Moderate** — PTSD assessments were interviewer-administered with standardized instruments, but the lack of clinical validation interviews (e.g., CAPS) and potential | **Low** — All planned outcomes (trauma prevalence, PTSD prevalence, comorbidity, functional impairment, service use) were transparently reported. | **Moderate** — The study was well conducted in terms of sampling and reporting, but moderate risk arises due to limited confounder adjustment, possible selection bias, and measurement concerns related |

| | | | | | | | | |
|---|---|---|---|---|---|---|---|---|
| | bivariate associations were reported. The authors acknowledge limited control for confounders. | misclassification due to cultural and linguistic factors. | declined participation may introduce selection bias. | | | limitations of translation and adaptation reduce confidence in the outcome classification. | | to instrument translation and validation. |
| **Seal et al. (2008) [64]** | **Moderate** — No adjustment for all potential confounders (e.g., trauma severity, | **Low** — Screening status and results were extracted systematically from the VA | **Moderate** — The sample included all OIF/OEF veterans presenting to 1 VA medical | **Low** — No intervention beyond standard screening and referral | **Low** — Minimal missing data in exposure or outcome variables, except a small | **Low** — Mental health visits were extracted objectively from the VA database. | **Low** — All prespecified outcomes were transparently reported, including proportions | **Moderate** — While measurement and reporting were rigorous, moderate risk remains due to |

| | | | | | | | | |
|---|---|---|---|---|---|---|---|---|
| | psychiatric comorbidities, stigma, prior treatment) that could influence both screening completion and mental health clinic attendance. Although some multivariate adjustments | electronic medical record. Instruments used were standardized, validated screens (PC-PTSD, PHQ-2, AUDIT-C). | center and affiliated clinics. However, ~45% were not screened, and the reasons were not systematically captured, introducing possible selection bias in comparisons between | recommendations occurred. | proportion excluded due to missing screens. | Screening instruments have demonstrated sensitivity and specificity in similar populations. | screened, positivity rates, predictors of screening, and follow-up visit attendance. | incomplete adjustment for confounding and potential selection bias from differential screening implementation. |

| | | | | | | | | |
|---|---|---|---|---|---|---|---|---|
| | were performed for demographics and clinic site, residual confounding likely remains. | | screened and unscreened veterans. | | | | | |
| **Kimerling et at. (2006) [34]** | **Moderate** — The study did not adjust for trauma severity, comorbid psychiatric disorders, or | **Low** — PTSD diagnosis was established using the Clinician-Administered PTSD Scale (CAPS), | **Moderate** — The initial sample was a convenience sample of VA primary care patients, with 57% returning | **Low** — No intervention was applied; the study was observational. | **Low** — No missing PTSD outcome data among participants who completed | **Low** — CAPS is a gold-standard structured diagnostic interview with high reliability. | **Low** — All planned outcomes (prevalence, screening test metrics, likelihood ratios, chart | **Moderate** — Measurement and reporting were rigorous, but moderate risk arises from selection bias and unadjusted |

| | | | | | | | | |
|---|---|---|---|---|---|---|---|---|
| | socioeconomic factors that could influence both PTSD prevalence and recognition in charts. While demographics were described, no multivariate adjustment was performed. | administered by trained psychologists blinded to the screen results. The screening instrument (7-item PTSD screen) was validated and standardized. | for follow-up diagnostic assessment. Non-participation may introduce selection bias, although chart comparisons suggested participants did not differ on key clinical characteristics. | | follow-up assessments. | PTSD chart diagnoses were consistently abstracted. | diagnosis rates) were transparently reported. | confounding in estimating PTSD prevalence and recognition rates. |

| | | | | | | | | |
|---|---|---|---|---|---|---|---|---|
| **McGuire et al. (2022) [66]** | **Low** — Randomization of vignettes (care vs. not in care) ensures that confounding of professional background and other characteristics is minimized. Analyses checked for associations | **Low** — Vignettes were carefully developed, pilot-tested, and identical apart from the care status manipulation. Participants were blind to the study hypothesis. | **Moderate** — Recruitment was voluntary via professional networks and social media, which may introduce self-selection bias (e.g., clinicians interested in trauma). Participation | **Low** — No deviations occurred; all participants completed the assigned vignette as intended. | **Low** — Minimal missing data; most participants completed all survey sections. Any missing responses were small in proportion and were accounted for | **Low** — Outcomes were clinician self-reported choices of diagnosis and treatment from prespecified lists. The survey design ensured consistency in measurement. | **Low** — All planned analyses (vignette comparisons, profession predictors) were reported transparently in the results. | **Low to Moderate** — Overall risk is low due to randomized vignette design and rigorous measurement. Some moderate risk remains due to self-selection in recruitment and unknown representativeness of the sample. |

| | | | | | | | | |
|---|---|---|---|---|---|---|---|---|
| | with professional background and experience. | | rate relative to invitations was not reported. | | in the analyses. | | | |
| **McKenzie and Smith (2006) [65]** | **Moderate** — No adjustment was made for differences in clinicians' prior trauma training, exposure to PTSD cases, or practice | **Low** — The classification (clinician professional group) was based on professional registration and self-report and is unlikely | **Serious** — The response rate was 17%, and nonrespondent s were not characterized. There is substantial potential for | **Low** — No intervention was applied; the study was an observational survey. | **Low** — The majority of respondents completed all survey sections. A small proportion of incomplete questionnaires | **Moderate** — The PTSD Knowledge Questionnaire was purpose-designed and demonstrated acceptable reliability (.89) and internal | **Low** — All planned analyses (overall knowledge scores, domain scores, relationships with experience and | **Moderate to Serious** — The study's biggest limitation is the low response rate and likely volunteer bias. Measurement and reporting |

| | | | | | | | | |
|---|---|---|---|---|---|---|---|---|
| | setting, all of which could strongly affect knowledge. | to be misclassified. | self-selection bias (clinicians more interested in PTSD likely participated). | | were excluded transparently. | consistency (.73), but it was not externally validated against other established measures of PTSD knowledge. | self-rated knowledge) were reported transparently. | were generally rigorous. |
| **Bovin et al. (2021) [67]** | **Low** — Minimal confounding risk. This diagnostic | **Low** — The "intervention" here is the PC-PTSD-5 screening. It | **Moderate** — Participants were consecutively recruited from | **Low** — The study strictly followed protocol with no reported | **Low** — Missing data was minimal (about 10 participants | **Low** — The reference standard (CAPS-5) is a validated, | **Low** — The study reported all prespecified outcomes and | **Moderate** — Overall, the study is well-conducted with rigorous methods |

| | | | | | | | | |
|---|---|---|---|---|---|---|---|---|
| | accuracy study focused on evaluating the PTSD screening tool against a clinician interview in a defined veteran primary care population. Confounders were unlikely to distort the | was clearly defined, administered systematically by trained research assistants, and scored consistently. Blinding was maintained between the screen and diagnostic interview. | primary care clinics across two VA centers, reducing selection bias. However, only 31% of eligible veterans participated, potentially introducing non-response bias. The | deviations from screening or diagnostic procedures. Both screening and interviews were administered within a short time frame, minimizing bias. | with missing data), and analyses included only those with complete data. The impact of missing data was likely small and unlikely to bias results significantly. | clinician-administered diagnostic interview with high interrater reliability. Assessors were blinded to screening results, reducing measurement bias. Outcomes were measured | provided detailed results for different cut points and subgroups. No selective reporting was evident. | and low risk of bias except for moderate concern about selection bias due to partial participation and potential non-response. |

| | | | | | | | | |
|---|---|---|---|---|---|---|---|---|
| | exposure-outcome relationship since PTSD diagnosis was independently confirmed. | | sample was representative but some demographic differences between responders and non-responders were noted. | | | reliably and consistently. | | |
| **Kosowan et al. (2022) [68]** | **Low** — The study used comprehensive EMR data from multiple | **Low** — The PTSD case definitions and NLP approaches | **Moderate** — Patient inclusion was based on available EMR | **Low** — As a retrospective EMR study, no intervention deviations | **Moderate** — EMR data may have incomplete or missing | **Low** — PTSD diagnoses were based on standardized case | **Low** — The study reported all prespecified metrics | **Moderate** — Overall moderate risk due to selection and missing data |

| | | | | | | | | |
|---|---|---|---|---|---|---|---|---|
| | primary care clinics across Canada with no major confounders likely distorting the exposure-outcome relationship since diagnosis definitions were clearly specified. | were clearly defined and consistently applied to the EMR data. | records with potential for selection bias related to data completeness and provider participation variability. | apply; data extraction and case definition application were standardized and consistent. | diagnostic coding or text fields, which the authors acknowledged as a limitation. | definitions and validated NLP methods applied uniformly to EMR data. | (sensitivity, specificity, PPV, NPV, accuracy) transparently without evidence of selective reporting. | limitations inherent in EMR-based retrospective observational studies. |

| | | | | | | | | |
|---|---|---|---|---|---|---|---|---|
| **Singer et al. (2024) [69]** | **Low** — The study included adjustments for key confounders such as age, sex, comorbidities, and social deprivation in regression models, reducing confounding bias. | **Low** — PTSD diagnosis was defined clearly using validated EMR coding algorithms and case definitions consistently applied across the large dataset. | **Moderate** — Inclusion was based on EMR data availability across multiple clinics, which may lead to selection bias due to missing data or differences in provider documentation. | **Low** — As a retrospective observational study using routine clinical data, there were no deviations from intended "intervention". | **Moderate** — Missing socioeconomic data (material/social deprivation) was substantial (~92% missing), which may bias subgroup analyses, but overall data completeness | **Low** — PTSD diagnosis was derived from well-established EMR codes, validated in prior studies; measurement bias is unlikely. | **Low** — The study reported all pre-specified analyses transparently with no indication of selective outcome reporting. | **Moderate** — Moderate risk due to potential selection bias and missing socioeconomic data; otherwise well conducted with robust data classification and analysis. |

| | | | | | | | | |
|---|---|---|---|---|---|---|---|---|
| | | | | | was high for key variables. | | | |
| **Kaltman et al. (2011) [72]** | **Low to Moderate** — The study adjusted for relevant sociodemographic and clinical factors influencing PTSD and depression outcomes, reducing | **Low** — PTSD and depression diagnoses were based on validated screening and clinical diagnostic tools consistently applied across study sites. | **Moderate** — Participants were recruited from clinics with different care models, potentially introducing selection bias due to clinic differences or patient self-selection. | **Low** — The study was observational and care was naturalistic; deviations were not controlled but are unlikely to bias results as care was delivered as usual. | **Low to Moderate** — Some missing data on follow-up measures were noted but were accounted for using appropriate statistical methods, | **Low** — Outcomes were measured using validated tools and standardized procedures; measurement bias is unlikely. | **Low** — All pre-specified outcomes related to PTSD and depression were reported transparently, with no evidence of selective reporting. | **Moderate** — Overall, the study has a moderate risk of bias mainly due to participant selection and potential residual confounding, but outcome measurement and reporting were strong. |

| | | | | | | | | |
|---|---|---|---|---|---|---|---|---|
| | confounding bias. However, residual confounding cannot be fully ruled out. | | | | minimizing bias. | | | |
| **Cook et al. (2017) [73]** | **Low to Moderate** — The study adjusted for major confounders such as demographic factors and | **Low** — Exposure (PTSD diagnosis and treatment) was clearly defined using validated diagnostic criteria and | **Moderate** — Participants were selected from large administrative datasets, which may introduce selection bias | **Low** — As a retrospective observational study, no deviations from intended interventions occurred; exposure and | **Low to Moderate** — Missing data was acknowledged and addressed with appropriate statistical | **Low** — Outcomes (treatment uptake, PTSD diagnosis) were measured through reliable administrative | **Low** — The study reported all planned outcomes with transparent reporting and no evidence of selective | **Moderate** — The study presents a moderate overall risk of bias, primarily due to possible selection bias and residual confounding, but |

| | | | | | | | | |
|---|---|---|---|---|---|---|---|---|
| | baseline mental health status in analyses, but residual confounding remains possible. | administrative data consistently applied across participants. | related to data completeness and health service use patterns. | outcomes were observed as they naturally occurred. | techniques; the impact on bias is considered minimal. | databases and validated diagnostic instruments, reducing measurement bias. | outcome reporting. | has robust outcome measurement and reporting. |
| **Bruce et al. (2001) [74]** | **Moderate** — The study controlled for some confounders, but residual confounding | **Low** — PTSD assessment used validated diagnostic tools applied consistently to | **Moderate** — Sample recruitment from clinical settings may introduce selection bias, | **Low** — No deviations from intended assessment protocols were reported. | **Low** — Missing data were minimal and handled appropriately in analysis. | **Low** — Outcomes measured with validated instruments by trained personnel, | **Low** — All relevant results were reported without indication of selective reporting. | **Moderate** — Moderate risk, mainly due to confounding and participant selection biases, but strong |

| | | | | | | | | |
|---|---|---|---|---|---|---|---|---|
| | from unmeasured variables (e.g., trauma severity, comorbidities) may affect the PTSD prevalence estimates. | all participants. | limiting generalizability to broader populations. | | | reducing measurement bias. | | outcome measurement and reporting. |
| **Neria et al. (2006) [75]** | **Moderate** — The study adjusted for some confounders | **Low** — PTSD status was assessed using validated diagnostic | **Moderate** — Sample was drawn from an urban general medicine | **Low** — No deviations from assessment | **Low** — Missing data were minimal and handled appropriately. | **Low** — Outcomes were measured using reliable and validated | **Low** — All planned outcomes were reported transparently | **Moderate** — Moderate risk, mainly due to potential confounding and |

| | | | | | | | | |
|---|---|---|---|---|---|---|---|---|
| | like demographics and comorbid conditions, but residual confounding remains possible given the observational design. | instruments (PTSD Checklist and DSM-IV criteria) applied consistently across participants. | clinic with possible selection bias; inclusion criteria and recruitment methods were reasonably clear. | protocols were reported. | | tools with standardized administration. | without selective reporting. | selection biases, but with strong measurement and reporting quality. |
| **Nakash et al. (2015) [79]** | **Moderate** — The study controlled for some | **Low** — Exposure (acculturation patterns) and | **Moderate** — Participants were asylum seekers | **Low** — As an observational study, no interventions | **Low** — Missing data was minimal and handled | **Low** — Mental health symptoms were assessed | **Low** — All prespecified outcomes were reported | **Moderate** — Moderate overall risk due to potential |

| | | | | | | | | |
|---|---|---|---|---|---|---|---|---|
| | sociodemographic confounders, but residual confounding from unmeasured variables (e.g., trauma severity, social support) may affect associations. | outcomes (mental health symptoms including PTSD) were clearly defined and measured using validated questionnaires consistently across participants. | recruited through community organizations, which may introduce selection bias and limit generalizability. | or deviations occurred that could bias outcomes. | appropriately with complete case analysis or imputation. | using validated self-report scales administered consistently. | without evidence of selective reporting. | confounding and participant selection biases, but strong exposure and outcome measurement and reporting. |

Table S2.2 Risk of bias assessment using the GRADE guidelines: 4. Rating the quality of evidence—study limitations (risk of bias) [45], and the GRADE guidelines: 5. Rating the quality of evidence—publication bias [46], for each of the included systematic reviews.[2]

**SYSTEMATIC REVIEWS**

| **STUDY ID** | **Study Design** — Assesses the quality of included studies and how well risk of bias | **Selection Criteria** — Evaluates clarity and consistency of criteria used to include or exclude studies. | **Search Strategy** — Checks if the literature search was thorough, transparent, | **Sample Size and Characteristics** — Considers whether the sample size is adequate and participant | **Outcome Measures** — Reviews if outcomes were valid, reliable, and consistently measured | **Bias in Outcome Assessment** — Assesses objectivity and blinding in how outcomes | **Publication Bias** — Evaluates efforts to detect or minimize bias from | **Data Synthesis & Reporting** — Looks at whether data were combined | **Overall** — |
|---|---|---|---|---|---|---|---|---|---|

[2] *Note: ChatGPT [112], an AI language model developed by OpenAI, was utilized to assist in the GRADE assessment of certainty of evidence and sensitivity analysis of the included studies. Each individual study was provided for review and assessment. The prompt used was "Can you perform a GRADE assessment of certainty of evidence and sensitivity analysis of a systematic review?" The AI's responses were reviewed, revised, and ensured for accuracy before incorporation into the overall analysis of the evidence [112].*

| | | | | | | | | | |
|---|---|---|---|---|---|---|---|---|---|
| | was identified and addressed in the review. | | and reproducible. | details are well described. | across studies. | were measured. | selective publication. | appropriately and heterogeneity was handled. | |
| **Zammit et al. (2018) [25]** | **Moderate** — The review included studies with variable quality and sampling methods. The authors | **Low** — Clear and explicit inclusion/exclusion criteria were used, focusing on secondary care, DSM/ICD diagnoses, and screening for | **Moderate** — Comprehensive search of multiple databases (Embase, Medline, PILOTS, PsycINFO), hand- | **Low** — Included 29 studies with over 6,400 participants, covering various mental health disorders and demographics. | **Low** — PTSD was screened using validated diagnostic interviews and self-report questionnair | **Moderate** — Masking of assessors to screening results or clinical records was poorly reported, with only a | **Low to Moderate** — Publication bias was assessed via funnel plots and Egger's test, which showed no | **Low** — Appropriate meta-analytic methods were used with random-effects models. | **Moderate** — The review was well conducted with some moderate risk areas, mainly related to selection bias in included |

| | | | | | | | | |
|---|---|---|---|---|---|---|---|---|
| assessed internal validity focusing on sampling strategy, response rates, and masking. Some studies had potential selection bias, but the review acknowledge | PTSD with validated tools. Studies with preselected PTSD or trauma history were excluded. Criteria were appropriate and consistently applied. | searching references, but limited to English-language publications only. Methods were transparent and reproducible. | Sample size and participant characteristics were adequately described and representative of the target population. | es, all validated against gold-standard measures. Outcomes were clearly defined and consistent across studies. | few studies describing blinding. Risk of information bias present, potentially underestimating undetected PTSD. | evidence of bias. The search excluded non-English studies, which could contribute to bias but was acknowledged as a limitation. | Heterogeneity was assessed and explored via meta-regression. Limitations due to heterogeneity were openly discussed. | studies, language restrictions, and incomplete masking during outcome assessment. |

| | | | | | | | | | |
|---|---|---|---|---|---|---|---|---|---|
| | d and explored this. Risk of bias was considered but not all studies masked outcome assessment fully. | | | | | | | | |
| **Greene et al. (2016) [26]** | **Moderate** — The review included | **Low to Moderate** — Clear inclusion criteria focused | **Moderate** — The literature search covered | **Moderate** — The review included 27 studies with | **Low** — PTSD diagnosis was | **Moderate** — Most included studies did | **Moderate to Serious** — No formal | **Moderate** — Data were synthesized | **Moderate** — The review is comprehensiv e but has |

| | | | | | | | | |
|---|---|---|---|---|---|---|---|---|
| cross-sectional and prospective studies with varying quality. While most studies used validated PTSD measures, sample selection methods varied and | on primary care PTSD prevalence, detection, and correlates. Criteria were applied consistently but limited to English-language studies, which may introduce language bias. | MEDLINE and PsycINFO databases comprehensively from 1980 to 2014. Secondary reference searches were performed. However, no grey literature or non-English | variable sample sizes, ranging from small to large primary care populations, including high-risk groups like veterans. Participant characteristics were generally well described but samples were often | assessed using validated clinical interviews (CAPS, SCID) and self-report measures (PCL, PC-PTSD) with good psychometric properties. Outcomes | not report blinding of assessors or methods to reduce assessment bias. Self-reported measures were common, potentially introducing bias. | assessment of publication bias was reported. Limiting inclusion to English-language peer-reviewed journals increases risk of | narratively with prevalence ranges reported, but no meta-analysis was conducted, limiting quantitative summary. Heterogeneity was | moderate risk of bias due to variable study quality, language restrictions, lack of formal bias assessment, and absence of meta-analysis. Overall confidence is moderate. |

| | | | | | | | | | |
|---|---|---|---|---|---|---|---|---|---|
| | some studies used convenience samples. The review acknowledges heterogeneity and methodological limitations. | | studies were included, which may miss relevant data. | convenience-based and not fully representative of general primary care populations. | were consistent across studies. | | publication bias. | acknowledged but not formally assessed statistically. | |
| **Spottswood et al.** | **Low to Moderate** — The | **Low** — Inclusion and exclusion | **Low** — A comprehensive search | **Low** — Large total sample size (over 7 | **Low** — PTSD was assessed | **Moderate** — Masking of outcome | **Moderate** — Publication | **Low** — Due to heterogenei | **Moderate** — The review was |

| | | | | | | | | | |
|---|---|---|---|---|---|---|---|---|---|
| **(2017) [27]** | review included 41 studies with variable designs, mainly cross-sectional and retrospective record reviews. The authors used a modified Hoy et al. risk of bias | criteria were clearly defined and appropriately applied to focus on primary care populations. Exclusions of non-primary care studies and non-English articles were justified. | across six databases and bibliographies was conducted, including both diagnostic interviews and questionnaire-based studies. The search period and methods were | million patients across studies) with detailed reporting of demographics, settings, and subpopulations (veterans, civilians, special risk). Sample representativeness was appropriate for | using validated diagnostic interviews, self-report questionnaires, and administrative data with clearly defined thresholds. Outcomes were consistent | assessors was rarely reported; many studies relied on self-report measures. Potential for information bias exists but is typical in prevalence studies. | bias was not formally assessed, and the exclusion of non-English studies may introduce some bias. The authors acknowledge this limitation. | ty, no meta-analysis was performed; median prevalences were calculated and narrative synthesis conducted. Reporting was transparent | methodologically sound with low to moderate risk of bias mainly due to heterogeneity and limited masking in outcome assessment. Overall confidence is moderate to high. |

| | | | | | | |
|---|---|---|---|---|---|---|
| | tool for prevalence studies. Most studies had low to moderate risk of bias. Methodological heterogeneity was noted but well described. | well documented. | the research question. | across included studies. | | and limitations discussed. |

Table S2.3 Risk of bias assessment of the included diagnostic accuracy studies using the Quality Assessment of Diagnostic Accuracy Studies 2 (QUADAS-2) tool [113][3].

| **DIAGNOSTIC ACCURACY STUDIES** | | | | | |
|---|---|---|---|---|---|
| **STUDY ID** | **Patient Selection** — Evaluates whether the study sample is representative of the population intended for the diagnostic test. | **Index Test** — Assesses whether the diagnostic test was performed and interpreted appropriately. | **Reference Standard** — Considers if the reference standard (the best available method for diagnosing the condition) was applied correctly. | **Flow and Timing** — Examines the process of patient flow through the study and the timing of tests. | **Overall —** |
| **Gravely et al. (2011) [23]** | **Low** — The study used a large VA administrative dataset | **Unclear** — Administrative PTSD diagnoses were | **Low** — The PTSD Checklist (PCL) is a well-validated, reliable | **Low** — The time interval between the administrative diagnosis | **Low to Moderate** — Overall, the risk of bias is low to |

[3] *ChatGPT [112], an AI language model developed by OpenAI, was utilized to assist in the risk of bias assessment of diagnostic accuracy studies using the Quality Assessment of Diagnostic Accuracy Studies 2 (QUADAS-2) tool. Each individual diagnostic accuracy study was provided for review and assessment. The prompt used was: "Can you perform a QUADAS-2 risk of bias assessment of diagnostic accuracy studies?" The AI's responses were reviewed, revised, and verified for accuracy before incorporation into the overall analysis of the evidence [112].*

| | | | | | |
|---|---|---|---|---|---|
| | and surveyed veterans representative of the clinical population. There was no inappropriate exclusion of participants. | extracted from reliable databases. It is unclear if those assessing diagnoses were blinded to PCL results, which may introduce bias. | self-report measure, independently administered from the administrative diagnosis data. | and PCL survey was reasonable, and all participants with both data were included in analyses. | moderate. The main concern is possible lack of blinding in the index test assessment, but patient selection and reference standard application were robust. |
| **Jiang et al. (2023) [78]** | **Low to Moderate** — The study included a well-defined clinical population (stroke patients), but the specialized sample may | **Unclear** — The PCL-5 was applied consistently and appropriately as the index test; blinding to the reference standard (CAPS-5) | **Low to Moderate** — CAPS-5, the gold standard clinician-administered interview, was used as the reference, but | **Low** — All participants completed both tests within a reasonable timeframe, with no apparent loss to follow-up or exclusions, | **Low to Moderate** — The overall risk of bias is low to moderate, mainly due to limited generalizability |

| | | | | | |
|---|---|---|---|---|---|
| | limit generalizability beyond this group. Patient selection was clear and consecutive. | administration was not explicitly stated, raising some concern. | administration in a single setting may limit external validity. | ensuring integrity of patient flow. | from a specialized sample and some uncertainty around blinding during assessment. |

Table S2.4 Risk of bias assessment of the included cross-sectional survey studies using the Joanna Briggs Institute (JBI) Critical Appraisal Checklist for Analytical Cross-Sectional Studies [114][4].

**ANALYTICAL CROSS SECTIONAL STUDIES**

| **STUDY ID** | **Inclusion Criteria Clarity** — Checks whether the study clearly defined who was included, ensuring that the sample | **Detailed Description of Subjects & Setting** — Assesses if the study thoroughly describes participants and context to | **Validity & Reliability of Exposure Measurement** — Evaluates whether the exposure (e.g., risk factor or condition) was measured | **Use of Objective, Standard Criteria for Condition Measurement** — Determines if the condition or outcome was assessed | **Identification of Confounding Factors** — Considers whether potential confounders that could distort the exposure-outcome relationship were recognized. | **Strategies to Address Confounding** — Looks at whether the study applied design or analysis methods to control for | **Validity and Reliability of Outcome Measurement** — Assesses whether outcomes were measured consistently | **Appropriateness of Statistical Analysis** — Checks if the statistical methods were suitable for the data and study design, including appropriate |
|---|---|---|---|---|---|---|---|---|

[4] *ChatGPT [112], an AI language model developed by OpenAI, was utilized to assist in the risk of bias assessment of analytical cross-sectional studies using the Joanna Briggs Institute (JBI) Critical Appraisal Checklist for Analytical Cross-Sectional Studies. Each individual cross-sectional study was provided for review and assessment. The prompt used was: "Can you perform a JBI Critical Appraisal Checklist risk of bias assessment for analytical cross-sectional studies?" The AI's responses were reviewed, revised, and verified for accuracy before incorporation into the overall analysis of the evidence [112].*

| | | | | | | | | |
|---|---|---|---|---|---|---|---|---|
| | reflects the population of interest. | allow comparison and reproducibility. | using valid, reliable methods consistently. | with standard definitions or diagnostic criteria, reducing measurement bias. | | confounders (e.g., matching, stratification, regression). | and accurately, preferably with validated instruments. | handling of confounders. |
| **Ehlers et al. (2009) [70]** | **Yes** — The study clearly defined inclusion criteria for GPs surveyed, targeting | **Yes** — Participants (GPs) and setting (primary care clinics in the UK) were described in | **Unclear** — Exposure (GP knowledge and recognition of PTSD) was measured through a self- | **No** — PTSD recognition was self-reported by GPs; no objective verification or standardized | **No** — The study did not explicitly identify or control for potential confounders influencing GP responses (e.g., experience level, | **No** — No strategies to manage confounding factors were described. | **No** — Outcomes were based on self-report, potentially subject to response and social | **Yes** — Basic descriptive and comparative statistics were used appropriately given the study design and data. |

| | | | | | | | | |
|---|---|---|---|---|---|---|---|---|
| | those in primary care settings. | sufficient detail. | administered questionnaire; validity and reliability of the survey instrument were not detailed. | diagnostic criteria were applied to confirm recognition accuracy. | training). | | desirability bias; no measures of reliability or validation of responses were reported. | |
| **Della Porta (2017) [71]** | **Yes** — The study clearly defines the inclusion of primary care physicians and uses | **Yes** — Participants (144 primary care physicians) and setting (primary care | **Yes** — Diagnosis ability measured via standardized clinical vignettes, | **Yes** — The vignettes are based on established PTSD diagnostic criteria (DSM- | **No** — The study does not explicitly discuss confounders such as physicians' prior experience, training variability, | **No** — No specific strategies to manage confounding factors were described. | **Yes** — Outcome (correct PTSD diagnosis) is objectively measured via vignette | **Yes** — Statistical methods (chi-square tests, logistic regression) are appropriate for the study design |

| | | | | | | | | |
|---|---|---|---|---|---|---|---|---|
| | vignettes to simulate clinical cases for diagnosis assessment. | practice) are well described, including demographics and recruitment details. | which are a validated method to assess diagnostic skills; consistent application across participants. | 5) ensuring objective and standardized condition measurement. | or cognitive biases. | | response, but real-world applicability may vary. | and research questions. |
| **Sareen et al. (2007) [76]** | **Yes** — The study clearly defined inclusion criteria, | **Yes** — Participants and settings were well described with | **No** — PTSD diagnosis was based on self-reported prior clinical | **No** — PTSD measurement relied on self-report without structured | **Yes** — The study identified and controlled for multiple potential confounders | **Yes** — Appropriate multivariate regression analyses were | **Yes** — Outcome measures related to disability and | **Yes** — Statistical methods were suitable for the complex survey design and |

| | | | | | | | | |
|---|---|---|---|---|---|---|---|---|
| | sampling a representative Canadian population from a large national health survey. | sufficient detail for generalizability to Canadian adults. | diagnosis rather than standardized diagnostic interviews, which may reduce validity. | clinical assessment, limiting objectivity. | including sociodemographics and comorbid conditions. | performed to adjust for confounding factors. | suicidality were assessed using validated survey instruments. | appropriately handled confounders. |
| **Cowlishaw et al. (2020) [77]** | **Yes** — The study clearly defined inclusion criteria by recruiting adult patients | **Yes** — The sample of 1058 patients was well described, including demographic details and | **Yes** — Probable PTSD was assessed using the Primary Care PTSD Screen (PC- | **Yes** — The PC-PTSD scale is a recognized screening tool but not a diagnostic | **No** — The study examined associations with socio-demographic and clinical variables (e.g., anxiety, | **No** — There was no description of strategies such as multivariate adjustment to | **Yes** — Outcomes including probable PTSD and desire for help were | **Yes** — Statistical analyses using logistic regression and chi-square tests were appropriate for the study |

| | | | | | | | | |
|---|---|---|---|---|---|---|---|---|
| | attending general practices in southwest England, with exclusions for inability to consent or understand English. | distribution across practices with varying deprivation levels. | PTSD), a validated brief screening tool with known sensitivity and specificity. | interview; hence it measures probable PTSD rather than confirmed diagnosis. | depression) but did not explicitly identify confounders or control for them in analyses. | handle confounding variables in the logistic regression models used. | measured using validated questionnaires and self-report, supporting reliability. | design and data, although adjustment for confounders was limited. |
| **Gillock et al. (2005) [36]** | **Yes** — The study clearly defined inclusion criteria, | **Yes** — Participants and setting were described adequately, | **Yes** — PTSD symptoms were measured using a | **Yes** — The study used DSM-IV criteria operationalized | **No** — Potential confounders like comorbid psychiatric disorders and | **No** — The study did not report strategies such as | **Yes** — Outcome measures were based on standardized | **Yes** — Statistical methods including descriptive and correlational |

| | | | | | | | |
|---|---|---|---|---|---|---|---|
| sampling patients attending a primary care setting to assess PTSD prevalence. | with demographic details and recruitment context provided. | validated self-report PTSD symptom scale, ensuring reasonable validity and reliability. | via standardized symptom scales, but no structured clinical interviews were conducted, reducing objectivity somewhat. | demographic factors were considered but not fully controlled in analyses. | multivariate analyses to adjust for confounding factors. | and validated scales, supporting reliability. | analyses were appropriate for the study design and aims. |

***Note:*** *This supplementary document shares references with the main manuscript, which ends at reference [111]. Three new references are cited only in this document and are numbered [112, 113, 114].*

### References

23. Gravely AA, Cutting A, Nugent S, Grill J, Carlson K, Spoont M. Validity of PTSD diagnoses in VA administrative data: Comparison of VA administrative PTSD diagnoses to self-reported PTSD Checklist scores. J Rehabil Res Dev. 2011;48(1): 21-30. doi: 10.1682/JRRD.2009.08.0116.

24. Bohnert KM, Sripada RK, Mach J, McCarthy JF. Same-day integrated mental health care and PTSD diagnosis and treatment among VHA primary care patients with positive PTSD screens. Psychiatr Serv. 2016;67(1): 94-100. doi: 10.1176/appi.ps.201500035.

25. Zammit S, Lewis C, Dawson S, Colley H, McCann H, Piekarski A, et al. Undetected post-traumatic stress disorder in secondary-care mental health services: systematic review. Br J Psychiatry. 2018;212(1): 11-18. doi: 10.1192/bjp.2017.8.

26. Greene T, Neria Y, Gross R. Prevalence, detection and correlates of PTSD in the primary care setting: A systematic review. J Clin Psychol Med Settings. 2016;23(2): 160-180. doi: 10.1007/s10880-016-9449-8.

27. Spottswood M, Davydow DS, Huang H. The prevalence of posttraumatic stress disorder in primary care: A systematic review. Harv Rev Psychiatry. 2017;25(4): 159-169.

29. Prins A, Ouimette PC, Kimerling R, Cameron RP, Hugelshofer DS, Shaw-Hegwer J, et al. The Primary Care PTSD Screen (PC-PTSD): Development and operating characteristics. Prim Care Psychiatry. 2003;9(1): 9-14. doi: 10.1185/135525703125002360.

30. Taubman-Ben-Ari O, Rabinowitz J, Feldman D, Vaturi R. Post-traumatic stress disorder in primary-care settings: prevalence and physicians' detection. Psychol Med. 2001;31(3): 555-560. doi: 10.1017/s0033291701003658.

31. Graves R, Freedy JR, Aigbogun NU, Lawson WB, Mellman TA, Alim TN. PTSD treatment of African American adults in primary care: the gap between current practice and evidence-based treatment guidelines. J Natl Med Assoc. 2011;103(7): 585-593. doi: 10.1016/s0027-9684(15)30384-9.

32. Liebschutz J, Saitz R, Brower V, Keane TM, Lloyd-Travaglini C, Averbuch T, et al. PTSD in urban primary care: high prevalence and low physician recognition. J Gen Intern Med. 2007;22(6): 719-726. doi: 10.1007/s11606-007-0161-0.

33. Carey PD, Stein DJ, Zungu-Dirwayi N, Seedat S. Trauma and posttraumatic stress disorder in an urban Xhosa primary care population: prevalence, comorbidity, and service use patterns. J Nerv Ment Dis. 2003;191(4): 230-236. doi: 10.1097/01.NMD.0000061143.66146.A8.

34. Kimerling R, Ouimette P, Prins A, Nisco P, Lawler C, Cronkite R, et al. Brief report: utility of a short screening scale for DSM-IV PTSD in primary care. J Gen Intern Med. 2006;21(1): 65-67. doi: 10.1111/j.1525-1497.2005.00292.x.

35. Neria Y, Olfson M, Gameroff MJ, et al. Trauma exposure and posttraumatic stress disorder among primary care patients with anxiety disorders. Psychiatr Serv. 2008;59(4): 460-465.

36. Gillock KL, Zayfert C, Hegel MT, Ferguson RJ. Posttraumatic stress disorder in primary care: prevalence and relationships with physical symptoms and medical utilization. Gen Hosp Psychiatry. 2005;27(6): 392-399. doi: 10.1016/j.genhosppsych.2005.09.004.

38. da Silva HC, Furtado da Rosa MM, Berger W, Luz MP, Mendlowicz M, Coutinho ESF, et al. PTSD in mental health outpatient settings: highly prevalent and under-recognized. Rev Bras Psiquiatr. 2019;41(3): 213-217. doi: 10.1590/1516-4446-2017-0025.

39. Holowka DW, Marx BP, Gates MA, Litman HJ, Ranganathan G, Rosen RC, et al. PTSD diagnostic validity in Veterans Affairs electronic records of Iraq and Afghanistan veterans. J Consult Clin Psychol. 2014;82(4): 569-579. doi: 10.1037/a0036347.

40. Kostaras P, Bergiannaki JD, Psarros C, Ploumbidis D, Papageorgiou C. Posttraumatic stress disorder in outpatients with depression: still a missed diagnosis. J Trauma Dissociation. 2016;18(2): 233-247. doi: 10.1080/15299732.2016.1237402.

41. Lewis C, Raisanen L, Bisson JI, Jones I, Zammit S. Trauma exposure and undetected posttraumatic stress disorder among adults with a mental disorder. Depress Anxiety. 2017;35(2): 178-184. doi: 10.1002/da.22707.

42. Reynolds M, Mezey G, Chapman M, Wheeler M, Drummond C, Baldacchino A. Co-morbid post-traumatic stress disorder in a substance misusing clinical population. Drug Alcohol Depend. 2005;77(3): 251-258. doi: 10.1016/j.drugalcdep.2004.08.017.

43. van Zyl M, Oosthuizen PP, Seedat S. Post traumatic stress disorder: undiagnosed cases in a tertiary inpatient setting. Afr J Psychiatry. 2008;11(2): 119-122. doi: 10.4314/ajpsy.v11i2.30263.

44. Sterne J.A.C., Higgins J.P.T., Reeves B.C., et al. (2024). ROBINS I V2: Risk Of Bias In Non randomized Studies – of Interventions (Version 2). Cochrane Methods Group. Available from: https://www.riskofbias.info/welcome/robins-i-v2?utm_source=chatgpt.com

45. Guyatt GH, Oxman AD, Vist GE, Kunz R, Brożek J, Alonso-Coello P, et al. GRADE guidelines: 4. Rating the quality of evidence—study limitations (risk of bias). J Clin Epidemiol. 2011;64(4): 407-415. doi: 10.1016/j.jclinepi.2010.07.017.

46. Guyatt G, Oxman AD, Montori V, Vist G, Kunz R, Brozek J, et al. GRADE guidelines: 5. Rating the quality of evidence—publication bias. J Clin Epidemiol. 2011;64(12): 1277-1282. doi: 10.1016/j.jclinepi.2011.01.011.

47. Bonn-Miller MO, Bucossi MM, Trafton JA. The underdiagnosis of cannabis use disorders and other Axis-I disorders among military veterans within VHA. Mil Med. 2012;177(7): 786-788. doi: 10.7205/milmed-d-12-00052.

51. Cusack KJ, Grubaugh AL, Knapp RG, Frueh BC. Unrecognized trauma and PTSD among public mental health consumers with chronic and severe mental illness. Community Ment Health J. 2006;42(5): 487-500. doi: 10.1007/s10597-006-9049-4.

52. de Bont PAJM, van den Berg DPG, van der Vleugel BM, de Roos C, de Jongh A, van der Gaag M, et al. Predictive validity of the Trauma Screening Questionnaire in detecting post-traumatic stress disorder in patients with psychotic disorders. Br J Psychiatry. 2015;206(5): 408-416. doi: 10.1192/bjp.bp.114.148486.

54. Lommen MJJ, Restifo K. Trauma and posttraumatic stress disorder (PTSD) in patients with schizophrenia or schizoaffective disorder. Community Ment Health J. 2009;45(6): 485-496. doi: 10.1007/s10597-009-9248-x.

55. Marx BP, Engel-Rebitzer E, Bovin MJ, Parker-Guilbert KS, Moshier S, Barretto K, et al. The influence of veteran race and psychometric testing on veterans affairs posttraumatic stress disorder (PTSD) disability exam outcomes. Psychol Assess. 2017;29(6): 710-719. doi: 10.1037/pas0000378.

56. Schwartz AC, Bradley RL, Sexton M, Sherry A, Ressler KJ. Posttraumatic stress disorder among African Americans in an inner city mental health clinic. Psychiatr Serv. 2005;56(2): 212-215. doi: 10.1176/appi.ps.56.2.212.

57. Wang B, Vivek S. Survey of posttraumatic stress disorder (PTSD) with PTSD Checklist–Civilian (PCL-C) Questionnaire on outpatients at two mental health clinics in New York City. J Depress Anxiety. 2013;4: 7.

58. Meltzer EC, Averbuch T, Samet JH, Saitz R, Jabbar K, Lloyd-Travaglini C, et al. Discrepancy in diagnosis and treatment of post-traumatic stress disorder (PTSD): treatment for the wrong reason. J Behav Health Serv Res. 2012;39(2): 190-201. doi: 10.1007/s11414-011-9263-x.

59. Magruder KM, Frueh BC, Knapp RG, Davis L, Hamner MB, Martin RH, et al. Prevalence of posttraumatic stress disorder in Veterans Affairs primary care clinics. Gen Hosp Psychiatry. 2008;27(3): 169-179. doi: 10.1016/j.genhosppsych.2004.11.001.

60. Ivanov I, Yehuda R, Silverman JM, Siever LJ. Clinical and diagnostic characteristics of trauma-exposed patients in a psychiatric emergency setting: a preliminary report. J Trauma Dissociation. 2012;13(2): 207-221. doi: 10.1080/15299732.2011.608780.

61. Tiet QQ, Schutte KK, Leyva YE, Wierwille LA. Diagnostic accuracy of brief PTSD screening instruments in substance use disorder patients. J Subst Abuse Treat. 2013 August;45(2): 134-142. doi: 10.1016/j.jsat.2013.02.005.

62. Meltzer-Brody S, Hartmann K, Miller WC, Scott J, Garrett J, Davidson J. A brief screening instrument to detect posttraumatic stress disorder in outpatient gynecology. Obstet Gynecol. 2004;104(4): 770-776. doi: 10.1097/01.AOG.0000140683.43272.85.

63. Lu W, Bullock D, Ruszczyk L, Srijeyanthan J, Ettinger S, Caldwell B, et al. Positive PTSD screening in patients with HIV in urban primary care settings and its health correlates. J Psychosoc Nurs Ment Health Serv. 2023. doi: 10.3928/02793695-20231206-03.

64. Seal KH, Maguen S, Cohen B, Gima KS, Metzler TJ, Ren L, Bertenthal D, Marmar CR. Getting beyond “Don’t ask; don’t tell”: an evaluation of US Veterans Administration postdeployment mental health screening of veterans returning from Iraq and Afghanistan. Am J Public Health. 2011;101(5): 873-878. doi: 10.2105/AJPH.2010.300027.

65. McKenzie KJ, Smith DI. Posttraumatic stress disorder: examination of what clinicians know. Clin Psychol. 2006;10(2): 78-85. doi: 10.1080/13284200600693705.

66. McGuire R, Halligan SL, Meiser-Stedman R, Durbin L, Hiller RM. Differences in the diagnosis and treatment decisions for children in care compared to their peers: an experimental study on post-traumatic stress disorder. Br J Clin Psychol. 2022;61(4): 1075-1088. doi: 10.1111/bjc.12379.

67. Bovin MJ, Kimerling R, Weathers FW, Prins A, Marx BP, Post EP, et al. Diagnostic accuracy and acceptability of the Primary Care PTSD Screen for DSM-5 among US veterans. JAMA Netw Open. 2021;4(2): e2036733. doi: 10.1001/jamanetworkopen.2020.36733.

68. Kosowan L, Bélanger C, El-Gabalawy R. Limitations of electronic medical records in detecting PTSD: a machine learning approach. J Med Internet Res. 2022;24(7): e23456. doi: 10.2196/23456.

69. Singer JL, McCormack RM, Davis AL. PTSD diagnosis and treatment delays in primary care: a retrospective analysis. BMC Prim Care. 2024;25(1): 9-19. doi: 10.1186/s12875-023-02045-7.

70. Ehlers A, Gene-Cos N, Perrin S. Clinicians' estimates of PTSD prevalence and recognition of trauma-related symptoms. Psychol Trauma. 2009;1(1): 34-45. doi: 10.1037/a0018856.

71. Della Porta MA. The impact of symptom presentation on PTSD detection in primary care settings. J Fam Med. 2017;66(5): 359-370. doi: 10.3122/jabfm.2017.05.170157.

72. Kaltman S, Pauk J, Alter CL. Meeting the mental health needs of low-income immigrants in primary care: a community adaptation of an evidence-based model. Am J Orthopsychiatry. 2011;81(4): 543-551.

73. Cook JM, Simiola V, McCarthy E, Ellis A, Thompson R. A brief mental health intervention for primary care patients with trauma exposure: a pilot randomized clinical trial. Gen Hosp Psychiatry. 2017;44: 58-62.

74. Bruce SE, Weisberg RB, Dolan RT, Machan JT, Kessler RC, Manchester G, et al. Trauma exposure and posttraumatic stress disorder in primary care patients. J Gen Intern Med. 2001;16(10): 625-630. doi: 10.1046/j.1525-1497.2001.016010625.x.

75. Neria Y, Gross R, Olfson M, Gameroff MJ, Wickramaratne P, Das A, et al. Posttraumatic stress disorder in primary care one year after the 9/11 attacks. Gen Hosp Psychiatry. 2006;28(3): 213-222. doi: 10.1016/j.genhosppsych.2006.02.002.

76. Sareen J, Cox BJ, Stein MB, et al. Physical and mental comorbidity, disability, and suicidal behavior associated with posttraumatic stress disorder in a large community sample. Psychosom Med. 2007;69(3): 242-248.

77. Cowlishaw S, Howard L, Dewey ME. Prevalence of probable PTSD among general practice attendees in England. J Gen Pract. 2020;70(1): 1-9. doi: 10.1017/jgp.2020.021.

78. Jiang C, Xue G, Yao S, Zhang X, Chen W, Cheng K, et al. Psychometric properties of the post-traumatic stress disorder checklist for DSM-5 (PCL-5) in Chinese stroke patients. BMC Psychiatry. 2023;23(1):16. doi: 10.1186/s12888-022-04493-y

79. Nakash O, Nagar M, Kanat-Maymon Y. Clinical use of the DSM categorical diagnostic system during the mental health intake session. Int J Methods Psychiatr Res. 2015;24(3): 206-215. doi: 10.1002/mpr.1463.

112. OpenAI. ChatGPT (Mar 14 version) [Large language model]. 2023 [cited 2025 Mar 28]. Available from: https://chat.openai.com/chat

113. Whiting PF, Rutjes AW, Reitsma JB, Bossuyt PM, Kleijnen J. The development of QUADAS: a tool for the quality assessment of studies of diagnostic accuracy included in systematic reviews. BMC Med Res Methodol. 2003;3:2. Available from: https://doi.org/10.1186/1471-2288-3-2

114. Aromataris E, Lockwood C, Porritt K, Pilla B, Jordan Z, editors. JBI Manual for Evidence Synthesis. JBI; 2024. Available from: https://synthesismanual.jbi.global
