## Supplemental Table 3 for "Post-traumatic stress disorder diagnostic accuracy rates in clinical settings: a systematic review and meta-analysis"

### TABLE S3: INSTITUTIONAL REVIEW BOARD APPROVAL FOR EACH OF THE INCLUDED STUDIES.

***Note**: This supplementary document shares references with the main manuscript, which ends at reference [111].*

| STUDY ID | Institutional Review Board (IRB) approval |
|---|---|
| **Bonn-Miller et al. (2012) [47]** | *"All study procedures were approved by the Stanford University Institutional Review Board."* [47, p. 787] |
| **Cusack et al. (2006) [51]** | *"This study was conducted with full approval by the Institutional Review Boards (IRBs) of the University and the DMH, and the study was in compliance with the ethical treatment of human subjects. All participants signed informed consent documents prior to study participation."* [51, p. 490] |
| **da Silva et al. (2019) [38]** | *"The Universidade Federal do Rio de Janeiro Institute of Psychiatry ethics committee approved the study protocol (process 854.811), including the informed consent forms, questionnaires, and procedures for recruiting and interviewing participants, as well as the mechanisms for protecting the participants' privacy, integrity and rights in conformity with Declaration of Helsinki principles."* [38, p. 214] |
| **de Bont et al. (2015) [52]** | *"Participants were recruited in 2011 and 2012 by teams working in mental health organizations in the Netherlands. The local medical ethics committee approved the study (NL36649.029.12;*[…]*"* [52, p. 409]). |

| | |
|---|---|
| **Holowka et al. (2014) [39]** | *"Those who agreed to participate provided informed consent verbally over the telephone in accordance with the research protocol approved by all local Institutional Review Boards and the Human Research Protection Office of the U.S. Army Medical Research and Materiel Command."* [39, p. 3] |
| **Lommen and Restifo (2009) [54]** | The main paper did not mention IRB approval in this study. The author of PTSD diagnostic accuracy in clinical settings did not have access to any supplemental materials provided by the authors of this study [54].<br><br>*"After the study was described to the participants, written informed consent was obtained from all patients preceding participation in the study."* [54, p. 489] |
| **Marx et al. (2017) [55]** | *"Participants provided informed consent verbally over the telephone in accordance with the research protocol approved by all local Institutional Review Boards and the Human Research Protection Office of the U.S. Army Medical Research and Material Command."* [55, p. 712] |
| **Schwartz et al. (2005) [56]** | *"This study was approved by the institutional review board of Emory University and the research oversight committee of Grady Health System."* [56, p. 212]. |
| **Wang and Vivek (2013) [57]** | *"The protocol was approved by the Internal Review Board of the hospital."* [57, p. 2] |
| **van Zyl et al. (2008) [43]** | *"The study was approved by the institutional review board of the University of Stellenbosch."* [43, p. 120] |

| | |
|---|---|
| **Zammit et al. (2018) [25]** | The main paper did not mention IRB approval in this study. This study is a systematic review.<br><br>*"Our protocol (online supplement DS1 available at https://doi.org/10.1192/bjp.2017.8; not pre-registered) followed Meta-analysis Of Observational Studies in Epidemiology (MOOSE) guidelines13 and a PRISMA checklist was completed (online supplement DS2)."* [25, p. 11] |
| **Reynolds et al. (2005) [42]** | The main paper did not mention IRB approval in this study. The author of *PTSD diagnostic accuracy in clinical settings* did not have access to any supplemental materials provided by the authors of this study [42].<br><br>*"Participants were recruited from a specialist in-patient addiction services in South West Thames. All consenting patients were screened for trauma exposure, PTSD and SUD.* […] *Following a full explanation of the study written consent was obtained."* [42, p. 253] |
| **Lewis et al. (2017) [41]** | *"Data were obtained from the NCMH, a Welsh Government-funded Research Centre that investigates neurodevelopmental and psychiatric disorders across the lifespan. The Centre is operated by Cardiff, Swansea, and Bangor Universities in partnership with National Health Service (NHS) Health Boards across Wales."* [41, p. 2] |

| | |
|---|---|
| **Meltzer et al. (2012) [58]** | *"Boston University Medical Center's Institutional Review and HIPAA Privacy Review Boards approved the study."* [58, p. 3] |
| **Kostaras et al. (2016) [40]** | *"All patients gave their written consent, and the study was approved by the ethics committee of the University of Athens Medical School."* [40] |
| **Gravely et al. (2011) [23]** | The main article does not mention IRB approval or informed consent procedures. The authors do not describe any ethical oversight or participant consent, and no supplemental materials that may have accompanied the original publication were accessible to confirm whether IRB approval was obtained |
| **Magruder et al. (2008) [59]** | This study explicitly states that the project and its parent project were approved by the Institutional Review Board (IRB) of the Medical University of South Carolina, as well as by the appropriate IRBs at the participating VA medical centres in Columbia, Tuscaloosa, and Birmingham. Written informed consent was obtained from all participants prior to their participation in the study [59]. |
| **Ivanov et al. (2012) [60]** | *"The study protocol was approved by the local ethics committee, who considered the use of a diagnostic instrument for PTSD as good medical practice and therefore decided that patients were to be informed orally about the study, but not asked to provide an informed consent."* [60, p. 160]. |
| **Tiet et al. (2013) [61]** | |

| | |
|---|---|
| **Bohnert et al. (2016) [24]** | *"The VHA Ann Arbor Healthcare System Institutional Review Board approved this study."* [24, p. 95] |
| **Meltzer-Brody et al. (2004) [62]** | *"This project was reviewed and approved by the Institutional Review Board at the University of North Carolina at Chapel Hill, and study participants gave informed consent before taking part in this study."* [62, p. 771] |
| **Prins et al. (2003) [29]** | This study received approval from the Stanford University Panel on Medical Human Subjects. Participants were recruited from VA clinics and underwent structured diagnostic interviews. While compensation and consent are mentioned, specific language about written or verbal informed consent is limited [29]. |
| **Taubman-Ben-Ari et al. (2001) [30]** | The main paper did not mention IRB approval in this study. The author of *PTSD diagnostic accuracy in clinical settings* did not have access to any supplemental materials provided by the authors of this study [30].<br><br>*"The […] data were from a national study of primary-care attenders from a geographically representative sample of 26 Israeli primary-health care clinics run by the General Sick Fund, which provides health care under universal health care coverage to about 70% of the population nationally.* […] *After signing an informed consent form, respondents completed the General Health Questionnaire-34, PTSD Inventory and* |

| | |
|---|---|
| | *background questionnaire. Physicians completed a patient encounter form blind to patients' questionnaires."* [30, p. 556] |
| **Graves et al. (2011) [31]** | This study states that research assistants obtained written informed consent from participants following approved internal review board protocol. Although the institutional name is not specified, it is clear that ethical review was conducted [31]. |
| **Liebschutz et al. (2007) [32]** | The study was conducted with full approval from the Boston University Medical Center's Institutional Review and HIPAA Privacy Review Boards, and a Certificate of Confidentiality was obtained from the NIH. Written informed consent was obtained from all participants prior to data collection [32]. |
| **Lu et al. (2023) [63]** | The study received IRB approval from a university in the Northeastern U.S. and the participating primary care clinic. The protocol, including chart review and PTSD screening, was conducted in accordance with the approved IRB protocol [63]. |
| **Carey et al. (2003) [33]** | Despite thorough searches of the journal website, clinical trial registries, institutional repositories, and secondary literature, no explicit statement regarding institutional ethical approval or informed consent procedures was found for this study [33]. This may reflect reporting standards of the time, as detailed ethics disclosures were less consistently included in publications from that period. |

| | |
|---|---|
| **Seal et al. (2008) [64]** | *"This study was approved by the Committee on Human Research, University of California, San Francisco, the San Francisco Veterans Administration Medical Center, and the US Department of Defense."* [64, p. 719] |
| **Kimerling et al. (2006) [34]** | *"The Stanford University panel on medical human subjects approved this project."* [34, p. 66]. |
| **McGuire et al. (2022) [66]** | *"The study was given full ethical approval from the University of Bath Research Ethics Committee and the Health Research Authority."* [66, p. 1077] |
| **McKenzie and Smith (2006) [65]** | The main paper did not mention IRB approval in this study. The author of PTSD diagnostic accuracy in clinical settings did not have access to any supplemental materials provided by the authors of this study [65]<br>*"The sample consisted of 154 individuals from three professional groups: general practitioners (n=59), psychologists (n=56), and psychiatrists (n=39). All participants were registered with the Department of Veterans' Affairs (DVA), Victoria, Australia. Registration with DVA requires only that the clinician hold the mandatory qualifications to register in the State. Questionnaires were sent to all psychologists (n=310), and psychiatrists (n=218) registered with DVA, Victoria."* [65, p. 79] |
| **Bovin et al. (2021) [67]** | *"Local institutional review boards approved all study procedures. All participants provided written informed consent."* [67, p. 2] |

| | |
|---|---|
| **Kosowan et al. (2022) [68]** | *"Ethical approval for this study was obtained from the Health Research Ethics Board at the University of Manitoba, approval number HS21053(2017:257)."* [68, p. 5] |
| **Singer et al. (2024) [69]** | *"This study was approved by the Health Research Ethics Board at the University of Manitoba."* [69, p. 436] |
| **Greene et al. (2016) [26]** | *"All procedures followed were in accordance with ethical standards of the responsible committee on human experimentation (institutional and national) and with the Helsinki Declaration of 1975, as revised in 2000. Informed consent was obtained from all patients for being included in the study. No animal or human studies were carried out by the authors for this article."* [26] |
| **Ehlers et al. (2009) [70]** | Despite thorough searches of the journal website, clinical trial registries, institutional repositories, and secondary literature, no explicit statement regarding institutional ethical approval or informed consent procedures was found for this study [70]. |
| **Della Porta (2017) [71]** | A thorough review of the dissertation available at the Philadelphia College of Osteopathic Medicine's digital repository revealed no explicit statement regarding institutional ethical approval or informed consent procedures for this study [71]. |
| **Kaltman et al. (2011) [72]** | *"The Georgetown University Institutional Review Board and the primary care clinics' review committees approved all study procedures."* [72, p. 6] |

| | |
|---|---|
| **Cook et al. (2017) [73]** | *"Study procedures were approved or exempted from review by each healthcare system's Institutional Review Board. Where applicable, a waiver of informed consent was obtained."* [73, p. 1] |
| **Bruce et al. (2001) [74]** | The paper describes that recruitment followed ethical procedures, including providing participants with full study information and obtaining written informed consent [74]. However, after a thorough search, an explicit statement of institutional review board (IRB) or ethics committee approval was not found. |
| **Neria et al. (2006) [75]** | *"The Institutional Review Boards of the Columbia University Medical Center and the New York State Psychiatric Institute approved the study protocol, and all participants gave informed written consent. Subject recruitment started on April 1, 2002, and was completed on January 16, 2003."* [75, p. 214] |
| **Spottswood et al. (2017) [27]** | *"All studies included in this systematic review adhered to ethical standards, including obtaining informed consent from participants. However, specific statements about institutional review board or ethics committee approval were not consistently reported across all included studies."* |
| **Sareen et al. (2007) [76]** | After a thorough review of this systematic review [76], explicit statements regarding institutional review board (IRB) or ethics committee approval and informed consent procedures for the individual studies included in the review were not found. |

| | |
|---|---|
| **Cowlishaw et al. (2020) [77]** | *"Research Involving Human Participants and/or Animals All procedures performed in studies involving human participants were in accordance with the ethical standards of the institutional and/or national research committee (NHS Health Research Authority, REC Reference: 16/WA/0055) and with the 1964 Helsinki Declaration and its later amendments or comparable ethical standards. This article does not contain any studies with animals performed by any of the authors."* [77, p. 433] |
| **Jiang et al. (2023) [78]** | *"All methods were carried out in accordance with relevant guidelines and regulations. The study was approved by the Ethics Committee of General Hospital of Southern Theatre Command (No. 2020–55). Informed consent was obtained from all subjects."* [78, p. 11] |
| **Gillock et al. (2005) [36]** | After a thorough review, explicit statements regarding institutional review board (IRB) or ethics committee approval and informed consent procedures for this study [36] were not found. |
| **Nakash et al. (2015) [79]** | After a thorough review, explicit statements regarding institutional review board (IRB) or ethics committee approval and informed consent procedures for this study [79] were not found. |
