## Supplementary material for "Post-traumatic stress disorder diagnostic accuracy rates in clinical settings: a systematic review and meta-analysis": PRISMA Checklist 2

### PRISMA 2020 Checklist

| Section and Topic | Item # | Checklist item | Location where item is reported |
|---|---|---|---|
| **TITLE** | | | |
| Title | 1 | Identify the report as a systematic review. | Page 1 |
| **ABSTRACT** | | | |
| Abstract | 2 | See the PRISMA 2020 for Abstracts checklist. | Pages 2-3 |
| **INTRODUCTION** | | | |
| Rationale | 3 | Describe the rationale for the review in the context of existing knowledge. | Pages 3-5 |
| Objectives | 4 | Provide an explicit statement of the objective(s) or question(s) the review addresses. | Page 6 |
| **METHODS** | | | |
| Eligibility criteria | 5 | Specify the inclusion and exclusion criteria for the review and how studies were grouped for the syntheses. | Pages 9-10 & Page 12 |
| Information sources | 6 | Specify all databases, registers, websites, organisations, reference lists and other sources searched or consulted to identify studies. Specify the date when each source was last searched or consulted. | Page 6 & Text S11. |
| Search strategy | 7 | Present the full search strategies for all databases, registers and websites, including any filters and limits used. | Pages 7-8 & Text S11. |
| Selection process | 8 | Specify the methods used to decide whether a study met the inclusion criteria of the review, including how many reviewers screened each record and each report retrieved, whether they worked independently, and if applicable, details of automation tools used in the process. | Pages 11-12, 16-19 Fig. 1A, Fig. 1B, Figure S6 & Text S11 |
| Data collection process | 9 | Specify the methods used to collect data from reports, including how many reviewers collected data from each report, whether they worked independently, any processes for obtaining or confirming data from study investigators, and if applicable, details of automation tools used in the process. | Pages 11-12, 16-19 & Text S11 |
| Data items | 10a | List and define all outcomes for which data were sought. Specify whether all results that were compatible with each outcome domain in each study were sought (e.g. for all measures, time points, analyses), and if not, the methods used to decide which results to collect. | Pages 18-19 |
| | 10b | List and define all other variables for which data were sought (e.g. participant and intervention characteristics, funding sources). Describe any assumptions made about any missing or unclear information. | Pages 8-9 |
| Study risk of bias assessment | 11 | Specify the methods used to assess risk of bias in the included studies, including details of the tool(s) used, how many reviewers assessed each study and whether they worked independently, and if applicable, details of automation tools used in the process. | Pages 8, 13-19, 91-93 & Table S2 |
| Effect measures | 12 | Specify for each outcome the effect measure(s) (e.g. risk ratio, mean difference) used in the synthesis or presentation of results. | Pages 17-18 & 78-91 |
| Synthesis methods | 13a | Describe the processes used to decide which studies were eligible for each synthesis (e.g. tabulating the study intervention characteristics and comparing against the planned groups for each synthesis (item #5)). | Pages 10-16 |
| | 13b | Describe any methods required to prepare the data for presentation or synthesis, such as handling of missing summary statistics, or data conversions. | Page 14-16, 18-19 & Pages 79-89 |

| Section and Topic | Item # | Checklist item | Location where item is reported |
|---|---|---|---|
| | 13c | Describe any methods used to tabulate or visually display results of individual studies and syntheses. | Pages 17-18 & 79-91 |
| | 13d | Describe any methods used to synthesize results and provide a rationale for the choice(s). If meta-analysis was performed, describe the model(s), method(s) to identify the presence and extent of statistical heterogeneity, and software package(s) used. | Pages 17-18 & 78-93 |
| | 13e | Describe any methods used to explore possible causes of heterogeneity among study results (e.g. subgroup analysis, meta-regression). | Pages 91-92 |
| | 13f | Describe any sensitivity analyses conducted to assess robustness of the synthesized results. | Pages 92-93 |
| Reporting bias assessment | 14 | Describe any methods used to assess risk of bias due to missing results in a synthesis (arising from reporting biases). | Page 15-16, 18; & Table S2 |
| Certainty assessment | 15 | Describe any methods used to assess certainty (or confidence) in the body of evidence for an outcome. | Pages 92-93 & Table S2 |
| **RESULTS** | | | |
| Study selection | 16a | Describe the results of the search and selection process, from the number of records identified in the search to the number of studies included in the review, ideally using a flow diagram. | Pages 6, 10-12, 13-16 Fig.1A, Fig. 1B, Figure S6 & Text S11 |
| | 16b | Cite studies that might appear to meet the inclusion criteria, but which were excluded, and explain why they were excluded. | Pages 11-12, Fig. 1A, Fig. 1B & Fig. S6 |
| Study characteristics | 17 | Cite each included study and present its characteristics. | Pages 21-78, & Table S1 |
| Risk of bias in studies | 18 | Present assessments of risk of bias for each included study. | Table S2 |
| Results of individual studies | 19 | For all outcomes, present, for each study: (a) summary statistics for each group (where appropriate) and (b) an effect estimate and its precision (e.g. confidence/credible interval), ideally using structured tables or plots. | Pages 21-63, 79-88 Fig. 2, Fig. 3, Fig. 4, Table S1 & Dataset S7 |
| Results of syntheses | 20a | For each synthesis, briefly summarise the characteristics and risk of bias among contributing studies. | Table S1 & Table S2 |
| | 20b | Present results of all statistical syntheses conducted. If meta-analysis was done, present for each the summary estimate and its precision (e.g. confidence/credible interval) and measures of statistical heterogeneity. If comparing groups, describe the direction of the effect. | Pages 78-90, Fig. 2, Fig. 3, Fig.4 |

PRISMA 2020 Checklist

| Section and Topic | Item # | Checklist item | Location where item is reported |
|---|---|---|---|
| | | | & Dataset S7 |
| | 20c | Present results of all investigations of possible causes of heterogeneity among study results. | Pages 91-92 |
| | 20d | Present results of all sensitivity analyses conducted to assess the robustness of the synthesized results. | Pages 91-93 |
| Reporting biases | 21 | Present assessments of risk of bias due to missing results (arising from reporting biases) for each synthesis assessed. | Table S2 |
| Certainty of evidence | 22 | Present assessments of certainty (or confidence) in the body of evidence for each outcome assessed. | Pages 92-93 & Table S2 |
| **DISCUSSION** | | | |
| Discussion | 23a | Provide a general interpretation of the results in the context of other evidence. | Pages 93, 99 |
| | 23b | Discuss any limitations of the evidence included in the review. | Pages 91-92, 103-104 & Table S2 |
| | 23c | Discuss any limitations of the review processes used. | Pages 92-93 & 103-104 |
| | 23d | Discuss implications of the results for practice, policy, and future research. | Pages 93 & 99-103 |
| **OTHER INFORMATION** | | | |
| Registration and protocol | 24a | Provide registration information for the review, including register name and registration number, or state that the review was not registered. | Pages 6-7 |
| | 24b | Indicate where the review protocol can be accessed, or state that a protocol was not prepared. | Page 7 |
| | 24c | Describe and explain any amendments to information provided at registration or in the protocol. | N/A |
| Support | 25 | Describe sources of financial or non-financial support for the review, and the role of the funders or sponsors in the review. | Disclosed in the PLOS ONE submission system (not in the manuscript), per journal policy |
| Competing interests | 26 | Declare any competing interests of review authors. | Declared in the PLOS ONE submission system. |
| Availability of | 27 | Report which of the following are publicly available and where they can be found: template data collection forms; data extracted from included | Page 19 & |

| Section and Topic | Item # | Checklist item | Location where item is reported |
|---|---|---|---|
| data, code and other materials | | studies; data used for all analyses; analytic code; any other materials used in the review. | 104-105 |

*From:* Page MJ, McKenzie JE, Bossuyt PM, Boutron I, Hoffmann TC, Mulrow CD, et al. The PRISMA 2020 statement: an updated guideline for reporting systematic reviews. BMJ 2021;372:n71. doi: 10.1136/bmj.n71
