## Supplementary figures and images for "Post-traumatic stress disorder diagnostic accuracy rates in clinical settings: a systematic review and meta-analysis"

### Initial Review PRISMA Flow Diagram

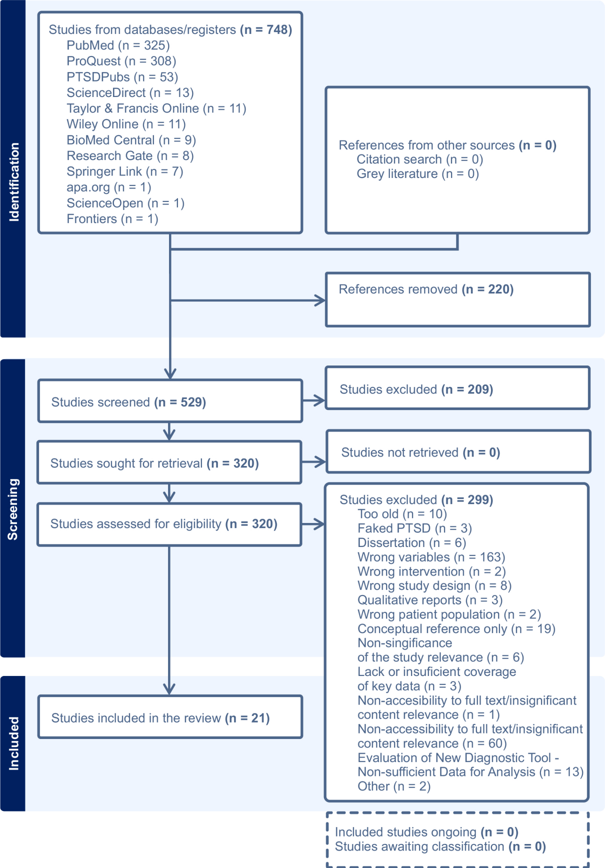
