## Supplementary material for "Post-traumatic stress disorder diagnostic accuracy rates in clinical settings: a systematic review and meta-analysis": Supplememental Dataset

| Study ID | Estimate | LL | UL | Limit | | Negative error | Positive error |
|---|---|---|---|---|---|---|---|
| Bonn-Miller et al. [47] | 0.581 | 0.4073 | 0.7547 | 0.1737 | 20 | 0.1737 | 0.1737 |
| Cusack et al. [51] | 0.116 | 0.0204 | 0.2116 | 0.0956 | 19 | 0.0956 | 0.0956 |
| da Silva et al. [38] | 0.024 | -0.0228 | 0.0708 | 0.0468 | 18 | 0.0468 | 0.0468 |
| de Bont et al. [52] | 0.029 | 0.0018 | 0.0562 | 0.0272 | 17 | 0.0272 | 0.0272 |
| Holowka et al. [39] | 0.853 | 0.8314 | 0.8746 | 0.0216 | 16 | 0.0216 | 0.0216 |
| Lommen and Restifo [54] | 0 | 0 | 0 | 0 | 15 | 0 | 0 |
| Marx et al. [55] | 0.828 | 0.7972 | 0.8588 | 0.0308 | 14 | 0.0308 | 0.0308 |
| Schwartz et al. [56] | 0.115 | 0.074 | 0.156 | 0.041 | 13 | 0.041 | 0.041 |
| Wang and Vivek [57] | 0.211 | 0.0812 | 0.3408 | 0.1298 | 12 | 0.1298 | 0.1298 |
| van Zyl et al. [43] | 0 | 0 | 0 | 0 | 11 | 0 | 0 |
| Zammit et al. [25] | 0.115 | 0.1015 | 0.1285 | 0.0135 | 10 | 0.0135 | 0.0135 |
| Reynolds et al. [42] | 0.05 | -0.046 | 0.146 | 0.096 | 9 | 0.096 | 0.096 |
| Lewis et al. [41] | 0.386 | 0.34 | 0.432 | 0.046 | 8 | 0.046 | 0.046 |
| Meltzer et al. [58] | 0.11 | 0.0569 | 0.1631 | 0.0531 | 7 | 0.0531 | 0.0531 |
| Kostaras et al. [40] | 0.282 | 0.1407 | 0.4233 | 0.1413 | 6 | 0.1413 | 0.1413 |
| Gravely et al. [23] | 0.794 | 0.7768 | 0.8112 | 0.0172 | 5 | 0.0172 | 0.0172 |
| Magruder et al. [59] | 0.43 | 0.332 | 0.528 | 0.098 | 4 | 0.098 | 0.098 |
| Ivanov et al. [60] | 0.375 | 0.2564 | 0.4936 | 0.1186 | 3 | 0.1186 | 0.1186 |
| Tiet et al. [61] | 0.565 | 0.4937 | 0.6363 | 0.0713 | 2 | 0.0713 | 0.0713 |
| Bohnert et al. [24] | 0.644 | 0.6275 | 0.6605 | 0.0165 | 1 | 0.0165 | 0.0165 |

| Study | Estimate | LL | UL | Position | Negative error | Positive error |
|---|---|---|---|---|---|---|
| Gravely et al. [23] | 0.693 | 0.6691 | 0.7169 | 11 | 0.0239 | 0.0239 |
| Bohnert et al. [24] | 0.479 | 0.472 | 0.486 | 10 | 0.007 | 0.007 |
| Meltzer-Brody et al. [62] | 0.12 | -0.0074 | 0.2474 | 9 | 0.1274 | 0.1274 |
| Prins et al. [29] | 0.391 | 0.2103 | 0.5717 | 8 | 0.1807 | 0.1807 |
| Taubman-Ben-Ari et al. [30] | 0.024 | 0.005 | 0.043 | 7 | 0.019 | 0.019 |
| Graves et al. [31] | 0.308 | 0.2131 | 0.4029 | 6 | 0.0949 | 0.0949 |
| Liebschutz et al. [32] | 0.111 | 0.0542 | 0.1678 | 5 | 0.0568 | 0.0568 |
| Lu et al. [63] | 0.239 | 0.1369 | 0.3411 | 4 | 0.1021 | 0.1021 |
| Carey et al. [33] | 0 | 0 | 0 | 3 | 0 | 0 |
| Seal et al. [64] | 0.24 | 0.1761 | 0.3039 | 2 | 0.0639 | 0.0639 |
| Kimerling et al. [34] | 0.382 | 0.219 | 0.545 | 1 | 0.163 | 0.163 |

| Study | Estimate | LL | UL | Limit | Position | Negative error | Positive error |
|---|---|---|---|---|---|---|---|
| McGuire et al. [66] | 0.227 | 0.177 | 0.277 | 0.05 | 2 | 0.05 | 0.05 |
| McKenzie and Smith [65] | 0.74 | 0.6708 | 0.8092 | 0.0692 | 1 | 0.0692 | 0.0692 |
