## Supplementary material for "Post-traumatic stress disorder diagnostic accuracy rates in clinical settings: a systematic review and meta-analysis": R Code Mental Health Settings Meta-analysis

### RStudio Code used to create Random-Effects Meta-analysis forest plots (Mental Health Settings).

```
# Load meta package
> library(meta)
Loading required package: metadat
Loading 'meta' package (version 8.1-0).
Type 'help(meta)' for a brief overview.
>
> # Create dataset with reference numbers only (no study names)
> df_mh <- data.frame(
+     Study = c("[47]", "[51]", "[38]", "[52]", "[39]", "[54]", "[55]", "[56]",
+               "[57]", "[43]", "[25]", "[42]", "[41]", "[58]", "[40]",
+               "[59]", "[60]", "[61]", "[24]"),
+     Events_Correct = c(18, 5, 1, 12, 886, 0, 481, 3, 8, 0, 245, 1, 169, 14, 11, 42, 24,
105, 2107),
+     N_True_PTSD = c(31, 43, 41, 146, 1039, 3, 581, 26, 38, 16, 2135, 20, 438, 133,
39, 98, 64, 186, 3270)
+ )
>
> # Run meta-analysis
> m_mh_nogravely <- metaprop(
+     event = Events_Correct,
+     n = N_True_PTSD,
+     studlab = Study,
+     data = df_mh,
+     sm = "PLOGIT",
+     method = "Inverse",
+     random = TRUE,
+     common = FALSE
+ )
>
> # Open TIFF device (standard PLOS ONE size)
> tiff(
+     filename = "~/Desktop/ForestPlot_MH_OnlyRefs.tiff",
+     width = 8.5,
+     height = 10,
+     units = "in",
+     res = 600,
+     compression = "lzw"
+ )
>
> # Plot forest plot with x-axis label
> forest(
+     m_mh_nogravely,
+     leftcols = c("studlab", "event", "n"),
+     leftlabs = c("Ref", "Events", "N True PTSD"),
+     rightlabs = c("Proportion [95% CI]"),
+     xlab = "Sensitivity (%)",
```

```
+     backtransf = TRUE,
+     digits = 1,
+     fontsize = 10
+ )
>
> # Close device
> dev.off()
null device
          1
```
