## Supplementary material for "Post-traumatic stress disorder diagnostic accuracy rates in clinical settings: a systematic review and meta-analysis": R Code Primary Settings Meta-analysis

### RStudio Code used to create Random-Effects Meta-analysis forest plots (Primary Care Settings).

```
# Load meta package
library(meta)

# Use Times font on Mac (set once)
quartzFonts(TNR = c("Times", "Times", "Times", "Times"))

# Data: Primary Care Settings (reference numbers only)
df_pc <- data.frame(
  Study = c("[24]", "[62]", "[29]", "[30]", "[31]", "[32]", "[63]", "[33]", "[64]", "[34]"),
  Events_Correct = c(8703, 3, 11, 6, 28, 13, 16, 0, 41, 13),
  N_True_PTSD = c(18157, 25, 28, 247, 91, 117, 67, 40, 172, 34)
)

# Run meta-analysis
m_pc <- metaprop(
  event = Events_Correct,
  n = N_True_PTSD,
  studlab = Study,
  data = df_pc,
  sm = "PLOGIT",
  method = "Inverse",
  random = TRUE,
  common = FALSE
)

# TIFF output: PLOS ONE compliant (high-resolution, Times font, readable width)
tiff(
  filename = "~/Desktop/ForestPlot_PrimaryCare.tiff",
  width = 8.5,
  height = 10,
  units = "in",
  res = 600,
  compression = "lzw",
  family = "TNR"
)

# Forest plot
forest(
  m_pc,
  leftcols = c("studlab", "event", "n"),
  leftlabs = c("Ref", "Events", "N True PTSD"),
  rightlabs = c("Proportion [95% CI]"),
  xlab = "Sensitivity (%)",
  backtransf = TRUE,
  digits = 1,
```

```
  fontsize = 10
)

# Close TIFF device
dev.off()
```
