## Supplementary material for "Post-traumatic stress disorder diagnostic accuracy rates in clinical settings: a systematic review and meta-analysis": R Code Clinician Bias/Knowledge Meta-analysis

### RStudio Code used to create Random-Effects Meta-analysis forest plots (Clinicians' PTSD diagnostic competence).

```
# Load meta package
library(meta)

# Study reference numbers only
Study <- c("[66]", "[65]")

# Number of professionals and number correctly identifying PTSD
n_professionals <- c(270, 154)
n_correct <- round(n_professionals * c(0.227, 0.74))

# Run random-effects meta-analysis
m_clinicians <- metaprop(
  event = n_correct,
  n = n_professionals,
  studlab = Study,
  sm = "PLOGIT",
  method = "Inverse",
  random = TRUE,
  common = FALSE
)

# Export high-resolution TIFF forest plot (PLOS ONE compliant)
tiff(
  filename = "~/Desktop/ForestPlot_Clinicians_OnlyRefs.tiff",
  width = 8.5,
  height = 10,
  units = "in",
  res = 600,
  compression = "lzw"
)

# Create the forest plot
forest(
  m_clinicians,
  leftcols = c("studlab", "event", "n"),
  leftlabs = c("Ref", "Events", "N Professionals"),
  rightlabs = c("Proportion [95% CI]"),
  xlab = "Knowledge/Competence (%)",
  backtransf = TRUE,
  digits = 1,
  fontsize = 10,
  print.I2 = TRUE,
  print.tau2 = TRUE,
```

```
  print.Q = TRUE,
  xlim = c(0, 1)
)

# Close the TIFF device
dev.off()
```
