## Supplemental Text - Reconstructed Search Strategy for "Post-traumatic stress disorder diagnostic accuracy rates in clinical settings: a systematic review and meta-analysis"

### S6: RECONSTRUCTED SEARCH STRATEGY.

This document describes the reconstructed search strategy used in this systematic review and meta-analysis, based on details reported in the manuscript and additional information retained by the author.

#### First Search Phase

- Databases searched: PubMed, ProQuest, PTSDPubs, ScienceDirect, Taylor & Francis Online, Wiley Online, BioMed Central, ResearchGate, Springer Link, apa.org, ScienceOpen, Frontiers
- Dates of search: May – June 2023
- Search terms used (combined using Boolean logic):
  - **PubMed:** ("post-traumatic stress disorder"[MeSH Terms] OR PTSD[Title/Abstract]) AND (diagnosis[Title/Abstract] OR "missed diagnosis"[Title/Abstract] OR underdiagnosis[Title/Abstract] OR overdiagnosis[Title/Abstract] OR misdiagnosis[Title/Abstract])
  - **ProQuest (via ProQuest platform):** TI,AB("post-traumatic stress disorder" OR PTSD) AND TI,AB(diagnosis OR "missed diagnosis" OR underdiagnosis OR overdiagnosis OR misdiagnosis)
  - **PTSDpubs (via National Center for PTSD / ProQuest interface):** TI,AB("post-traumatic stress disorder" OR PTSD) AND TI,AB(diagnosis OR "missed diagnosis" OR underdiagnosis OR overdiagnosis OR misdiagnosis)

- **ScienceDirect (via Elsevier interface):** TITLE-ABSTR-KEY("post-traumatic stress disorder" OR PTSD) AND TITLE-ABSTR-KEY(diagnosis OR "missed diagnosis" OR underdiagnosis OR overdiagnosis OR misdiagnosis)
- **Taylor & Francis Online:** "post-traumatic stress disorder" OR PTSD AND diagnosis OR "missed diagnosis" OR underdiagnosis OR overdiagnosis OR misdiagnosis. *(Free-text search).*
- **Wiley Online Library:** "post-traumatic stress disorder" OR PTSD AND diagnosis OR "missed diagnosis" OR underdiagnosis OR overdiagnosis OR misdiagnosis. *(Free-text search).*
- **BioMed Central:** ("post-traumatic stress disorder" OR PTSD) AND (diagnosis OR "missed diagnosis" OR underdiagnosis OR overdiagnosis OR misdiagnosis)
- **ResearchGate:** PTSD post-traumatic stress disorder diagnosis misdiagnosis underdiagnosis overdiagnosis. *(Free-text search).*
- **SpringerLink:** ("post-traumatic stress disorder" OR PTSD) AND (diagnosis OR "missed diagnosis" OR underdiagnosis OR overdiagnosis OR misdiagnosis).
- **APA.org (PsycNet):** ("post-traumatic stress disorder" OR PTSD) AND (diagnosis OR "missed diagnosis" OR underdiagnosis OR overdiagnosis OR misdiagnosis).
- **ScienceOpen:** ("post-traumatic stress disorder" OR PTSD) AND (diagnosis OR "missed diagnosis" OR underdiagnosis OR overdiagnosis OR misdiagnosis).
- **Frontiers:** ("post-traumatic stress disorder" OR PTSD) AND (diagnosis OR "missed diagnosis" OR underdiagnosis OR overdiagnosis OR misdiagnosis).

- Filters applied:

  - Language: English (including translated studies)
  - Publication date: 2000–2023
  - Study types: Observational studies, experimental studies, and reviews
  - Excluded: Animal studies, editorials, opinion pieces
- Search Results by Database (Total n = 748)
  - PubMed (n = 325)
  - ProQuest (n = 308)
  - PTSDPubs (n = 53)
  - ScienceDirect (n = 13)
  - Taylor & Francis Online (n = 11)
  - Wiley Online (n = 11)
  - BioMed Central (n = 9)
  - ResearchGate (n = 8)
  - Springer Link (n = 7)
  - apa.org (n = 1)
  - ScienceOpen (n = 1)
  - Frontiers (n = 1)

**Updated Search Phase**

- Date range: December 2024 – July 2025
- Method: AI-assisted consensus search using OpenAI's ChatGPT with the Consensus plugin
- Search strategy and prompts: To supplement the traditional database search and identify relevant studies published after mid-2023, the author used ChatGPT with access to the

Consensus research database plugin. A detailed description of the review objectives, inclusion criteria, and study context was provided to the AI. The prompts emphasized: "*Find peer-reviewed studies published after 2000 on the diagnostic accuracy of PTSD in clinical settings. Focus on clinician bias, underdiagnosis, misdiagnosis, comorbid mental health conditions, diagnostic delays, and the real-world performance of PTSD diagnostic tools*."

- Selection process:
    - All studies suggested by the AI were manually reviewed for relevance based on predefined inclusion criteria.
    - Studies were included if they provided quantitative data on diagnostic accuracy, clinical decision-making, or tool performance.
    - Studies offering contextual or qualitative insights were included in the qualitative synthesis.
    - Bibliographies of all AI-suggested and previously included studies were manually reviewed for additional relevant sources.
- Contribution of this method: This AI-assisted search strategy enabled broader identification of recent and interdisciplinary research that might be missed by traditional database indexing. It improved the review's comprehensiveness and relevance to current clinical practice.

**Manual Bibliography Review**

- Bibliographies of all included studies — from both the original database search and the AI-assisted search — were manually reviewed.
- Additional studies were identified and included if they met eligibility criteria.

**Notes**

The search strategy above is reconstructed post hoc from manuscript details and documentation retained by the authors. No saved database export strings are available. This reconstruction ensures transparency and reproducibility in line with PRISMA 2020 and PLOS ONE requirements.
